# Risks and etiology of bacterial vaginosis revealed by species dominance network analysis

**DOI:** 10.1101/2020.05.23.20104208

**Authors:** Zhanshan (Sam) Ma, Aaron M. Ellison

## Abstract

BV (bacterial vaginosis) influences 20-40% of women but its etiology is still poorly understood. An open question about the BV is which of the hundreds of bacteria found in the human vaginal microbiome (HVM) are the causal pathogens of BV? Existing attempts to identify them have failed for at least two reasons: (*i*) a focus on species *per se* that ignores species interactions; *(ii)* a lack of systems-level understanding of the HVM. Here, we recast the question of microbial causality of BV by asking if there are any reliable ‘signatures’ of community composition (or poly-microbial ‘cults’) associated with it? We apply a new framework (species dominance network analysis by Ma & Ellison (2019: *Ecological Monographs)* to detect critical structures in HVM networks associated with BV risks and etiology. We reanalyzed the metagenomic datasets of a mixed-cohort of 25 BV patients and healthy women. In these datasets, we detected 15 trio-motifs that occurred exclusively in BV patients. We failed to find any of these 15 trio-motifs in three additional cohorts of 1535 healthy women. Most member-species of the 15 trio motifs are BV-associated anaerobic bacteria (BVAB), Ravel’s community-state type indicators, or the most dominant species; virtually all species interactions in these trios are high-salience skeletons, suggesting that those trios are strongly connected ‘cults.’ The presence of trio motifs unique to BV may act as indicators for its personalized diagnosis and could help elucidate a more mechanistic interpretation of its risks and etiology.

**Single sentence summary:** 15 trio motifs unique to the BV (bacterial vaginosis) may act as indicators for personalized BV diagnosis, risk prediction and etiological study.

## Introduction

BV is a high-recurrence vaginal disease that occurs in 20–40% of sampled women (Koumans *et al*. 2001). It is associated with genital tract infections and many pregnancy complications, including pelvic inflammatory disease, premature rupture of membranes, intrauterine growth restriction, intrauterine fetal demise, chorioamnionitis, endometritis, preterm labor and delivery, postpartum infection, ectopic pregnancy, and tubal factor infertility (White *et al*. 2011). BV is also an independent risk factor for the acquisition and transmission of STDs (sexually-transmitted diseases) and HIV (Ma *et al*. 2012). Beginning in the late 1990s, clinical microbiologists using culture-dependent technology (that could only cultivate and identify very limited number of bacteria) used ecological interpretations (particularly community diversity-stability relationships and species dominance) to interpret BV etiology (Sobel 1999). The advent of metagenomics has made it possible to detect large number of uncultivable microbes in the human vaginal microbiome (HVM) but has focused attention on identifying a single causal agent of BV rather than a more holistic, ecological process (Fredricks *et al*. 2005, 2011, White *et al*. 2011, Ma *et al*. 2012). The recent characterization of BV as “a *syndrome linked to various community types that cause somewhat similar physiological symptoms”* by Ma *et al*. (2012) reflects the limited ecological aspect of the state-of-the-art of research on BV etiology.

The focus on a causal agent for BV asks: which of the hundreds of bacteria found in the HVM are the pathogens of BV? Even though Koch’s postulate of *one pathogen to one disease* has in some cases given way to a paradigm of *polymicrobial disease etiology*, studies of BV have neither identified a single causal pathogen nor a definite polymicrobial ‘signature’ (or what we call here a ‘cult’) of community composition associated with either a woman with a healthy vagina or one with BV. We were tempted to designate some cults of anaerobes in species dominance networks (SDNs) of the HVM as polymicrobial pathogens, but we detected the same cults in the SDN of healthy peers in the same investigated cohort (see Results and Discussion, below) (Ravel *et al*. 2011, 2013, White *et al*. 2011, Gajer *et al*. 2012, Hickey *et al*. 2012, Ma *et al*. 2012, Romero *et al*. 2014, Ma *et al*. 2011, 2015, Ma & Ellison 2018, 2019).

A series of longitudinal studies (Ravel *et al*. 2011, 2013, Gajer *et al*. 2012) together with other global efforts during the last decade (Fredricks *et al*. 2005, 2011, Bradshaw *et al* 2006, Gibbs 2007, Spear *et al*. 2007, Menard *et al*. 2008, Mania-Pramanik *et al*. 2009, Srinivasan *et al*. 2010, Lamont *et al*. 2010, Ling *et al*. 2010, White *et al*. 2011, Aagaard *et al*. 2012, Guedou *et al*. 2012, Jespers *et al*. 2012, Brotman *et al*. 2012, Romero *et al*. 2014, Doyle *et al*. 2018), have greatly expanded our knowledge on HVM and BV. It had been argued that a lack of ‘good’ dominant species in the HVM, especially *Lactobacillus iners*, led to increased risk of BV. This argument was extended to associate HVMs with high community “evenness” (no singularly dominant species) with elevated risk of BV. However, *L. iners* may still be abundant in BV patients and more even HVMs have been reported from asymptomatic women (Ma *et al*. 2012). Ma & Ellison (2018, 2019) re-examined the classic dominance (diversity)-stability relationship (DSR) paradigm (Costello *et al*. 2012, Donohue *et al*. 2016, Ma *et al*. 2019), distinguished between stability and resilience, and introduced a new SDN-based framework for investigating the DSR in the healthy HVM.

We suggest that a universal pathogen(s) or a single diagnostic taxon for BV is unlikely. Furthermore, a single, universal dysbiotic community state is unlikely to cause all BV states. Rather, we hypothesize that BV corresponds to one or more non-equilibrium states in the complex ecosystem that is the HVM (Ma *et al*. 2012) and that there may be multiple paths to one or more types of BV states (henceforth “BV states”). We further hypothesize that despite their multiplicity, BV states are likely to be ecologically equivalent in terms of their decreased ecological stability or resilience (Ma & Ellison 2019). With these hypotheses as a starting point, we predict that it is feasible to detect certain ‘signatures’ of BV states. If the HVM is modeled as a complex network, finding a signature of BV states is reducible to finding a network motif (see **Materials and Methods**, below for a brief presentation of motifs and other network characteristics; and Ma & Ellison 2019 for a more detailed discussion of them). We follow the principle of parsimony and search for simple trio motifs (motifs consisting of three taxa) in complex networks (Ma & Ye 2017). We also harness other characteristics of complex ecological networks (including core/periphery and high-salience structures) point at underlying processes (mechanisms) driving dynamics of vaginal microbiomes and effects of BV as ecological disturbances or perturbations. Integrated together, our motif detection and SDN analyses may be developable as predictors of BV risks and its etiology. Figure 1 illustrates the primary objectives and approaches of this study.

**Fig 1.**
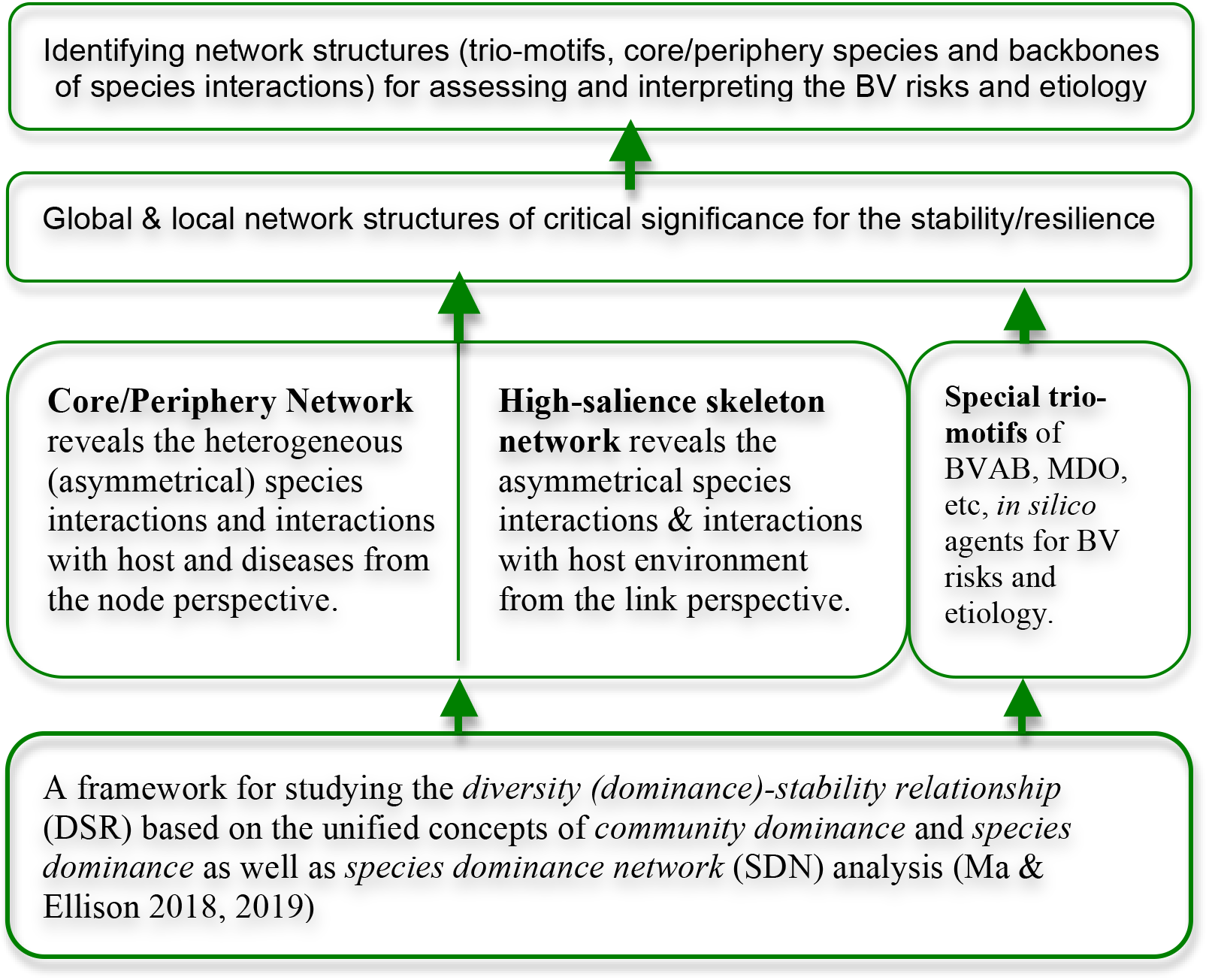
A diagram showing the primary objectives and approaches to achieving the objectives

## Results

### Basic properties of the SDNs

The MDO, MAO, and hubs of each subject are given in Table S1-1. In the whole cohort, the MDO of 11 (out of 25 or 44%) individuals was *Lactobacillus iners*. In the ABV group, the MDO of 5 (out of 6 or 83%) individuals was *Gardnerella vaginalis*, but in the SBV group, the MDO of 9 (out of 15 or 60%) individuals was *L. iners*. The MDOs of the four HEA individuals were all different: *L. iners*, *L. crispatus*, *L. jensenii* and *Bifidobacterium bifidum*. The MAO of 11 (44%) subjects also was *L. iners*, and the MAO and MDO were the same OTU in 23 of the 25 individuals (all but #s112 and #s17). The hub OUTs differed among individuals and the various groups.

Network graphs for all 25 subjects are displayed in Fig. S1. Here, we illustrate network graphs for three exemplar subjects (Figs 1-3) to illustrate basic network topology of the HVM. In these figures, we distinguish the MDO, MAO, and hub; one OTU may play two or all three of these roles. We also distinguish *core* (in cyan) and *periphery* (in azure blue) nodes; negative interactions (red) and positive ones (green). Thick edges identify the *high-salience skeletons* (salience-value *S ≥* 0.25, which measures the strengthen of species interactions).

Individual #s112 had different OTUs for its hub *(Peptoniphilus)*, MDO *(Gardnerella vaginalis)*, and MAO *(L. iners)* (Fig. 2). Although her MAO was *L. iners*, she also had BV (Ravel *et al*. 2013), in contrast with the current interpretation of BV etiology that suggests that high abundance of *L. iners* should lower susceptibility to BV (Ma *et al*. 2012). However, the MDO of individual #s112 was *Gardnerella vaginalis*, an anaerobic species indicative of an altered vaginal environment. Some even consider *Gardnerella vaginalis* as the predominant cause of BV (Bretelle *et al*. 2015), including asymptomatic BV (ABV) (Eren *et al*. 2011, Pleckaityte *et al*. 2012). The hub OTU of individual #s112 was *Peptoniphilus*, considered an indicator of higher risk of BV and of BV state type IV-B (Gajer *et al*. 2012). This example illustrates the utility of distinguishing among different roles (MDO, MAO, hub) in interacting networks of OTUs in the HVM (Ma & Ellison 2018, 2019).

**Fig 2.**
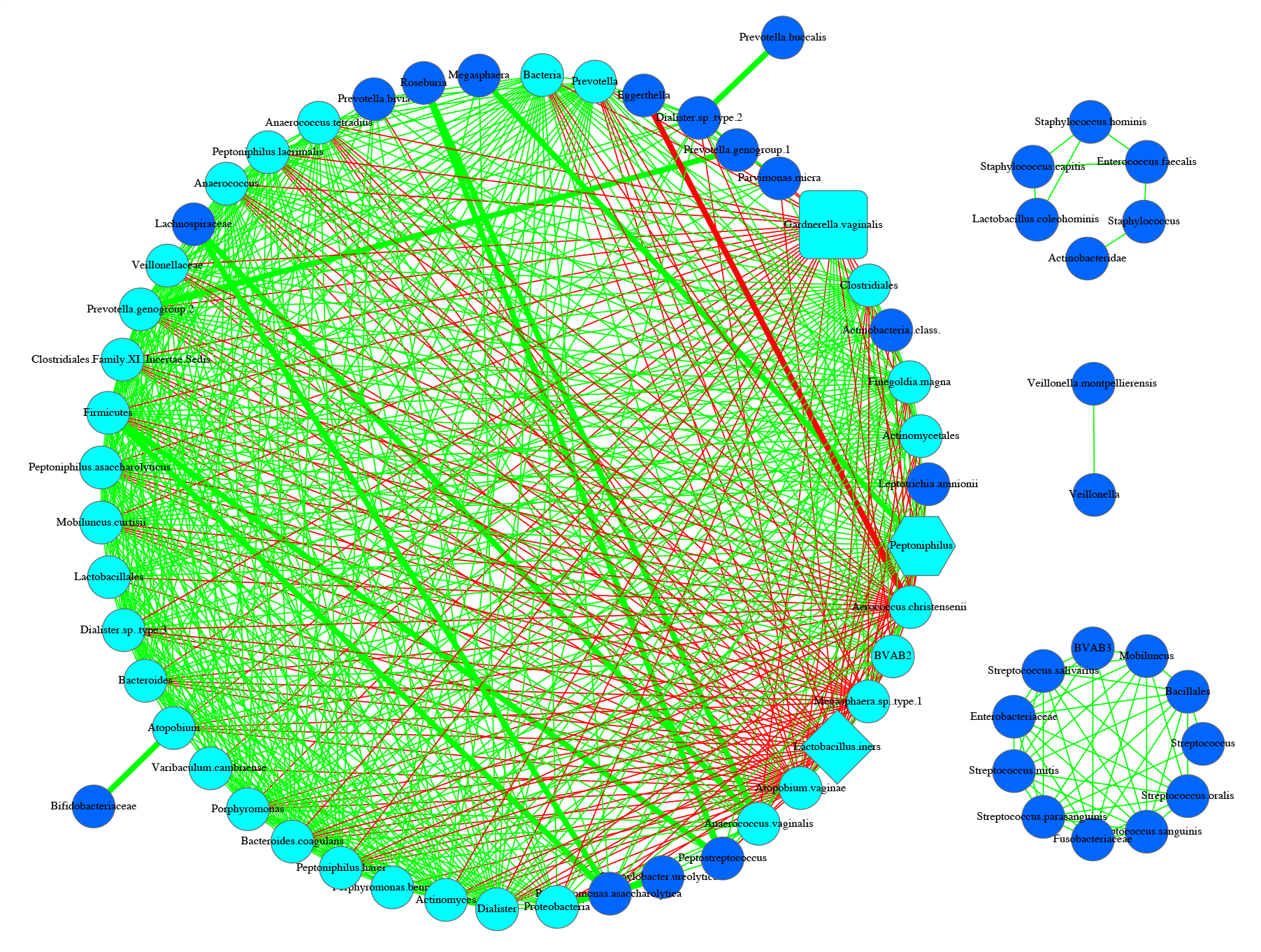
The SDN network graph for individual #s112 in the 25-mixed cohort *(hub=Peptoniphilus, MAO=Lactobacillus iners;* MDO= *Gardnerella vaginalis*; BV Status=SBV; hub, MAO and MDO all belong to the core; core nodes are in cyan color, and periphery nodes are in azure)

Individual #s23 had different dominant OTUs: the hub was *Prevotella buccalis* and *Gardnerella vaginalis* was both the MAO and MDO (Fig. 3). This individual was diagnosed with ABV. *Prevotella* also is one of the indicator species of the BV state type IV-B (Gajer *et al*. 2012). Increased abundance of *Prevotella* in the HVM has been associated to BV, and Randis & Ratner (2019) asserted that *Gardnerella* and *Prevotella* are “*co-conspirators in the pathogenesis of bacterial vaginosis”*.

**Fig 3.**
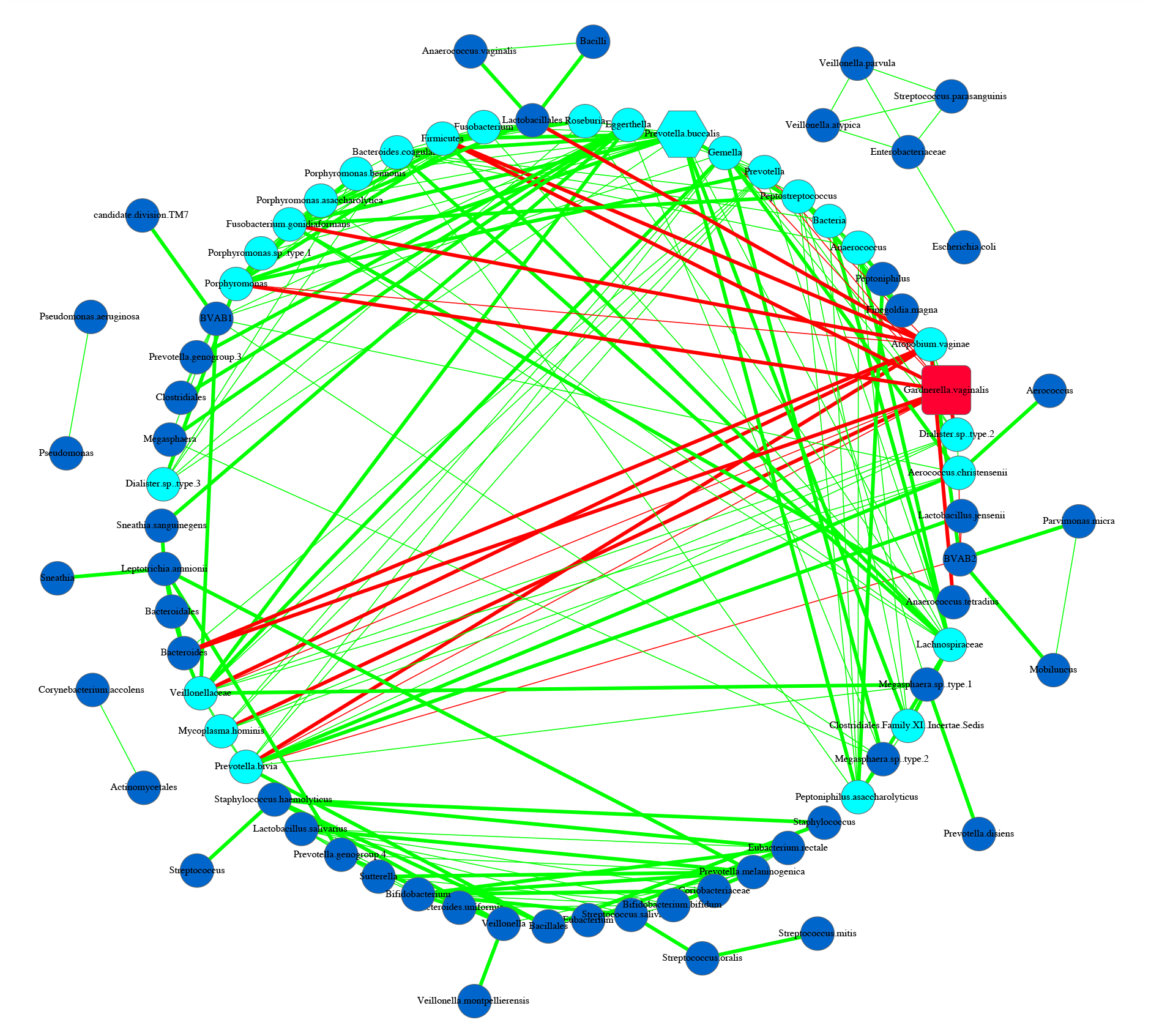
SDN network graph for individual #s23 in the 25-subjects cohort (hub, MDO & MAO are the same node, i.e., *Gardnerella vaginalis*, hub*=Prevotella buccalis;* MDO, MAO and hub all belong to the core; core nodes are in cyan color, and periphery nodes are in azure; BV-status=ABV)

Individual #s52, who was identified as healthy (HEA), had *Lactobacillus jensenii* as her MAO and MDO, and *Clostridiales* Family XI *(incertae sedis)* as her hub species (Fig. 4). Among the four sampled HEA women, the MDOs of three were *L. jensenii*. All negative interactions in the HVM network of #s52 were linked to *L. jensenii* and *L. gasseri*, which suggests their suppressive effects on BVABs and other opportunistic pathogens. Both *Lactobacillus* species are normal inhabitants of the lower reproductive tract in healthy women, whereas *Clostridiales* Family XI *(incertae sedis)* can be an opportunistic pathogen found in women diagnosed with BV (Lambert *et al*. 2013).

**Fig 4.**
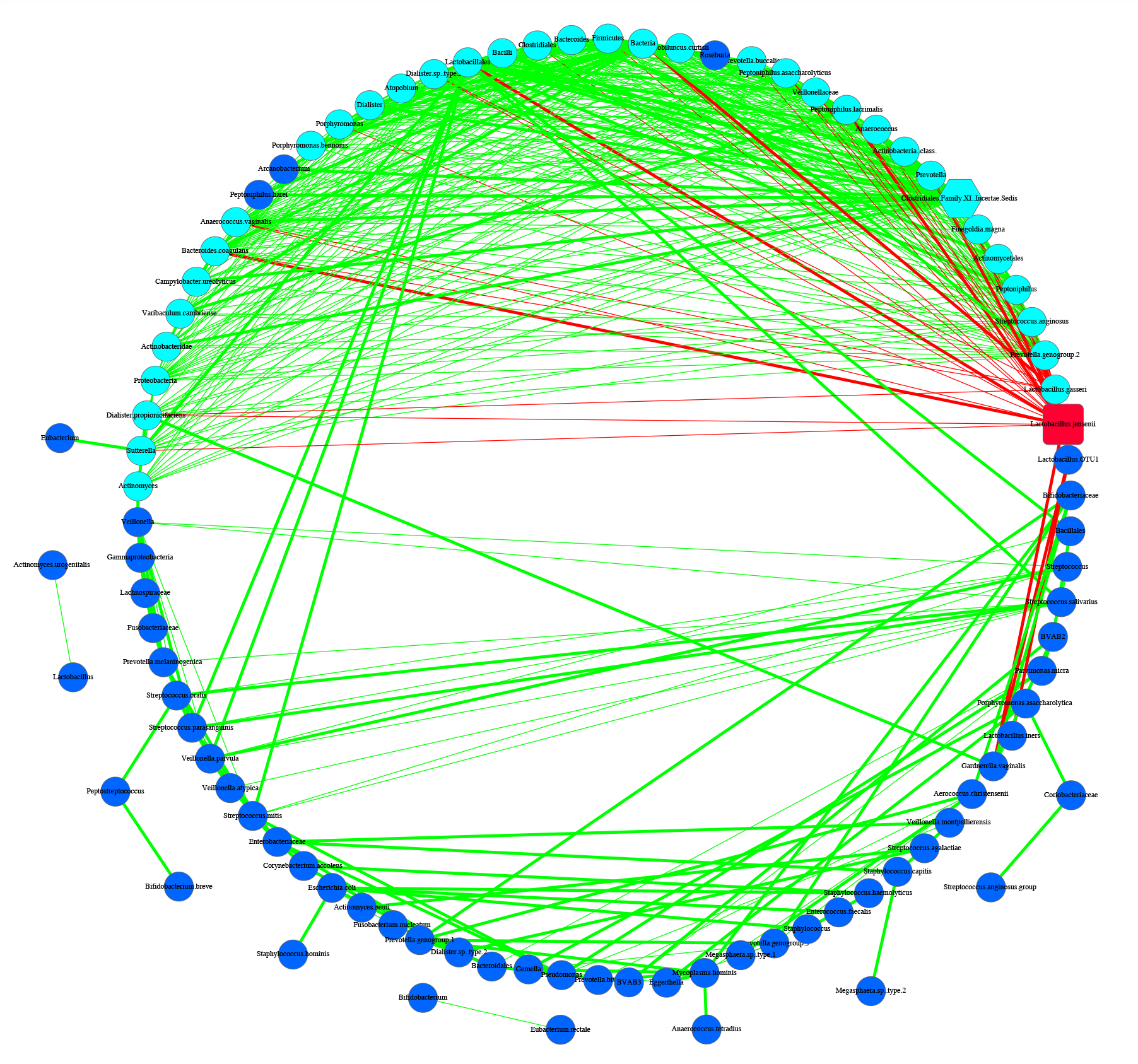
SDN network graph for individual #s52 in the 25-subjects cohort (hub, MDO & MAO are the same node, i.e., *Lactobacillus jensenii*, hub= *Clostridiales* Family XI *(incertae sedis);* MDO, MAO and hub all belong to the core; core nodes are in cyan color, and periphery nodes are in azure; BV-status=HEA)

The basic network properties of the individual SDNs of the 25-mixed cohort and the associated P-value from a Kruskal-Wallis one-way ANOVA are given in Table S1-2. Five network properties, *average degree*, *diameter*, *average path length*, *network density* and *modularity* were significantly different between ABV and SBV individuals (*P*< 0.05). All other network properties were not significantly different among various groups (*P*> 0.05).

Across individuals, SDNs are variable, heterogeneous, and specific to individual women (Figs 2-4, Fig S1, Table S1-2). As each woman has a unique HVM, HVM-associated BV requires personalized diagnosis and treatment. Individual SDN networks can reveal some unique aspects of individual subjects, but from standard network properties (Table S1-2), we can derive limited insights on either HVM heterogeneity or BV etiology.

### Trio motifs and indicators of BV risk

We searched for the trio motifs in the individual SDNs by considering (*i*) both positive and negative interactions; (*ii*) the MDO, MAO, and BVAB; and (*iii*) two types of trio motifs: 3-node *“Trios without handle*” (the three-node trios, *see* Tables S2-1A) and 4-node *“Trios with MDO handle”* (in which the MDO is connected to a three-node trio with one, two, or three links, *see* Table S2-1B). The total number of double-linked MDO (DLM) trios (in which the MDO is connected to three-nodes trio with two links) was significantly different only between ABV and SBV individuals (*P*<0.05). There were no significant differences in all other types of trios in any other comparisons (*P*>0.05). We draw the following findings from this trio motif analysis:

(*i*) 951 trios occurred simultaneously in four or more women in the 25-mixed cohort. The reason we chose counting the trios that occurred in four or more subjects was that the minimal group size of BV, ABV and HEA groups in the cohort was 4 for the HEA group. Among these 951 trios, one trio occurred in 15 women, another trio occurred in 12 women, four trios occurred in 10 women, six occurred in nine women, 16 occurred in eight women, 43 occurred in seven women, 114 occurred in six women, 255 occurred in five women, and 511 occurred in four women (*see* Fig. S2, Tables S2-2, S2-3 for more detailed information on the distribution of these 951 trios among ABV, BV and HEA groups).

As shown in Fig S2, 32% (301) of these 951 trios occurred exclusively in the SBV group, in which 12 trios occurred in near half of the subjects in the SBV group (Table 1). However, no trios were detected exclusively in ABV or HEA without also being detected in another group(s). Another 232 (24%) of the trios were detected in both SBV and ABV groups, in which three trios occurred in ten (50%) of the subjects in the BV (SBV+ABV) groups (Table 1). Finally, 340 trios (36%) occurred in all three groups.

Note that when we state a trio occurred in a group (e.g., SBV), it means that this trio occurred in at least one of the subjects in the group (SBV).

(*ii*) The 12 trios that occurred only in women with SBV potentially could be used as indicators of risk for BV (see Tables S2-3, S2-4 for their biomedical implications). Among these 12 trios, all three OTUs (nodes of the trio) in ten trios were BVABs and most were *core* species (see below for further explanation of the core species) of the individual SDNs. For example, trio #17 occurred in eight women with SBV subjects; its three nodes were *Atopobium vaginae, BVAB2* and *Parvimonas micra*. Interactions between three BVABs were positive (cooperative), but the trio itself was inhibited by the MDO in seven of the eight women. Previous studies also have identified *Atopobium* and *Parvimonas* as indicators of BV-state CST (community state type) 4-B or CST-4B (Gajer *et al*. 2012, Ma & Li 2017). The two others of these 12 trios included *L. iners* and two BVABs. *L. iners* interacted negatively with the two BVABs in all the SDNs that contained these two trios, corroborating previous observations of inhibitory effects of *L. iners* on BVABs.

(*iii*) Three trios were BV-only (BV=SBV+ABV) trios, occurring in 10 of the individuals diagnosed with either SBV or ABV (*see* Table S2-3 for their biomedical implications). Interactions among the three OTUs in these three trios were all positive. Trio #1—consisting of *Dialister* sp. type.2, *Eggerthella*, and *Veillonellaceae*—occurred in 11 women with SBV subjects and 4 women with ABV. These three species also were the core species in the SDNs of all four women with ABV and 8 of the 11 with SBV. Trio #3*—Anaerococcus, Anaerococcus tetradius*, and *Peptoniphilus*—occurred in 6 women with SBV and 4 with ABV. These three species were the core species in 7 of the 10 SDNs. Studies have identified *Anaerococcus* and *Peptoniphilus* as two other indicator species of the BV-state CST4-B (Gajer *et al*. 2012). Trio #5—*Atopobium vaginae, Bifidobacteriaceae* and *Gardnerella vaginalis*—was detected in 7 women with SBV and 3 with ABV. Among their 10 SDNs, *Atopobium vaginae, Bifidobacteriaceae* and *Gardnerella vaginalis* were detected, respectively, as the core species in 7, 8, and 5 of the SDNs. These three BV-only trios merit additional investigation as promising indicators of BV risk (Table S2-4). However, we consider they are more ambiguous indicators than the previous 12 SBV-only trios because they occurred in both SBV and ABV.

(*iv*) We further identified the trios occurred in 50% or more subjects in each of the three diagnostic groups (*see* Table S2-5). Note that the term *occurrence* is used non-exclusively here. In fact, there is no single trio that was detected in all-individual SDNs of the 25-mixed cohort, which simply displays enormous inter-subject heterogeneity of the HVM. For this, here we choose to detect trios occurred in simple majority (≥50%) of each diagnostic group (SVB, ABV or HEA).

In the SBV group, there were 22 trios detected in >50% of the women (≥7 out of 15), in which only one trio (trio #1) occurred in 73% (11/15) women, and 3 occurred in 53% (8/15) women. Among these 22 trios detected in the SBV group: 12 trios occurred exclusively in the SBV group, as described in (*<u>ii</u>*) previously; nine trios occurred in both the SBV and ABV groups, including trios #1, #3 and #5 described in (*iii*) previously; and one trio (trio #6) occurred in all three diagnostic groups including HEA group.

In the ABV group, there were 21 trios detected in 50% or more subjects (≥3 out of 6), in which one trio (#2) occurred in 80% (5/6) subjects, three trios (#1, #3 and #4) occurred in 70% (4/6) subjects. Among these 21 trios, 10 trios also occurred in both the SBV and ABV groups, including # 1, #3 and #5 discussed in (*iii*), and 11 trios occurred in all three diagnostic groups.

In the HEA (healthy) group, there were 244 trios detected in ≥50% (≥ 2 out of 4) subjects. However, only 2 trios (#4 & #72) out of the 244 were detected in 75% (3/4) subjects. Among these 244 trios, 201 trios including (# 4 & #72) were also detected in the SBV and ABV groups, and the remaining 43 occurred in both the HEA and SBV groups.

(*v*) In summary, we identified 15 special trios, including 12 SBV-only trios occurred exclusively in the SBV group and 3 BV-only trios occurred in ≥50% of the BV subjects only (BV=ABV+SBV groups). Table S2-4 provides brief biomedical information on the members of 11the 15 trios. These trios are of potentially significant importance for assessing BV risks and for investigating BV etiology.

**Table 1.**
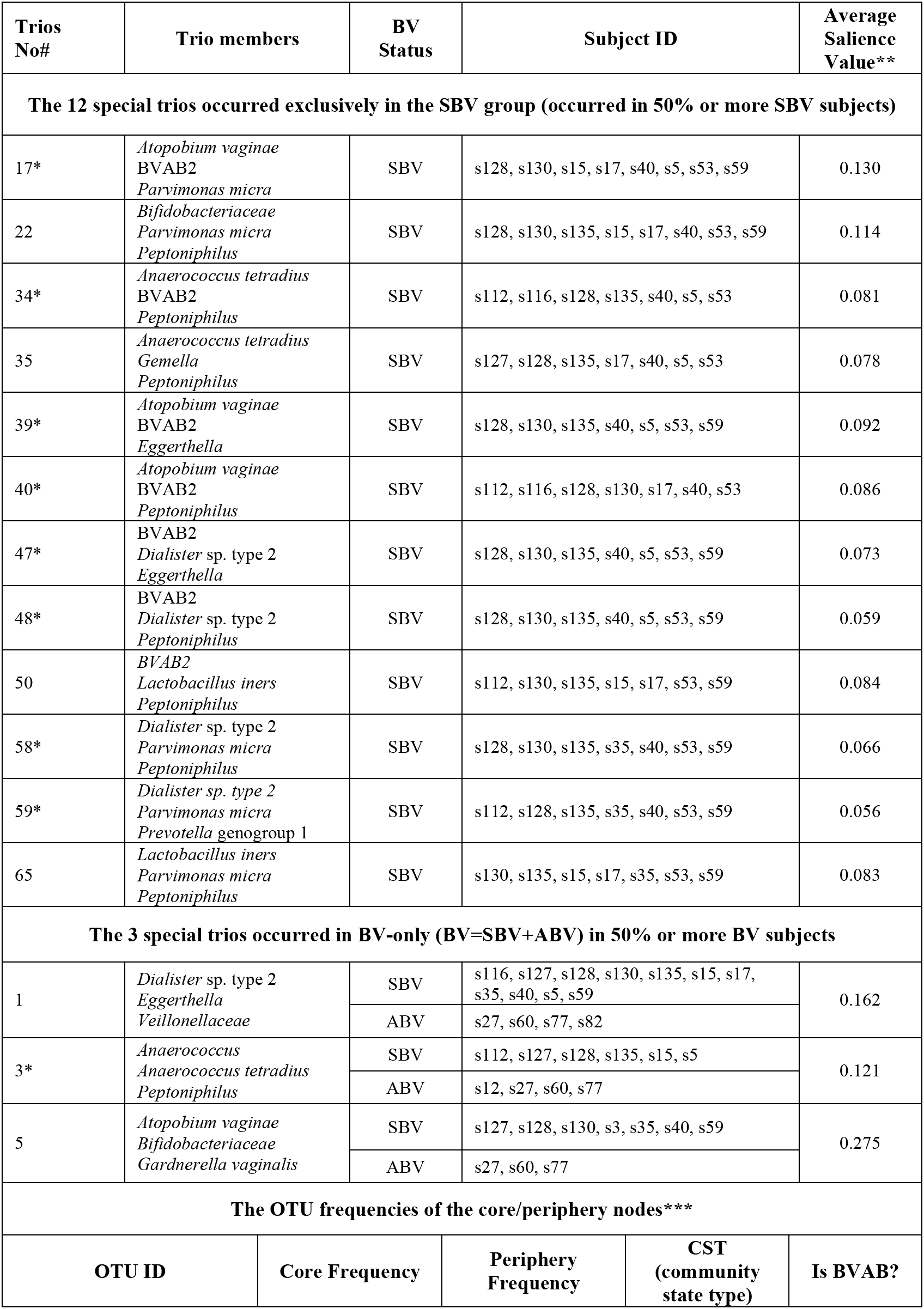
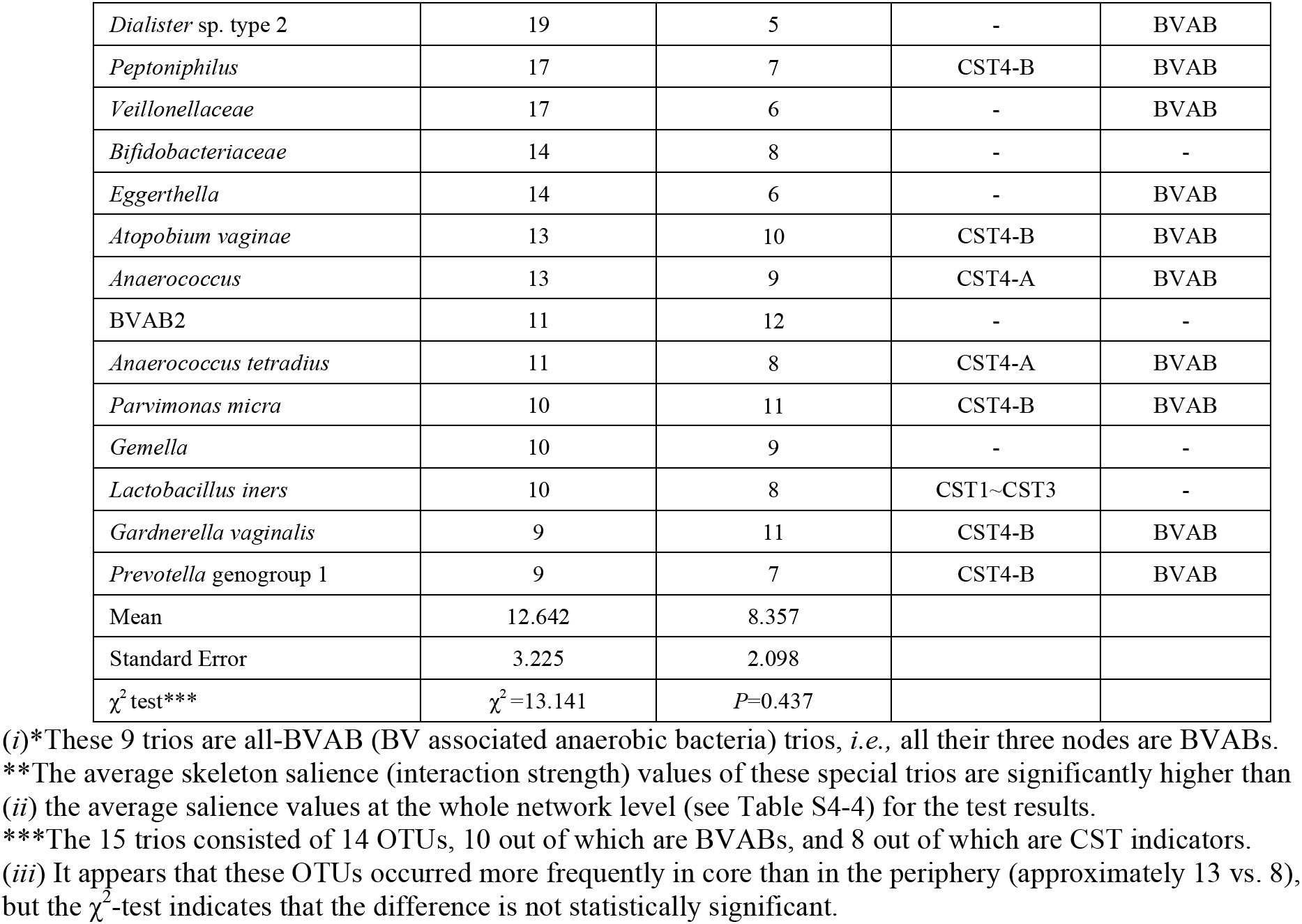
The 15 special trios, including the 12 trios exclusively occurred in the SBV group (SBV-only) and the 3 trios occurred in the BV-only (BV=SBV+ABV) (*see* Table S2-4 for biomedical information on the 15 trios), as well as the relevant core/periphery/skeleton information

| Trios No# | Trio members | BV Status | Subject ID | Average Saliense Value** |  |
| --- | --- | --- | --- | --- | --- |
| The 12 special trios occurred exclusively in the SBV group (occurred in 50% or more SBV subjects) |  |  |  |  |  |
| 17* | *Atopobium vaginae*<br>BVAB2<br>*Parvimonas micra* | SBV | s128, s130, s15, s17, s40, s5, s53, s59 | 0.130 |  |
| 22 | *Bifidobacteriaceae*<br>*Parvimonas micra*<br>*Peptoniphilus* | SBV | s128, s130, s135, s15, s17, s40, s53, s59 | 0.114 |  |
| 34* | *Anaerococcus tetradius*<br>BVAB2<br>*Peptoniphilus* | SBV | s112, s116, s128, s135, s40, s5, s53 | 0.081 |  |
| 35 | *Anaerococcus tetradius*<br>*Gemella*<br>*Peptoniphilus* | SBV | s127, s128, s135, s17, s40, s5, s53 | 0.078 |  |
| 39* | *Atopobium vaginae*<br>BVAB2<br>*Eggerthella* | SBV | s128, s130, s135, s40, s5, s53, s59 | 0.092 |  |
| 40* | *Atopobium vaginae*<br>BVAB2<br>*Peptoniphilus* | SBV | s112, s116, s128, s130, s17, s40, s53 | 0.086 |  |
| 47* | BVAB2<br>*Dialister* sp. type 2<br>*Eggerthella* | SBV | s128, s130, s135, s40, s5, s53, s59 | 0.073 |  |
| 48* | BVAB2<br>*Dialister* sp. type 2<br>*Peptoniphilus* | SBV | s128, s130, s135, s40, s5, s53, s59 | 0.059 |  |
| 50 | *BVAB2*<br>*Lactobacillus iners*<br>*Peptoniphilus* | SBV | s112, s130, s135, s15, s17, s53, s59 | 0.084 |  |
| 58* | *Dialister* sp. type 2<br>*Parvimonas micra*<br>*Peptoniphilus* | SBV | s128, s130, s135, s35, s40, s53, s59 | 0.066 |  |
| 59* | *Dialister* sp. type 2<br>*Parvimonas micra*<br>*Prevotella* genogroup 1 | SBV | s112, s128, s135, s35, s40, s53, s59 | 0.056 |  |
| 65 | *Lactobacillus iners*<br>*Parvimonas micra*<br>*Peptoniphilus* | SBV | s130, s135, s15, s17, s35, s53, s59 | 0.083 |  |
| The 3 special trios occurred in BV-only (BV=SBV+ABV) in 50% or more BV subjects |  |  |  |  |  |
| 1 | *Dialister* sp. type 2<br>*Eggerthella*<br>*Veillonellaceae* | SBV | s116, s127, s128, s130, s135, s15, s17, s35, s40, s5, s59 | 0.162 |  |
|  |  | ABV | s27, s60, s77, s82 |  |  |
| 3* | *Anaerococcus*<br>*Anaerococcus tetradius*<br>*Peptoniphilus* | SBV | s112, s127, s128, s135, s15, s5 | 0.121 |  |
|  |  | ABV | s12, s27, s60, s77 |  |  |
| 5 | *Atopobium vaginae*<br>*Bifidobacteriaceae*<br>*Gardnerella vaginalis* | SBV | s127, s128, s130, s3, s35, s40, s59 | 0.275 |  |
|  |  | ABV | s27, s60, s77 |  |  |
| The OTU frequencies of the core/periphery nodes*** |  |  |  |  |  |
| OTU ID |  | Core Frequency | Periphery Frequency | CST (community state type) | Is BVAB? |

|  |  |  |  |  |
| --- | --- | --- | --- | --- |
| *Dialister* sp. type 2 | 19 | 5 | - | BVAB |
| *Peptoniphilus* | 17 | 7 | CST4-B | BVAB |
| *Veillonellaceae* | 17 | 6 | - | BVAB |
| *Bifidobacteriaceae* | 14 | 8 | - | - |
| *Eggerthella* | 14 | 6 | - | BVAB |
| *Atopobium vaginae* | 13 | 10 | CST4-B | BVAB |
| *Anaerococcus* | 13 | 9 | CST4-A | BVAB |
| BVAB2 | 11 | 12 | - | - |
| *Anaerococcus tetradius* | 11 | 8 | CST4-A | BVAB |
| *Parvimonas micra* | 10 | 11 | CST4-B | BVAB |
| *Gemella* | 10 | 9 | - | - |
| *Lactobacillus iners* | 10 | 8 | CST1~CST3 | - |
| *Gardnerella vaginalis* | 9 | 11 | CST4-B | BVAB |
| *Prevotella* genogroup 1 | 9 | 7 | CST4-B | BVAB |
| Mean | 12.642 | 8.357 |  |  |
| Standard Error | 3.225 | 2.098 |  |  |
| $\chi^2$ test*** | $\chi^2=13.141$ | $P=0.437$ | | |

(i)\*These 9 trios are all-BVAB (BV associated anaerobic bacteria) trios, *i.e.*, all their three nodes are BVABs.
\*\*The average skeleton salience (interaction strength) values of these special trios are significantly higher than (ii) the average salience values at the whole network level (see Table S4-4) for the test results.
\*\*\*The 15 trios consisted of 14 OTUs, 10 out of which are BVABs, and 8 out of which are CST indicators.
(iii) It appears that these OTUs occurred more frequently in core than in the periphery (approximately 13 vs. 8), but the $\chi^2$ -test indicates that the difference is not statistically significant.

### Core/periphery species in the SDNs

Properties of the core/periphery structures for the individual SDNs of the 25-mixed cohort are given in Table S3-1. There were no significant differences in the network properties between women with BV (SBV+ABV) and those without (HEA). The core strength *(ρ*), its size as the proportion of connected network nodes (C/[C+P]), and degree of nestedness (*S*) were significantly larger in SBV than ABV individuals (*P* ≤ 0.05).

Regression analysis of overall network dominance as a function of core/periphery dominance revealed the following relationship:

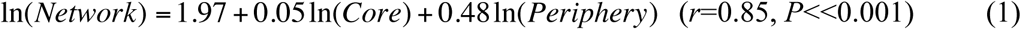

The above model clearly indicates that periphery has a larger effect on community-wide dominance given that the scaling parameter of periphery is nearly 10 times that of core (0.48 *vs*. 0.05). Back-transforming these models gives the exponential relationship between network dominance and core/periphery dominance:

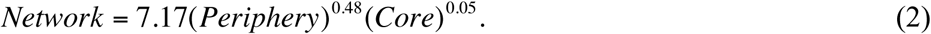

The core/periphery status of individual OTUs varied among individuals (Tables S3-2A, S3-2B). Examination of frequency distributions of OTUs of the HVM in the 25-mixed cohort revealed that although many species occurred in the core (104 OTUs) or periphery (146 OTUs), only a small number of them were exclusive to either (3 exclusive to the core, 42 exclusive to the periphery). Note that the core/periphery status is not fixed in the cohort and can be individual-specific. Interestingly, the most commonly observed species *L. iners*, arguably the most ‘prominent’ species in the HVM is only the 16^th^ most frequent core species and it occurred in 10 individuals (SDNs) only. It also occurred in the periphery of 8 SDNs (8 subjects) and ranked after the 50^th^. This indicates that *L. iners* may not be a universal ‘power’ player in the HVM.

Distribution in the core and periphery of BVAB differed significantly (*P*<0.0001, χ^2^ test; Table S3-3) and were more frequent in the periphery. Table S5 lists all the BVABs recovered from the samples (see also Ravel *et al*. 2012). Table S3-3 also shows that although the BVABs may occur in either core or periphery, they occurred more frequently in the periphery, 314 times in cores vs. 429 in peripheries, or 37% more in the periphery. This finding echoes the previous finding that the periphery is more influential than the core to the network dominance as a whole.

Finally, we analyzed shared core/periphery networks by comparing pairs of subjects from two different groups (ABV, SBV, or HEA; Table S3-4). Using two different algorithms (Ma *et al*. 2019)—the A1 algorithm that reshuffles reads and the A2 algorithm that reshuffles samples, we found that the observed numbers of shared core (or periphery) species were significantly lower than those expected by chance among all three groups (*P*<0.001 all cases). That is, there exist group-specific, unique core (or periphery) species for each of the SBV, ABV and HEA groups (Table S3-5).

### High-salience skeletons in the SDNs

The salience of skeletons measures the strength of interactions in terms of species co-dominance. Table S4-1 gives the properties of the high-salience skeletons in all the SDNs in the 25-mixed cohort; those with salience value ≥0.5 are reported in Table S4-2. In the latter, the two nodes (OTUs) connected by the skeleton are MDOs, MAOs, hubs, BV-state indicators, or BVABs. We also tested the number of shared skeletons between each pair of subjects from the three diagnostic groups (Table S4-3). The average of shared skeletons between the HEA and SBV group, between the HEA and ABV group, between the ABV and SBV group, was 79, 91, and 129, respectively (Table S4-3B), with relative similarity ordered as ABV vs. SBV > ABV vs. HEA > SBV vs. HEA This suggests that BV can change the strength of network links relative to those observed in healthy individuals.

## Conclusions and Discussion

Basic SDN analysis confirmed that human vaginal microbiomes are highly heterogeneous among individuals *(see* also Ma & Ellison 2019) even among women diagnosed with SBV or ABV. This result suggests that it is unlikely that either a universal BV pathogen (diagnostic OTUs or their cults) or a universal dysbiotic BV state will be discovered. Core/periphery and high-salience skeleton network analyses found additional complexities and did not identify any universal clear-cut network structures or properties for diagnosing BV or interpreting BV etiology. Nonetheless, the 15 BV-only trio motifs (especially the 12 found exclusively in women with SBV) have promise for assessing and predicting BV risks and provide new insights about its mechanisms and etiology. In addition to using these special trio motifs as indicators of BV risk, individualized (personalized) analysis of each SDN can offer further mechanistic interpretations for the BV status of a particular individual (Figs. 2-4, Fig S1). Although the presence of *L. iners*, certain BVABs, or their co-occurrence may not be sufficient evidence for determining BV status, the existence of their interactions (special trio motifs) and interaction strengths (measured by skeleton salience) do provide supporting diagnostic evidence.

Further insights can be shed on the ecological mechanisms of BV by the distinguishing core and periphery OTUs, or by the identifications of critical pathway of species interactions (network backbones consisting of high-salience skeletons). Although the vaginal microbiomes of different individuals may have different levels of stability or resilience to various disturbances including BV, BV may indeed change the core strength/size and nestedness of SDNs and the strength of species interactions within an SDN, resulting in a change in its stability or resilience. Therefore, the core/periphery network and high-salience skeleton network (Ma & Ellison 2019) further complemented the findings and insights from special trio motif and inductive inspections on individual SDNs. In conclusion, the 15 BV-only trio motifs, their nodes and links, and the core/periphery/high-salience skeleton structures of their associated SDNs are sufficiently unique to act as indicators of community composition and signaling the potential of dysbiosis associated with BV. Although we are still far from revealing clear-cut risk or diagnostic indicators, our study presents important material candidates (15 BV-only trio motifs) and tools (trio-motif detection technique, augmented with core /periphery/skeleton network analyses) for further deepening our understanding on BV risks and etiology.

Finally, we asked how specific are the 15 BV-only trio-motifs? We answered this question by testing whether the 15 special trio-motifs (listed in Table 1) could be detected in cohorts of healthy women. We used three additional 16S-rRNA datasets of 1535 healthy women to assess specificity. Although there were differeces among the three healthy-cohort datatsets (e.g., longitudinal vs. cross-sectional sampling designs), we were only interested in determining if we detected the special BV-only trio motifs in species dominance networks of healthy individuals. None of the 15 special trio-motifs were detected in any of the species doimnance networks of the healthy women, regardless of sampling design (Table 2). Thus, it appears that the 15 trio-motifs could be a highly specific indicator of BV to be used for its personalized diagnosis and treatment.

**Table 2.** The number of 15 special trios (originally detected in the BV patients and listed in Table 1) that were detected in the networks of HVM-32, HVM-400, and HVM-UK-1107 cohorts

| ID# | Nodes | Edges | #17 | #22 | #34 | #35 | #39 | #40 | #47 | #48 | #50 | #58 | #59 | #65 | #1 | #3 | #5 |
| --- | --- | --- | --- | --- | --- | --- | --- | --- | --- | --- | --- | --- | --- | --- | --- | --- | --- |
| **32-Healthy Cohort (Gajer et al. 2012)** |  |  |  |  |  |  |  |  |  |  |  |  |  |  |  |  |  |
| #400 | 43 | 196 | 0 | 0 | 0 | 0 | 0 | 0 | 0 | 0 | 0 | 0 | 0 | 0 | 0 | 0 | 0 |
| #401 | 69 | 263 | 0 | 0 | 0 | 0 | 0 | 0 | 0 | 0 | 0 | 0 | 0 | 0 | 0 | 0 | 0 |
| #402 | 73 | 523 | 0 | 0 | 0 | 0 | 0 | 0 | 0 | 0 | 0 | 0 | 0 | 0 | 0 | 0 | 0 |
| #403 | 19 | 29 | 0 | 0 | 0 | 0 | 0 | 0 | 0 | 0 | 0 | 0 | 0 | 0 | 0 | 0 | 0 |
| #404 | 65 | 203 | 0 | 0 | 0 | 0 | 0 | 0 | 0 | 0 | 0 | 0 | 0 | 0 | 0 | 0 | 0 |
| #405 | 30 | 90 | 0 | 0 | 0 | 0 | 0 | 0 | 0 | 0 | 0 | 0 | 0 | 0 | 0 | 0 | 0 |
| #406 | 62 | 131 | 0 | 0 | 0 | 0 | 0 | 0 | 0 | 0 | 0 | 0 | 0 | 0 | 0 | 0 | 0 |
| #407 | 71 | 183 | 0 | 0 | 0 | 0 | 0 | 0 | 0 | 0 | 0 | 0 | 0 | 0 | 0 | 0 | 0 |
| #408 | 22 | 27 | 0 | 0 | 0 | 0 | 0 | 0 | 0 | 0 | 0 | 0 | 0 | 0 | 0 | 0 | 0 |
| #410 | 41 | 53 | 0 | 0 | 0 | 0 | 0 | 0 | 0 | 0 | 0 | 0 | 0 | 0 | 0 | 0 | 0 |
| #411 | 46 | 98 | 0 | 0 | 0 | 0 | 0 | 0 | 0 | 0 | 0 | 0 | 0 | 0 | 0 | 0 | 0 |
| #412 | 23 | 23 | 0 | 0 | 0 | 0 | 0 | 0 | 0 | 0 | 0 | 0 | 0 | 0 | 0 | 0 | 0 |
| #413 | 22 | 24 | 0 | 0 | 0 | 0 | 0 | 0 | 0 | 0 | 0 | 0 | 0 | 0 | 0 | 0 | 0 |
| #414 | 24 | 45 | 0 | 0 | 0 | 0 | 0 | 0 | 0 | 0 | 0 | 0 | 0 | 0 | 0 | 0 | 0 |
| #415 | 35 | 112 | 0 | 0 | 0 | 0 | 0 | 0 | 0 | 0 | 0 | 0 | 0 | 0 | 0 | 0 | 0 |
| #416 | 39 | 45 | 0 | 0 | 0 | 0 | 0 | 0 | 0 | 0 | 0 | 0 | 0 | 0 | 0 | 0 | 0 |
| #418 | 54 | 133 | 0 | 0 | 0 | 0 | 0 | 0 | 0 | 0 | 0 | 0 | 0 | 0 | 0 | 0 | 0 |
| #420 | 49 | 117 | 0 | 0 | 0 | 0 | 0 | 0 | 0 | 0 | 0 | 0 | 0 | 0 | 0 | 0 | 0 |
| #423 | 59 | 102 | 0 | 0 | 0 | 0 | 0 | 0 | 0 | 0 | 0 | 0 | 0 | 0 | 0 | 0 | 0 |
| #424 | 84 | 439 | 0 | 0 | 0 | 0 | 0 | 0 | 0 | 0 | 0 | 0 | 0 | 0 | 0 | 0 | 0 |
| #429 | 20 | 13 | 0 | 0 | 0 | 0 | 0 | 0 | 0 | 0 | 0 | 0 | 0 | 0 | 0 | 0 | 0 |
| #430 | 41 | 65 | 0 | 0 | 0 | 0 | 0 | 0 | 0 | 0 | 0 | 0 | 0 | 0 | 0 | 0 | 0 |
| #431 | 11 | 9 | 0 | 0 | 0 | 0 | 0 | 0 | 0 | 0 | 0 | 0 | 0 | 0 | 0 | 0 | 0 |
| #432 | 65 | 317 | 0 | 0 | 0 | 0 | 0 | 0 | 0 | 0 | 0 | 0 | 0 | 0 | 0 | 0 | 0 |
| #435 | 22 | 25 | 0 | 0 | 0 | 0 | 0 | 0 | 0 | 0 | 0 | 0 | 0 | 0 | 0 | 0 | 0 |
| #436 | 74 | 210 | 0 | 0 | 0 | 0 | 0 | 0 | 0 | 0 | 0 | 0 | 0 | 0 | 0 | 0 | 0 |
| #437 | 70 | 316 | 0 | 0 | 0 | 0 | 0 | 0 | 0 | 0 | 0 | 0 | 0 | 0 | 0 | 0 | 0 |
| #439 | 15 | 47 | 0 | 0 | 0 | 0 | 0 | 0 | 0 | 0 | 0 | 0 | 0 | 0 | 0 | 0 | 0 |
| #443 | 75 | 178 | 0 | 0 | 0 | 0 | 0 | 0 | 0 | 0 | 0 | 0 | 0 | 0 | 0 | 0 | 0 |
| #444 | 41 | 28 | 0 | 0 | 0 | 0 | 0 | 0 | 0 | 0 | 0 | 0 | 0 | 0 | 0 | 0 | 0 |
| #445 | 2 | 1 | 0 | 0 | 0 | 0 | 0 | 0 | 0 | 0 | 0 | 0 | 0 | 0 | 0 | 0 | 0 |
| #446 | 51 | 272 | 0 | 0 | 0 | 0 | 0 | 0 | 0 | 0 | 0 | 0 | 0 | 0 | 0 | 0 | 0 |
| **400-Healthy Cohort (Ravel et al. 2011)** |  |  |  |  |  |  |  |  |  |  |  |  |  |  |  |  |  |
| Asian | 165 | 1008 | 0 | 0 | 0 | 0 | 0 | 0 | 0 | 0 | 0 | 0 | 0 | 0 | 0 | 0 | 0 |
| Black | 165 | 851 | 0 | 0 | 0 | 0 | 0 | 0 | 0 | 0 | 0 | 0 | 0 | 0 | 0 | 0 | 0 |
| Hispanic | 157 | 782 | 0 | 0 | 0 | 0 | 0 | 0 | 0 | 0 | 0 | 0 | 0 | 0 | 0 | 0 | 0 |
| White | 160 | 785 | 0 | 0 | 0 | 0 | 0 | 0 | 0 | 0 | 0 | 0 | 0 | 0 | 0 | 0 | 0 |
| Whole | 244 | 2494 | 0 | 0 | 0 | 0 | 0 | 0 | 0 | 0 | 0 | 0 | 0 | 0 | 0 | 0 | 0 |
| **1107-Healthy Cohort (Doyle *et al.* 2018)** |  |  |  |  |  |  |  |  |  |  |  |  |  |  |  |  |  |
| UK-1107 | 461 | 6008 | 0 | 0 | 0 | 0 | 0 | 0 | 0 | 0 | 0 | 0 | 0 | 0 | 0 | 0 | 0 |

## Transparent Methods

### The HVM datasets

We used the “25-mixed cohort” dataset originally described by Ravel *et al*. (2013) for primary species dominance network (SDN) analysis. Briefly, Ravel *et al*. (2013) sequenced samples from vaginal communities collected daily for ten weeks from 25 women who were diagnosed as symptomatic BV (SBV: *n* = 15 women), asymptomatic BV (ABV: *n* = 6), or healthy (HEA: *n* = 4). In total, Ravel *et al*. (2013) sequenced 1,657 samples (median = 67 per woman) and obtained 8,757,681 high-quality sequenced reads of the V1–V3 hypervariable region of 16S-rRNA genes, with a median of 5,093 reads per sample.

We used three additional 16S-rRNA datasets of healthy women originally collected by Ravel *et al*. (2010), Gajer *et al*. (2012), and Doyle *et al*. (2018), respectively, to test the specificity of the 15 special trio motifs.. A total of 1535 healthy women were sampled in these three additional datasets. The dataset of “32-healthy cohort” (Gajer *et al*. 2012) consisted of the longitudinal sampling of 32 healthy women and has virtually the same data structure as the previously described “25-mixed cohort”. For this dataset, 32 SDNs were constructed to verify the findings from the primary dataset (25-mixed cohort). The “400-healthy cohort” dataset (Ravel *et al*. 2011) consisted of the cross-sectional samples of 397 healthy women from four ethnic groups (Asian, Black, Hispanic, and White). For this dataset, we built a SDN for each of the four ethnic groups as well as a fifth SDN of the combined groups. Finally, the dataset of “1107-healthy cohort” (Doyle *et al*. 2018) consisted of the cross-sectional samples of 1107 healthy, postpartum women. For this dataset, we built a single SDN for all individuals in the cohort.

### SDN analysis

The species dominance network (SDN) analysis we used was detailed in Ma & Ye (2017) and Ma & Ellison (2018, 2019). Briefly, by noting that *dominance* (in terms of numbers of 17individuals or total biomass) of a one or a few taxa in a multi-species assemblage results in an *uneven* distribution of all the taxa in the assemblage, measures of dominance can be equated to measures of unevenness, and by inversion, evenness of the assemblage. Whereas species diversity and species evenness have been applied routinely to multi-species assemblages or “communities”, we have shown that measures of dominance can be applied to both communities and individual species (Ma & Ellison 2018, 2019). The dominance metric that we derived provides a unified mathematical approach to measuring overall community dominance (≡ diversity), dominance by individual species (Ma & Ellison 2018, 2019), and relationships between dominance and community-wide stability and resilience. Additional insights were gained by developing techniques to detect *trio motifs*, *core/periphery* networks, and *high-salience skeleton* networks, and an approach to model network structure phenomenologically.

#### Trio motifs

A trio motif is the simplest *local* network of species interactions. In the present study, we detect the trios primarily consisting of BV-associated anaerobic bacteria (BVAB), and the structure and composition of trio motifs can have significant implications for the stability of the HVMC and BV etiology (Ma & Ye 2017, Ma & Ellison 2018, 2019). Unlike standard correlation analysis of networks, descriptions and analyses of trio motifs can take into accounts multiple properties of network “nodes” (taxa or operational taxonomic units [OTUs]) or “edges” (links between taxa or OTUs). These properties include node types—the most dominant OTU (MDO), the most abundant OTU (MAO), or the “hub” (most connections to other OTUs)—or edge types— positive or negative between OTUs.

#### Core/periphery and high-salience skeleton networks

Core/periphery (Csermely *et al*. 2013, Ma & Ellison 2019) and skeleton network analyses (Grady *et al*. 2012, Shekhtman *et al*. 2014, Ma & Ellison 2019) detect *global* (network- or community-wide) topological structures with significant implications for community stability or resilience. Core/periphery analyses focuses on nodes, identifying more stable and highly connected “core” OTUs and less stable and often loosely-connected “peripheral” OTUs. In contrast, skeleton analyses address the edges, including the interaction paths connecting OTUs that form the “backbone” or high-salience skeletons in a SDN. Because the sets of nodes and edges fully determine the topology of a network, our SDN analyses (Ma & Ellison 2018, 2019, Ma & Ye 2017) are complete—they address both nodes and edges (Fig. 1).

#### Analysis

We used the algorithms and computational procedures for describing and analyzing SDNs (Ma & Ellison 2018, 2019) to analyze the HVM datasets originally described by Ravel *et al*. (2013). Specifically, we used Spearman’s rank correlation coefficient (R) and for testing its significance, set α=0.001 after FDR (false discovery rate) adjustment. A SDN was considered valid if absolute |*R*| ≥ 0.5. In addition, we set α =0.05 for testing significance of data analyzed using a Kruskal-Wallis one-way analysis of variance (ANOVA).

## Data Availability

The data sources have been listed in the main manuscript.

**Abbreviations**
ABV: asymptomatic bacterial vaginosis
BV: bacterial vaginosis
BVAB: BV-associated anaerobic bacteria
CPN: core/periphery network
CST: community state type
DSR: diversity-stability relationship
HEA: healthy treatment
HSN: high-salience skeleton network
HVM: human vaginal microbiome
MAO: most abundant species or OTU
MDO: most dominant species or OTU
OTU: operational taxonomic unit
SBV: symptomatic BV
SDN: species dominance network

## Acknowledgements

This study received funding from the following sources: A National Natural Science Foundation (NSFC) Grant (No. 31970116); Cloud-Ridge Industry Technology Leader Grant; A China-US International Cooperation Project on Genomics/Metagenomics Big Data. We appreciate Wendy Li and Lianwei Li of the Chinese Academy of Sciences for their computational support.

## Author contributions

ZSM conceived and performed the analysis, interpreted the results and wrote the paper. AME participated in interpretation of the results and revised the paper. All authors approved the submission.

**Correspondence: should be addressed to ZSM**

## Data availability

The datasets are available in public domain and the source are given in Ravel *et al*. (2013) *Microbiome* 2013, Ravel *et al*. (2011) PNAS, Gajer *et al*. (2012) *Science Translational Medicine*, and Doyle *et al*. (2018) *Applied Environmental Microbiology*.

**Online Supplementary Information (OSI)** 3 OSI files (2 PDF & 1 Excel files) included.

**File-A:** “HVM OSI-File-A (Figures).pdf” contains: Fig S0 (Legend for the SDN graphs) and Fig S1 (the SDN graphs for 25 individuals in the 25-mixed cohort).

**File-B:** “HVM-OSI-File-B (Tables).pdf” contains: Table S1-1, Table S1-2, Table S2-1A, Table S2-1B, Table S3-1, Table S3-4B, Table S3-5B, Table S4-1, Table S4-3B, Table S4-4

**File-C:** “HVM-OSI-File-C (Excel Tables).xlsx” contains: Table S2-2, Table S2-3, Table S2-4, Table S2-5, Table S3-2A, Table S3-2B, Table S3-3, Table S3-4A, Table S3-5A, Table S4-2, Table S4-3A.

**List of Online Supplementary Tables** (included in “HVM-OSI-File-B (Tables).pdf” & “HVM-OSI-File-C (Excel Tables).xlsx”:

**(1) Basic properties of the SDNs**

**Table S1-1.**
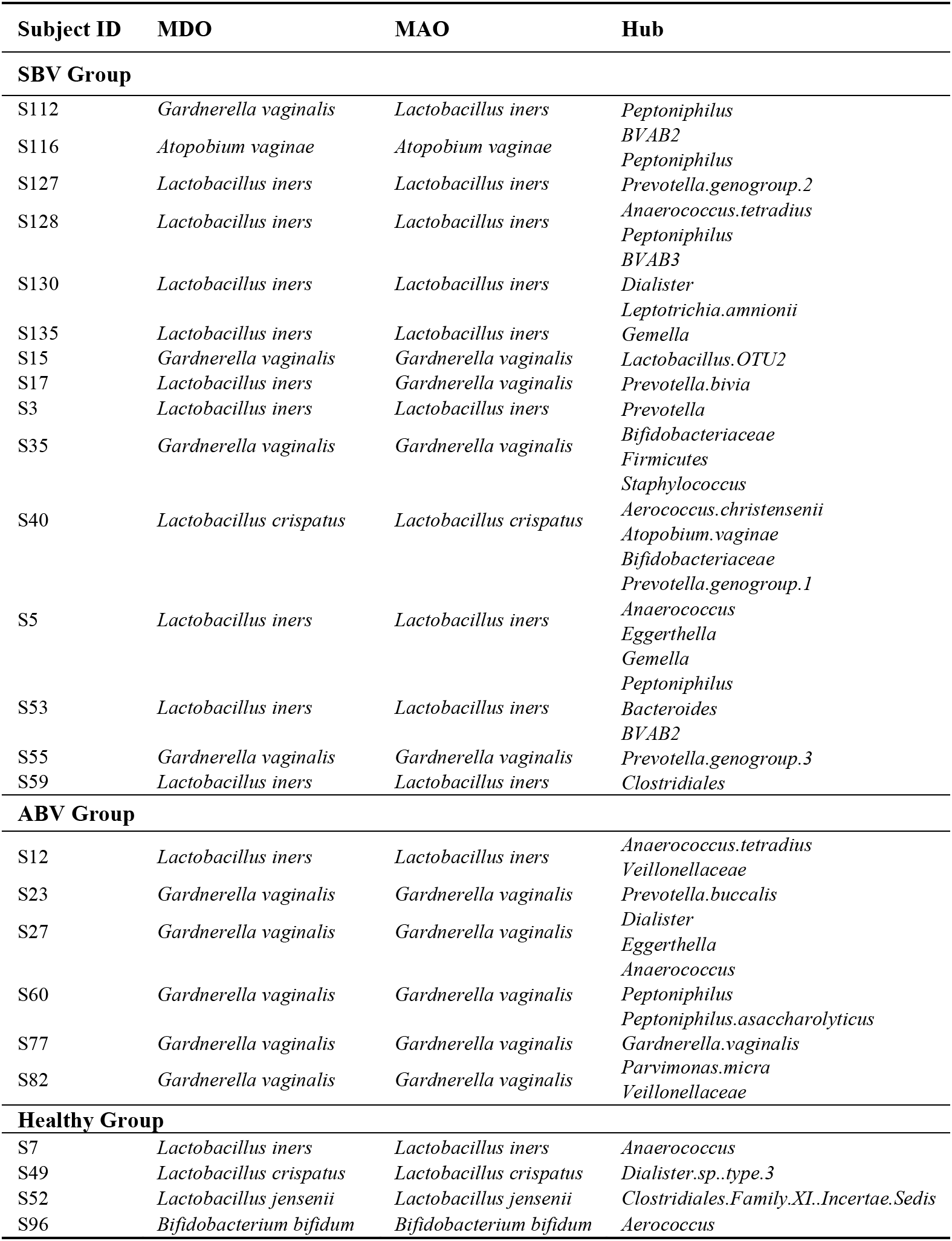
The MDO, MAO and hub in the SDNs of the 25-mixed cohort

**Table S1-2.** The basic network properties (parameters) of the SDNs for the 25-mixed cohort

| SDN ID | Num. of Nodes | Num. of Edges | Average Degree | Avg. Local Cluster Coefficient | Diameter | Average Path Length | Connected Components | Network Density | Network Modularity | Num. of Communities |
| --- | --- | --- | --- | --- | --- | --- | --- | --- | --- | --- |
| **SBV (Symptomatic BV) Group** |  |  |  |  |  |  |  |  |  |  |
| S112 | 68 | 626 | 18.412 | 0.806 | 4 | 1.617 | 4 | 0.275 | 0.155 | 19 |
| S116 | 50 | 155 | 6.200 | 0.626 | 7 | 2.691 | 6 | 0.127 | 0.315 | 8 |
| S127 | 71 | 287 | 8.085 | 0.710 | 6 | 2.772 | 6 | 0.115 | 0.263 | 12 |
| S128 | 61 | 242 | 7.934 | 0.592 | 10 | 3.664 | 3 | 0.132 | 0.300 | 7 |
| S130 | 42 | 342 | 16.286 | 0.869 | 5 | 1.467 | 4 | 0.397 | 0.057 | 5 |
| S135 | 78 | 422 | 10.821 | 0.779 | 5 | 1.728 | 7 | 0.141 | 0.347 | 8 |
| S15 | 76 | 284 | 7.474 | 0.568 | 7 | 3.044 | 2 | 0.100 | 0.454 | 5 |
| S17 | 46 | 203 | 8.826 | 0.669 | 5 | 1.793 | 3 | 0.196 | 0.156 | 3 |
| S3 | 12 | 19 | 3.167 | 0.907 | 2 | 1.136 | 3 | 0.288 | 0.499 | 3 |
| S35 | 83 | 492 | 11.855 | 0.697 | 7 | 2.819 | 2 | 0.145 | 0.513 | 6 |
| S40 | 54 | 185 | 6.852 | 0.771 | 8 | 3.208 | 3 | 0.129 | 0.370 | 10 |
| S5 | 61 | 178 | 5.836 | 0.489 | 7 | 3.081 | 4 | 0.097 | 0.344 | 11 |
| S53 | 57 | 342 | 12.000 | 0.791 | 6 | 1.861 | 7 | 0.214 | 0.117 | 8 |
| S55 | 74 | 261 | 7.054 | 0.473 | 7 | 2.884 | 4 | 0.097 | 0.290 | 11 |
| S59 | 65 | 328 | 10.092 | 0.604 | 7 | 2.593 | 4 | 0.158 | 0.204 | 23 |
| **Mean** | 59.867 | 291.067 | 9.393 | 0.690 | 6.200 | 2.424 | 4.133 | 0.174 | 0.292 | 9.267 |
| **Std. Err.** | 4.622 | 38.045 | 1.037 | 0.034 | 0.480 | 0.195 | 0.424 | 0.022 | 0.035 | 1.436 |
| **ABV (Asymptomatic BV) Group** |  |  |  |  |  |  |  |  |  |  |
| S12 | 90 | 609 | 13.533 | 0.739 | 10 | 3.887 | 3 | 0.152 | 0.388 | 8 |
| S23 | 79 | 203 | 5.139 | 0.637 | 11 | 4.012 | 4 | 0.066 | 0.637 | 10 |
| S27 | 63 | 130 | 4.127 | 0.548 | 10 | 3.697 | 4 | 0.067 | 0.660 | 8 |
| S60 | 84 | 159 | 3.786 | 0.491 | 13 | 4.152 | 6 | 0.046 | 0.656 | 13 |
| S77 | 66 | 175 | 5.303 | 0.689 | 10 | 3.953 | 5 | 0.082 | 0.629 | 12 |
| S82 | 46 | 109 | 4.739 | 0.694 | 6 | 2.625 | 4 | 0.105 | 0.620 | 6 |
| **Mean** | 71.333 | 230.833 | 6.105 | 0.633 | 10.000 | 3.721 | 4.333 | 0.086 | 0.598 | 9.500 |
| **Std. Err.** | 6.601 | 76.827 | 1.505 | 0.039 | 0.931 | 0.228 | 0.422 | 0.015 | 0.043 | 1.088 |
| **HEA Group (Healthy)** |  |  |  |  |  |  |  |  |  |  |
| S7 | 71 | 413 | 11.634 | 0.624 | 10 | 3.178 | 1 | 0.166 | 0.243 | 12 |
| S49 | 24 | 31 | 2.583 | 0.646 | 5 | 2.321 | 3 | 0.112 | 0.584 | 5 |
| S52 | 94 | 511 | 10.872 | 0.616 | 11 | 3.800 | 3 | 0.117 | 0.347 | 7 |
| S96 | 52 | 56 | 2.154 | 0.410 | 8 | 2.898 | 8 | 0.042 | 0.785 | 11 |
| **Mean** | 60.250 | 252.750 | 6.811 | 0.574 | 8.500 | 3.049 | 3.750 | 0.109 | 0.490 | 8.750 |
| **Std. Err.** | 14.823 | 122.562 | 2.571 | 0.055 | 1.323 | 0.307 | 1.493 | 0.025 | 0.122 | 1.652 |
| **The *p*-value of Kruskal-Wallis one-way analysis of variance (ANOVA)** |  |  |  |  |  |  |  |  |  |  |
| SBV vs. HEA | 0.960 | 0.841 | 0.424 | 0.162 | 0.092 | 0.134 | 0.472 | 0.194 | 0.110 | 0.840 |
| SBV vs. ABV | 0.150 | 0.110 | 0.036 | 0.350 | 0.006 | 0.004 | 0.545 | 0.010 | 0.001 | 0.456 |
| ABV vs. HEA | 0.670 | 0.670 | 0.670 | 0.286 | 0.377 | 0.088 | 0.274 | 0.394 | 0.286 | 0.592 |

**(2) Trio motifs and indicators of BV risk**

**Table S2-1A.**
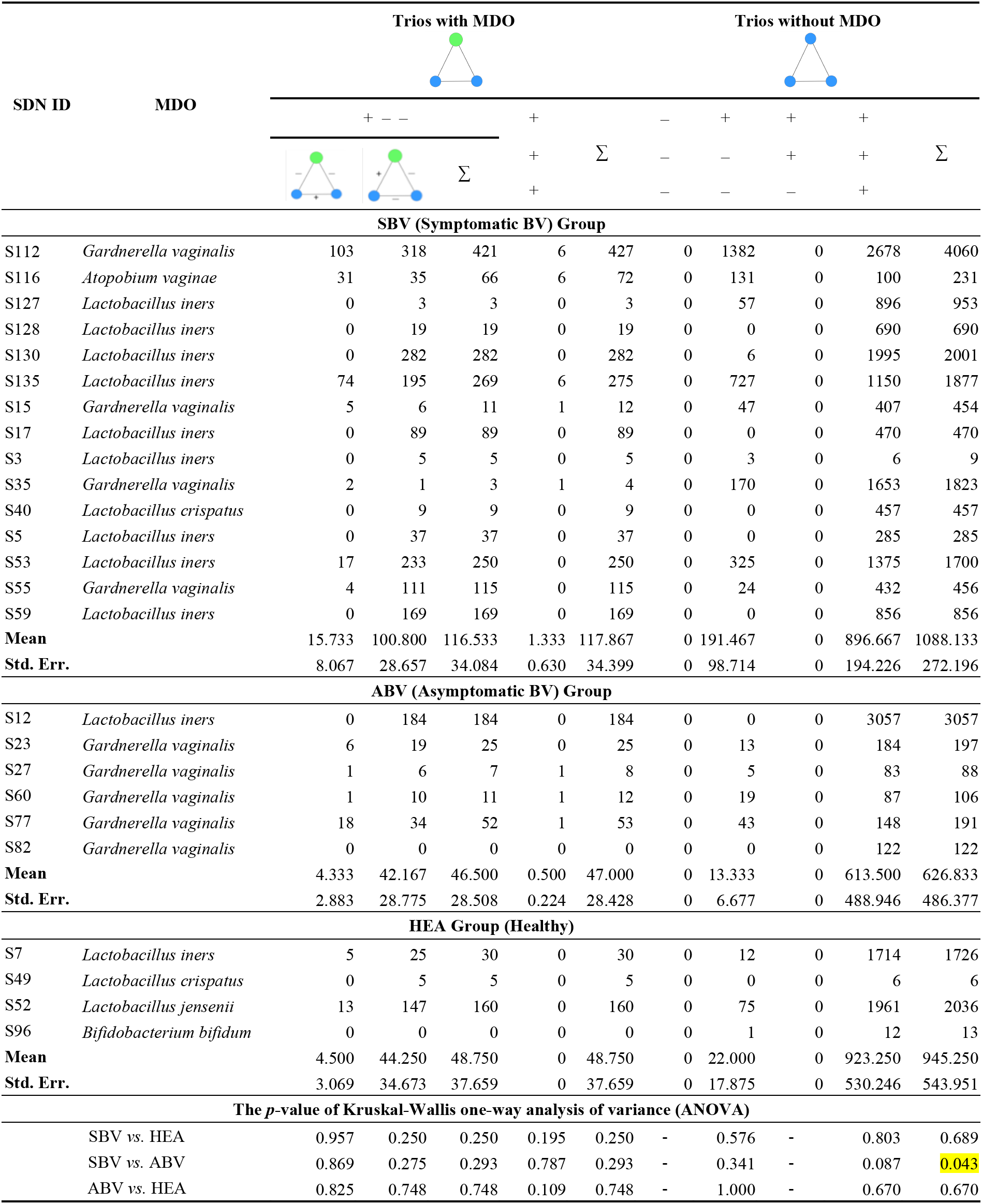
The occurrence of MDO-trios in the SDNs of the 25-mixed cohorts

**Table S2-1B.**
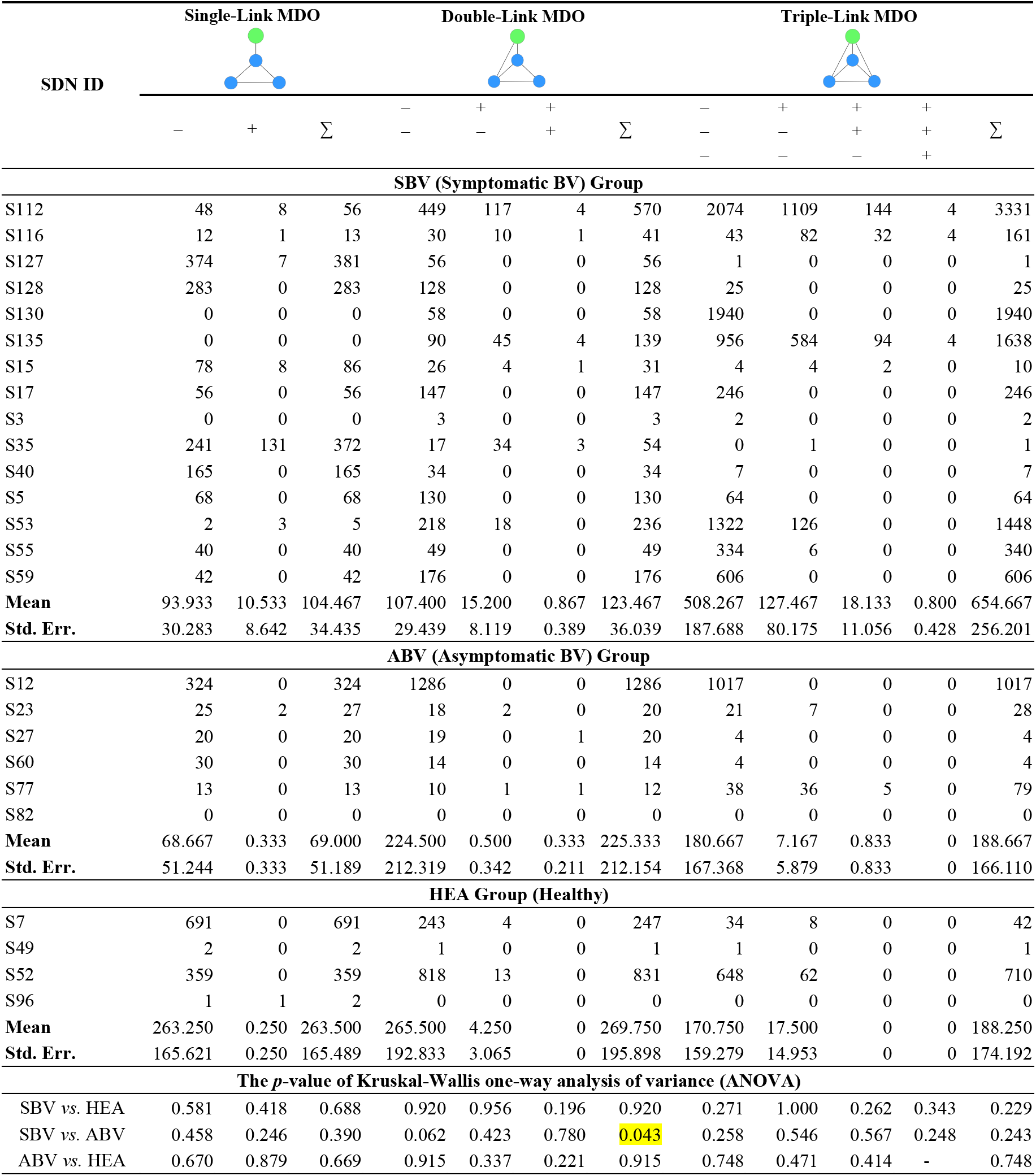
The occurrence of “MDO-trios with a handle” in the SDNs of the 25-mixed cohorts

**Table S2-2.** The 951 trios that occurred across 4 or more SDNs (subjects) of the 25-mixed cohort

*This table was listed in Excel File: HVM-OSI-File-C (Excel Tables).xlsx*

**Table S2-3.** The 71 trios occurred in 7 or more SDNs, and their relationships with MDO in the 25-mixed cohort. Note there were 26 BVAB-trios (all their nodes are BVAB), 12 trios occurred only in the SBV group, and 43 occurred only in SBV and ABV groups.

*This table was listed in Excel File: HVM-OSI-File-C (Excel Tables).xlsx*

**Table S2-4.** The 15 special trios, including the 12 trios occurred exclusively in the SBV group and 3 trios occurred in 50% or more BV subjects (BV=SBV+ABV)

*This table was listed in Excel File: HVM-OSI-File-C (Excel Tables).xlsx*

**Table S2-5.** The trios occurred in 50% or more subjects in each of three groups This table was listed in Excel File: HVM-OSI-File-C (Excel Tables).xlsx

**(3) Core/periphery species in the SDNs**

**Table S3-1.**
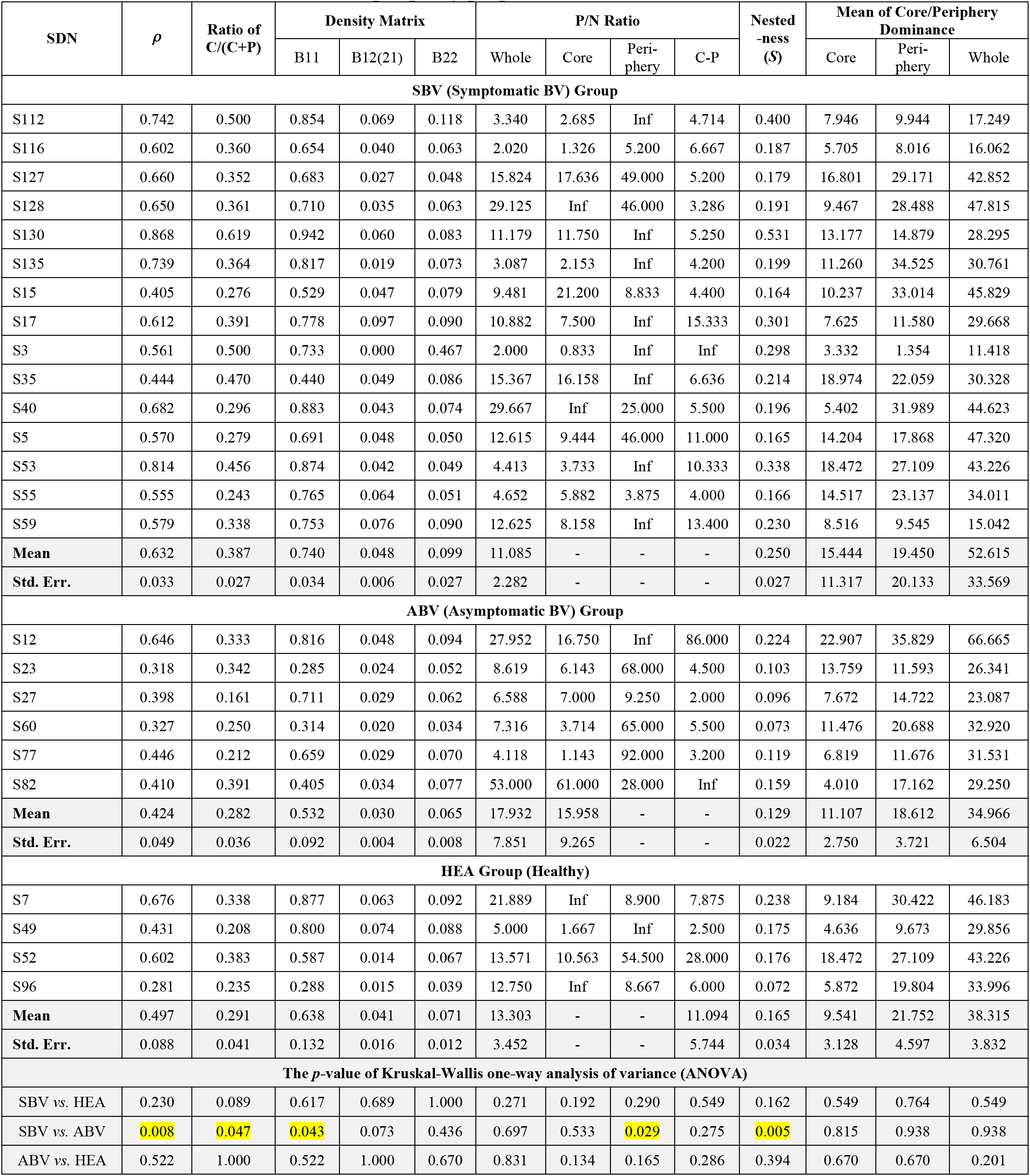
The core/periphery properties in the SDNs of the 25-mixed cohort

**Table S3-2A.** The OTU frequencies of the core and periphery nodes, respectively

*This table was listed in Excel File: HVM-OSI-File-C (Excel Tables).xlsx*

**Table S3-2B.** The OTU occurrence frequency in the core and periphery, respectively

*This table was listed in Excel File: HVM-OSI-File-C (Excel Tables).xlsx*

**Table S3-3.** The frequency of BVAB as core or periphery species

*This table was listed in Excel File: HVM-OSI-File-C (Excel Tables).xlsx*

**Table S3-4A.** Shared core (or periphery) OUT analysis between two SDNs from different groups

*This table was listed in Excel File: HVM-OSI-File-C (Excel Tables).xlsx*

**Table S3-4B.** Summary results of the shared core (periphery) species between each paired SDNs, from two of the SBV, ABV and HEA groups (summarized from Table S3-4A)

| Core |  |  |  |  |  |  |
| --- | --- | --- | --- | --- | --- | --- |
| Treatment |  | Observed<br>Shared OTUs | Algorithm A1 |  | Algorithm A2 |  |
|  |  |  | Expected<br>Shared OTUs | *p-value* | Expected<br>shared OTUs | *p-value* |
| HEA vs. ABV | Mean | 5.33 | 32.86 | <0.001 | 33.34 | <0.001 |
|  | Std. Err. | 0.99 | 2.20 | <0.001 | 2.13 | <0.001 |
| HEA vs. SBV | Mean | 5.18 | 34.92 | <0.001 | 35.50 | <0.001 |
|  | Std. Err. | 0.60 | 1.53 | <0.001 | 1.47 | <0.001 |
| ABV vs. SBV | Mean | 7.96 | 34.02 | <0.001 | 34.07 | <0.001 |
|  | Std. Err. | 0.40 | 0.94 | <0.001 | 0.91 | <0.001 |
| Periphery |  |  |  |  |  |  |
| Treatment |  | Observed<br>Shared OTUs | Algorithm A1 |  | Algorithm A2 |  |
|  |  |  | Expected<br>Shared OTUs | *p-value* | Expected<br>shared OTUs | *p-value* |
| HEA vs. ABV | Mean | 17.38 | 65.66 | <0.001 | 59.59 | <0.001 |
|  | Std. Err. | 1.74 | 2.61 | <0.001 | 2.18 | <0.001 |
| HEA vs. SBV | Mean | 12.77 | 53.01 | <0.001 | 50.85 | <0.001 |
|  | Std. Err. | 0.91 | 2.49 | <0.001 | 1.65 | <0.001 |
| ABV vs. SBV | Mean | 16.29 | 64.27 | <0.001 | 56.36 | <0.001 |
|  | Std. Err. | 0.74 | 1.78 | <0.001 | 1.19 | <0.001 |

**Table S3-5A.** The observed numbers of shared core (periphery) OTUs between each paired SDNs, from two of the SBV, ABV and HEA groups

*This table was listed in Excel File: HVM-OSI-File-C (Excel Tables).xlsx*

**Table S3-5B.**
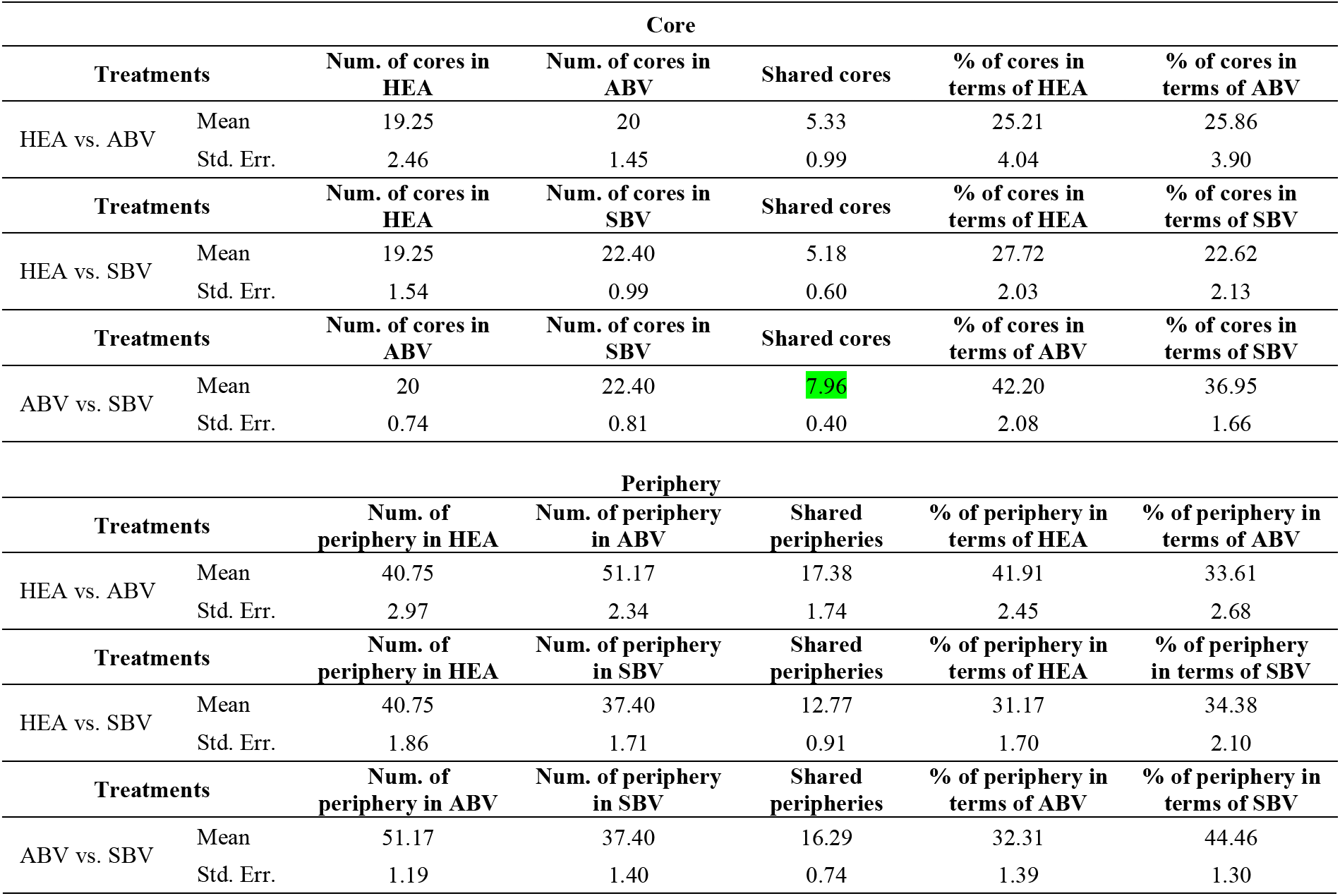
The *observed* number of shared core (periphery) OTUs among the SBV, ABV and HEA groups (Summarized from Table S3-5A)

| Core |  |  |  |  |  |  |
| --- | --- | --- | --- | --- | --- | --- |
| Treatments |  | Num. of cores in HEA | Num. of cores in ABV | Shared cores | % of cores in terms of HEA | % of cores in terms of ABV |
| HEA vs. ABV | Mean | 19.25 | 20 | 5.33 | 25.21 | 25.86 |
|  | Std. Err. | 2.46 | 1.45 | 0.99 | 4.04 | 3.90 |
| Treatments |  | Num. of cores in HEA | Num. of cores in SBV | Shared cores | % of cores in terms of HEA | % of cores in terms of SBV |
| HEA vs. SBV | Mean | 19.25 | 22.40 | 5.18 | 27.72 | 22.62 |
|  | Std. Err. | 1.54 | 0.99 | 0.60 | 2.03 | 2.13 |
| Treatments |  | Num. of cores in ABV | Num. of cores in SBV | Shared cores | % of cores in terms of ABV | % of cores in terms of SBV |
| ABV vs. SBV | Mean | 20 | 22.40 | 7.96 | 42.20 | 36.95 |
|  | Std. Err. | 0.74 | 0.81 | 0.40 | 2.08 | 1.66 |
| Periphery |  |  |  |  |  |  |
| Treatments |  | Num. of periphery in HEA | Num. of periphery in ABV | Shared peripheries | % of periphery in terms of HEA | % of periphery in terms of ABV |
| HEA vs. ABV | Mean | 40.75 | 51.17 | 17.38 | 41.91 | 33.61 |
|  | Std. Err. | 2.97 | 2.34 | 1.74 | 2.45 | 2.68 |
| Treatments |  | Num. of periphery in HEA | Num. of periphery in SBV | Shared peripheries | % of periphery in terms of HEA | % of periphery in terms of SBV |
| HEA vs. SBV | Mean | 40.75 | 37.40 | 12.77 | 31.17 | 34.38 |
|  | Std. Err. | 1.86 | 1.71 | 0.91 | 1.70 | 2.10 |
| Treatments |  | Num. of periphery in ABV | Num. of periphery in SBV | Shared peripheries | % of periphery in terms of ABV | % of periphery in terms of SBV |
| ABV vs. SBV | Mean | 51.17 | 37.40 | 16.29 | 32.31 | 44.46 |
|  | Std. Err. | 1.19 | 1.40 | 0.74 | 1.39 | 1.30 |

**4. High-salience skeletons in the SDNs**

**Table S4-1.**
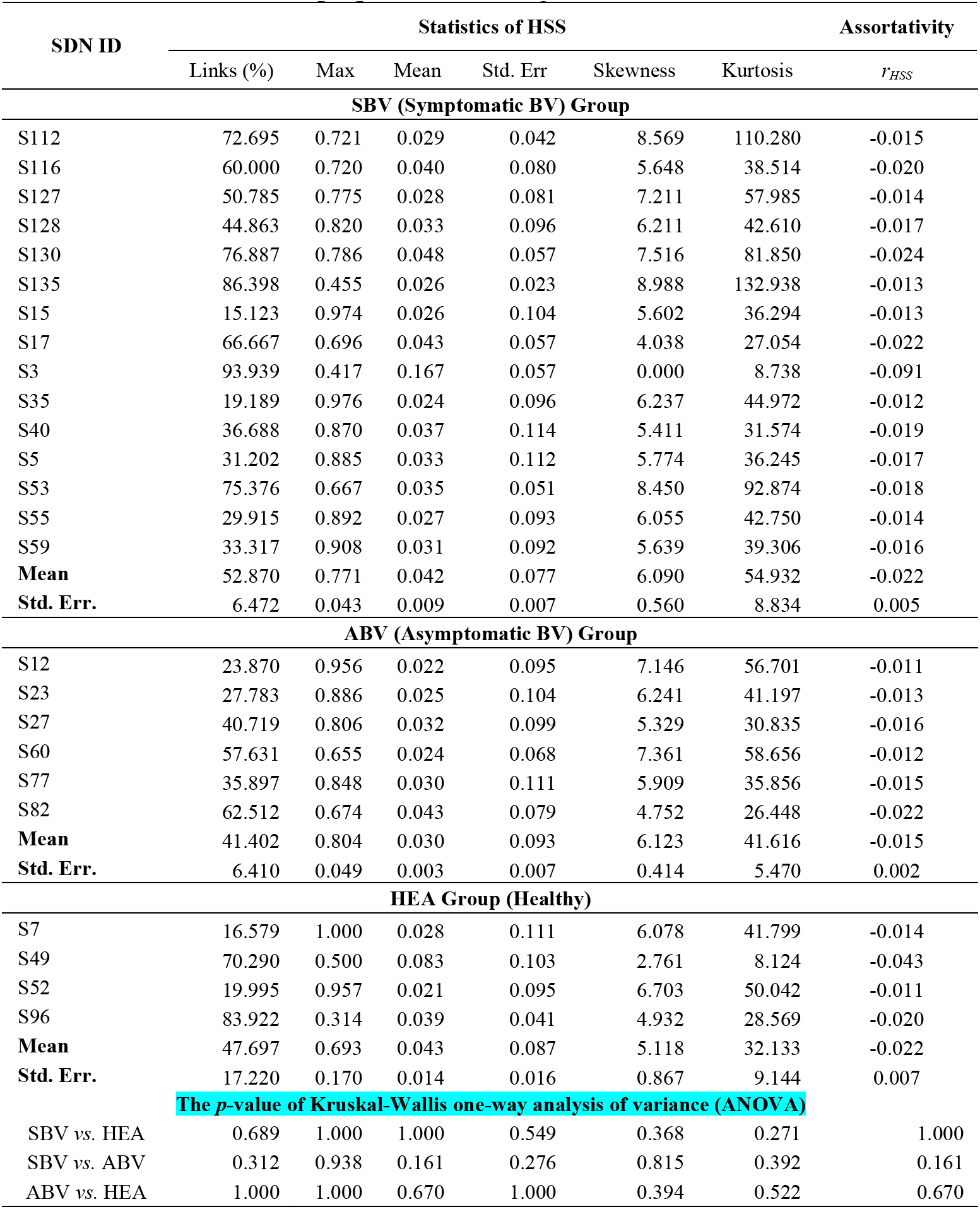
Statistical properties of the high-salience skeletons in the SDNs

**Table S4-2.** The list of high-salience skeletons that are incident on the two special OTUs they connect *(e.g*., MDO, MAO, hub, CST-indicator species or BVAB) with salience value ≥ 0.5 in the SDNs of the 25-mixed cohort

*This table was listed in Excel File: HVM-OSI-File-C (Excel Tables).xlsx*

**Table S4-3A.** The *observed* number of shared skeletons between each paired SDNs, from two of the HEA, ABV and SBV groups

*This table was listed in Excel File: HVM-OSI-File-C (Excel Tables).xlsx*

**Table S4-3B.**
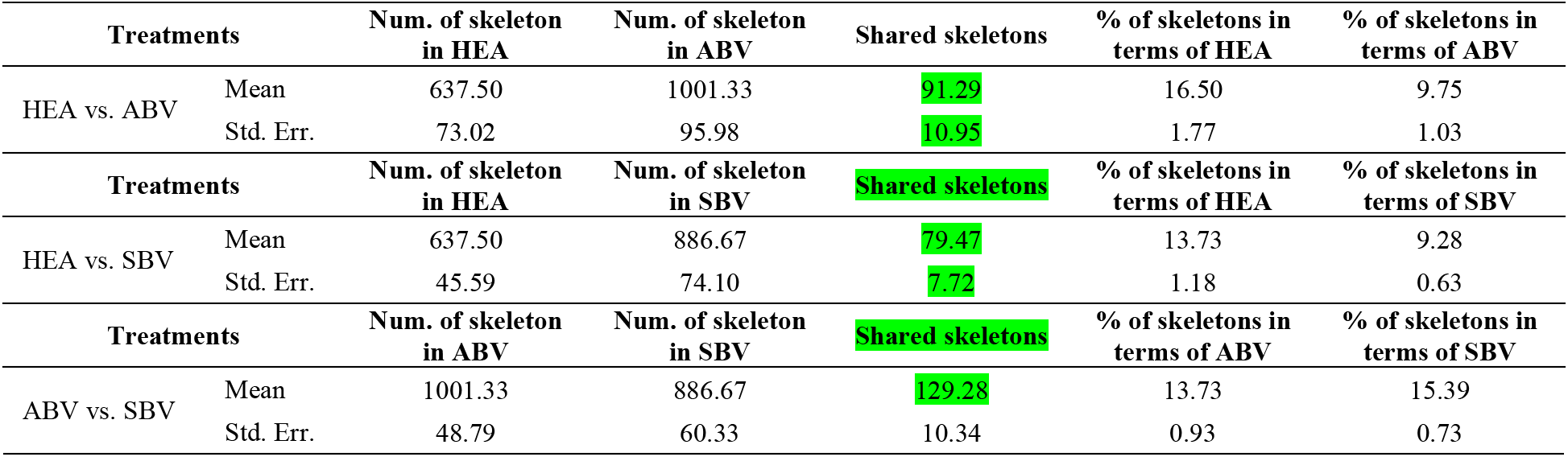
The *observed* number of shared skeletons among HEA, ABV and SBV groups (Summarized from Table S4-3A)

**Table S4-4.** The differential significant test for the salience values between the special trio-motifs and the whole SDN

| SDN ID | Special trio ID | Special Trio |  | Whole Network |  | *p*-value of Wilcoxon test |
| --- | --- | --- | --- | --- | --- | --- |
|  |  | Mean | Std. Err | Mean | Std. Err |  |
| SBV (Symptomatic BV) Group |  |  |  |  |  |  |
| s112 | #3 #34 #40 #50 #59 | 0.047 | 0.014 | 0.029 | 0.042 | 0.000 |
| s116 | #1 #34 #40 | 0.248 | 0.288 | 0.040 | 0.080 | 0.000 |
| s127 | #1 #3 #5 #35 | 0.097 | 0.110 | 0.028 | 0.081 | 0.000 |
| s128 | #1 #3 #5 #17 #22<br>#34 #35 #39 #40<br>#47 #48 #58 #59 | 0.109 | 0.101 | 0.033 | 0.096 | 0.000 |
| s130 | #1 #5 #17 #22 #39<br>#40 #47 #48 #50<br>#58 #65 | 0.091 | 0.095 | 0.048 | 0.057 | 0.000 |
| s135 | #1 #3 #22 #34 #35<br>#39 #47 #48 #50<br>#58 #59 #65 | 0.038 | 0.019 | 0.026 | 0.023 | 0.000 |
| s15 | #1 #3 #17 #22 #50<br>#65 | 0.317 | 0.222 | 0.026 | 0.104 | 0.000 |
| s17 | #1 #17 #22 #35 #40<br>#50 #65 | 0.128 | 0.101 | 0.043 | 0.057 | 0.000 |
| s3 | #5 | 0.194 | 0.048 | 0.167 | 0.057 | 0.000 |
| s35 | #1 #5 #58 #59 #65 | 0.245 | 0.284 | 0.024 | 0.096 | 0.000 |
| s40 | #1 #5 #17 #22 #34<br>#35 #39 #40 #47<br>#48 #58 #59 | 0.111 | 0.109 | 0.037 | 0.114 | 0.000 |
| s5 | #1 #3 #17 #34 #35<br>#39 #47 #48 | 0.106 | 0.066 | 0.033 | 0.112 | 0.000 |
| s53 | #17 #22 #34 #35<br>#39 #40 #47 #48<br>#50 #58 #59 #65 | 0.050 | 0.016 | 0.035 | 0.051 | 0.000 |
| s59 | #1 #5 #17 #22 #39<br>#47 #48 #50 #58<br>#59 #65 | 0.095 | 0.071 | 0.031 | 0.092 | 0.000 |
| ABV (Asymptomatic BV) Group |  |  |  |  |  |  |
| s12 | #3 | 0.037 | 0.017 | 0.022 | 0.095 | 0.003 |
| s27 | #1 #3 #5 | 0.301 | 0.149 | 0.032 | 0.099 | 0.000 |
| s60 | #1 #3 #5 | 0.169 | 0.232 | 0.024 | 0.068 | 0.000 |
| s77 | #1 #3 #5 | 0.135 | 0.199 | 0.030 | 0.111 | 0.000 |
| s82 | #1 | 0.065 | 0.022 | 0.043 | 0.079 | 0.110 |

Online Supplementary Figures for: Ma & Ellison (2020) *“Risks and etiology of bacterial vaginosis revealed by species dominance network analysis”*

**Figure S0.**
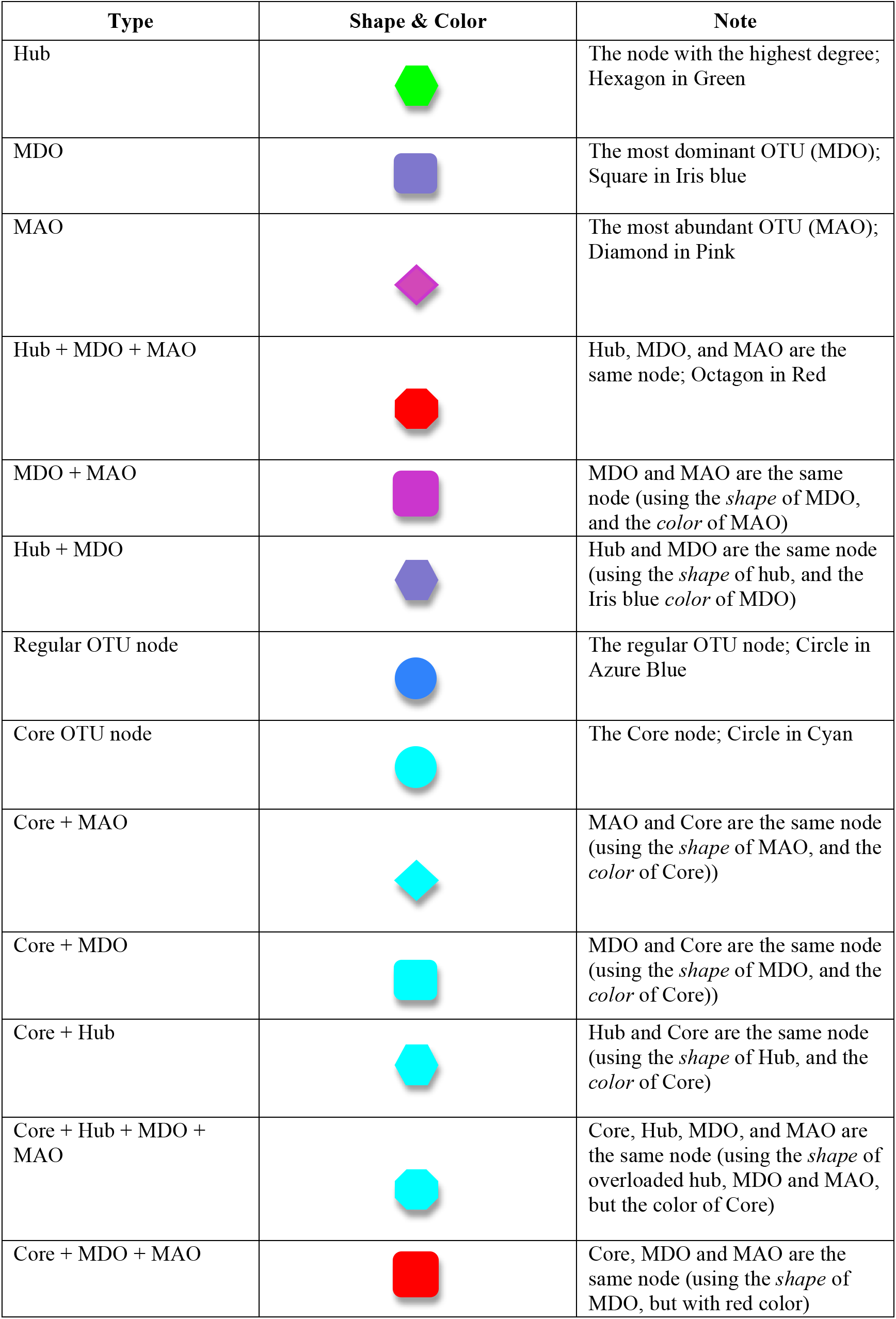
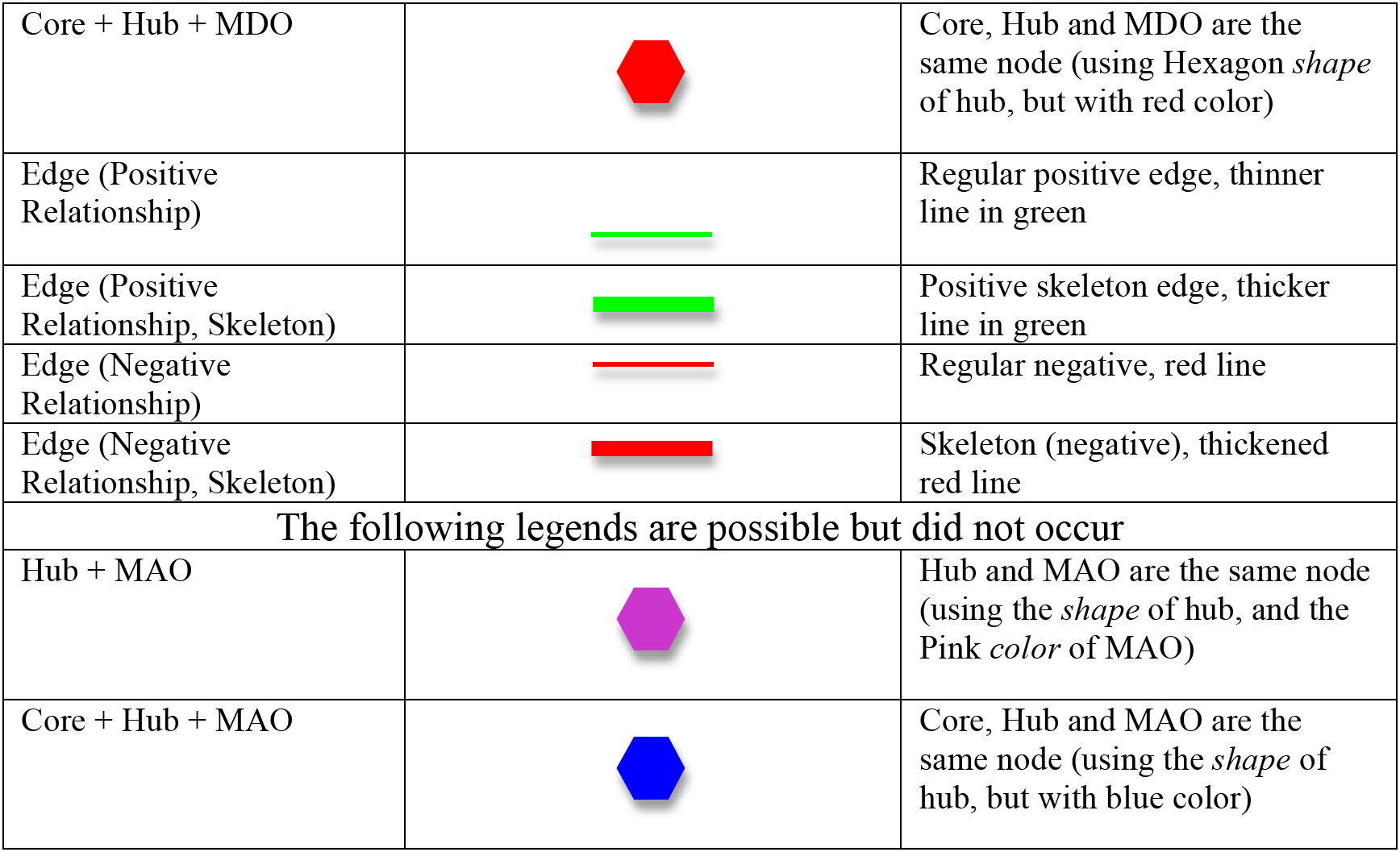
The legends of network symbols for the SDNs of the 25-mixed cohort

**Fig S1-1.**
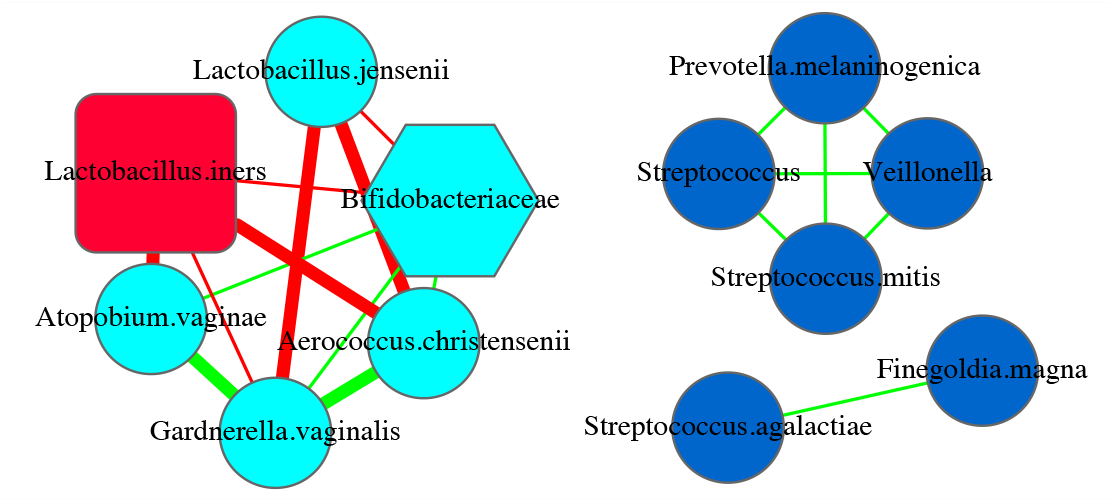
SDN network graph for Subject #s3 of the 25-mixed cohort

**Fig S1-2.**
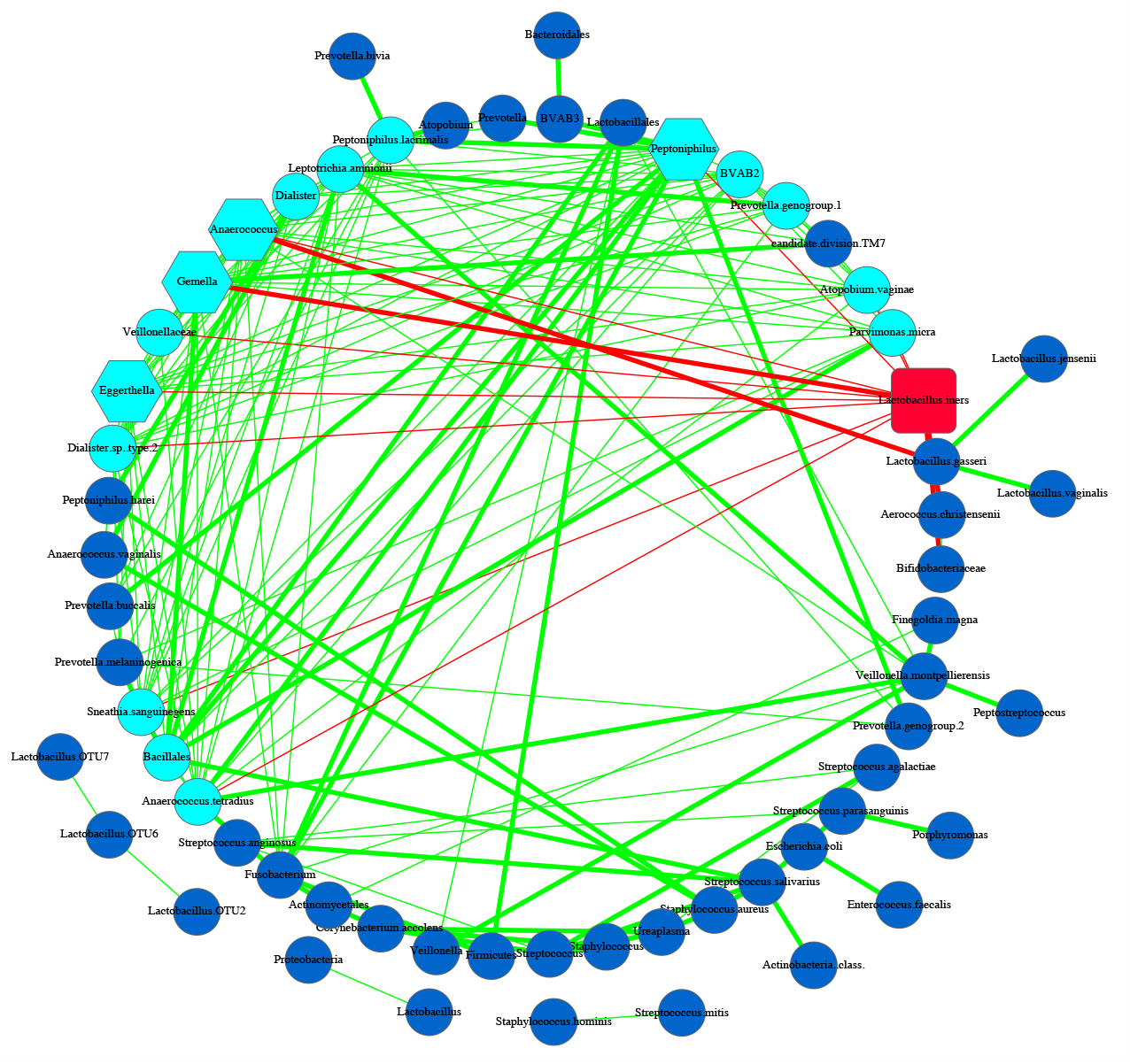
SDN network graph for Subject #s5 of the 25-mixed cohort

**Fig S1-3.**
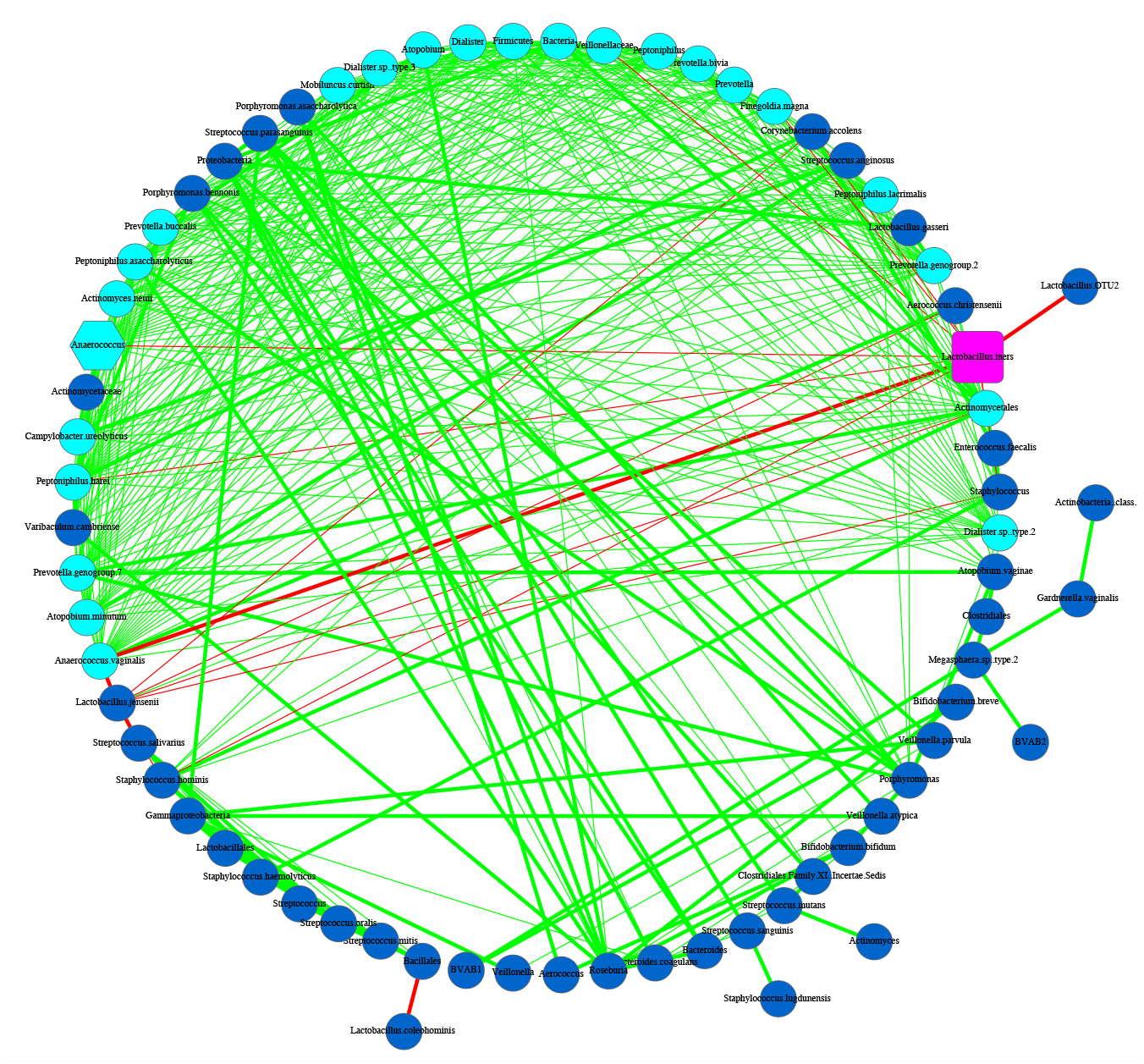
SDN network graph for Subject #s7 of the 25-mixed cohort

**Fig S1-4.**
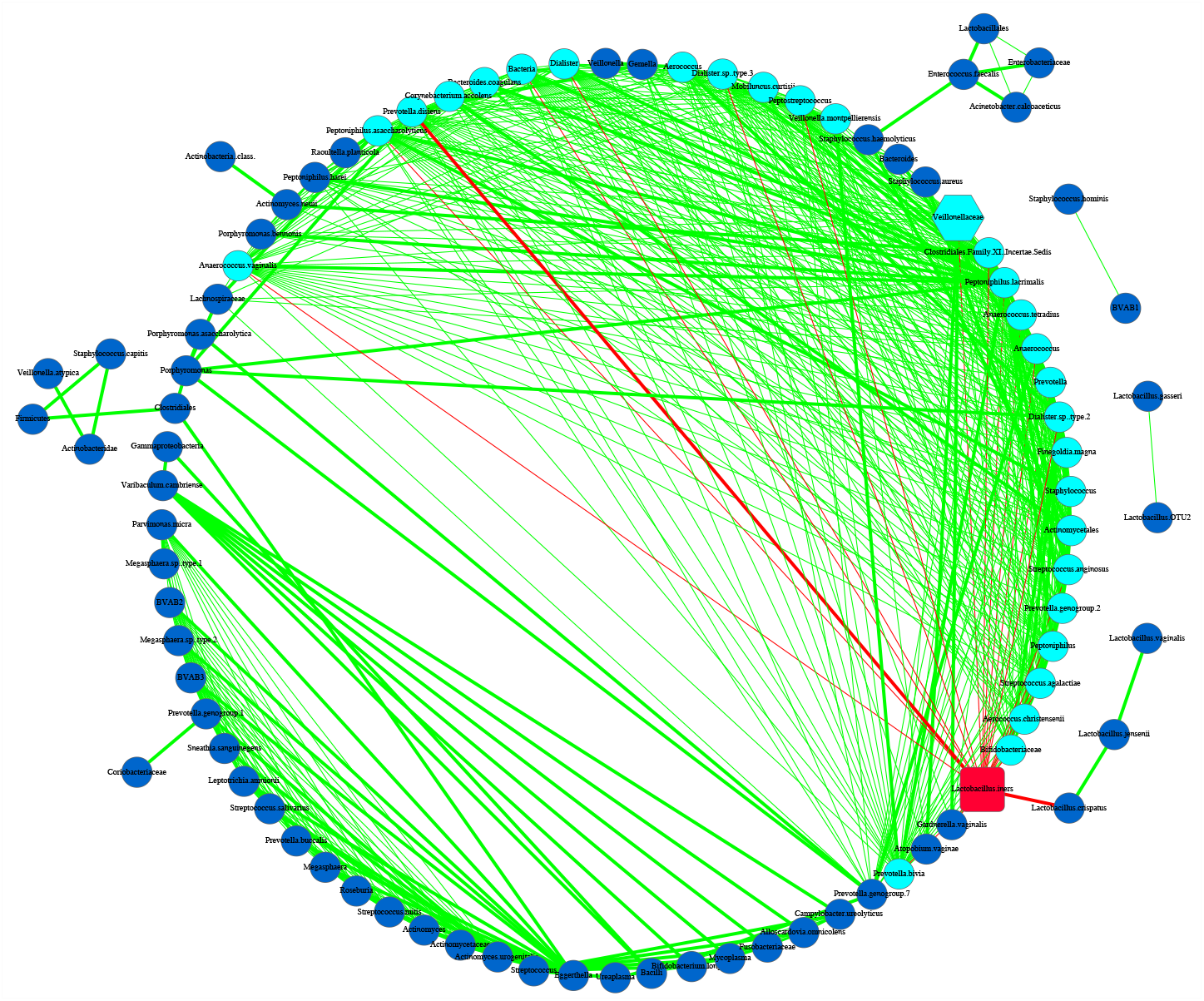
SDN network graph for Subject #s12 of the 25-mixed cohort

**Fig S1-5.**
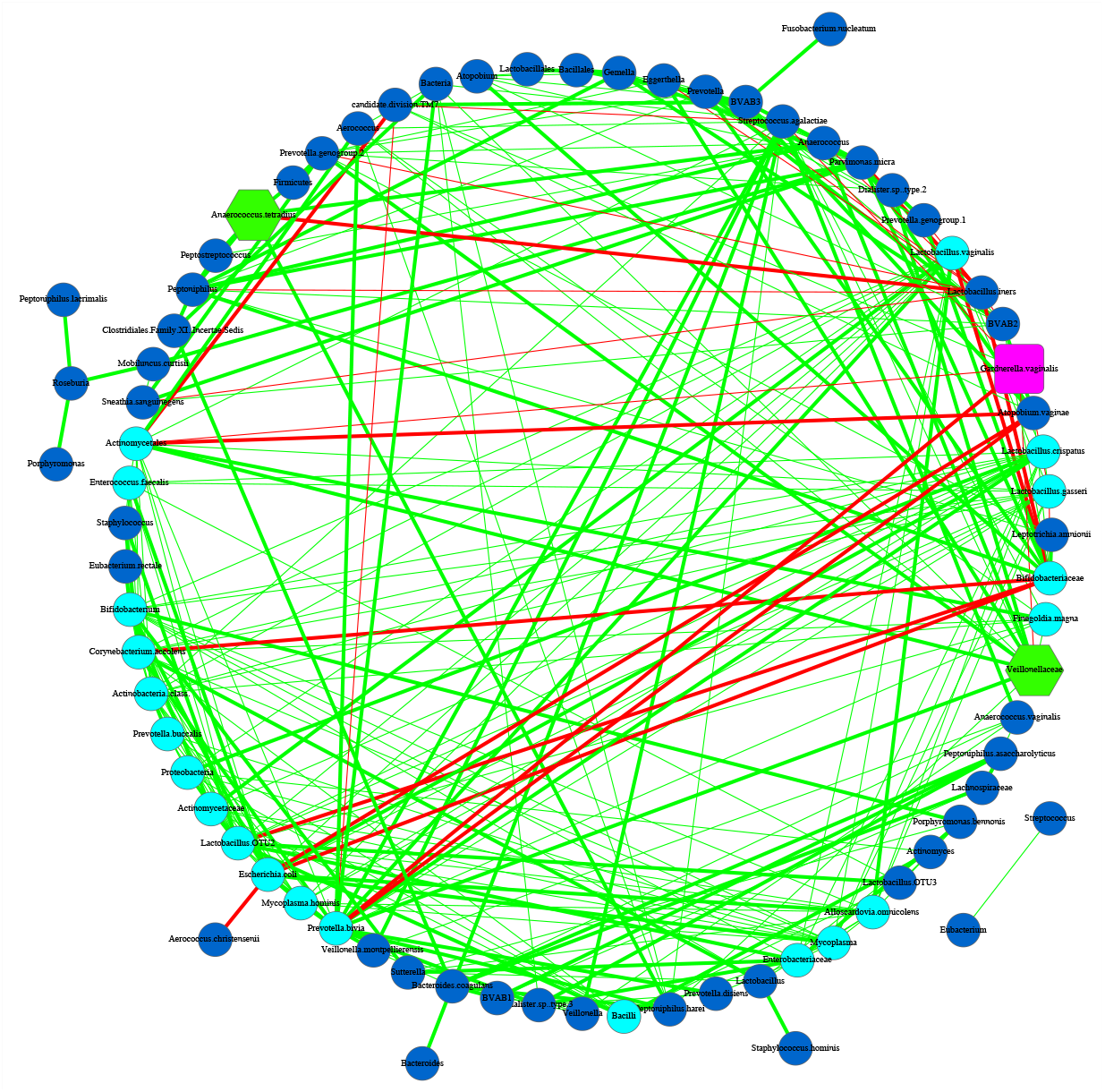
SDN network graph for Subject #s15 of the 25-mixed cohort

**Fig S1-6.**
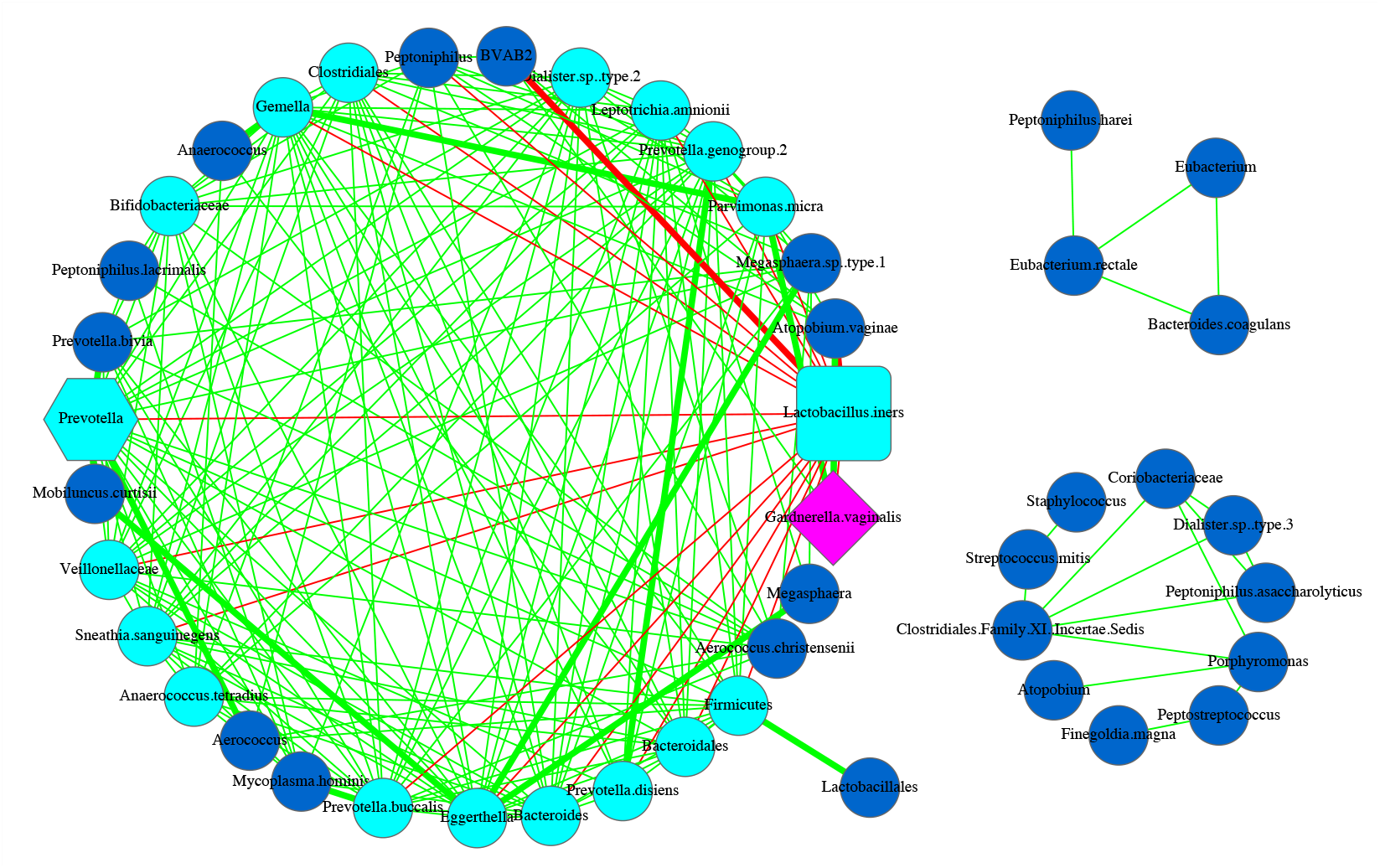
SDN network graph for Subject #s17 of the 25-mixed cohort

**Fig S1-7.**
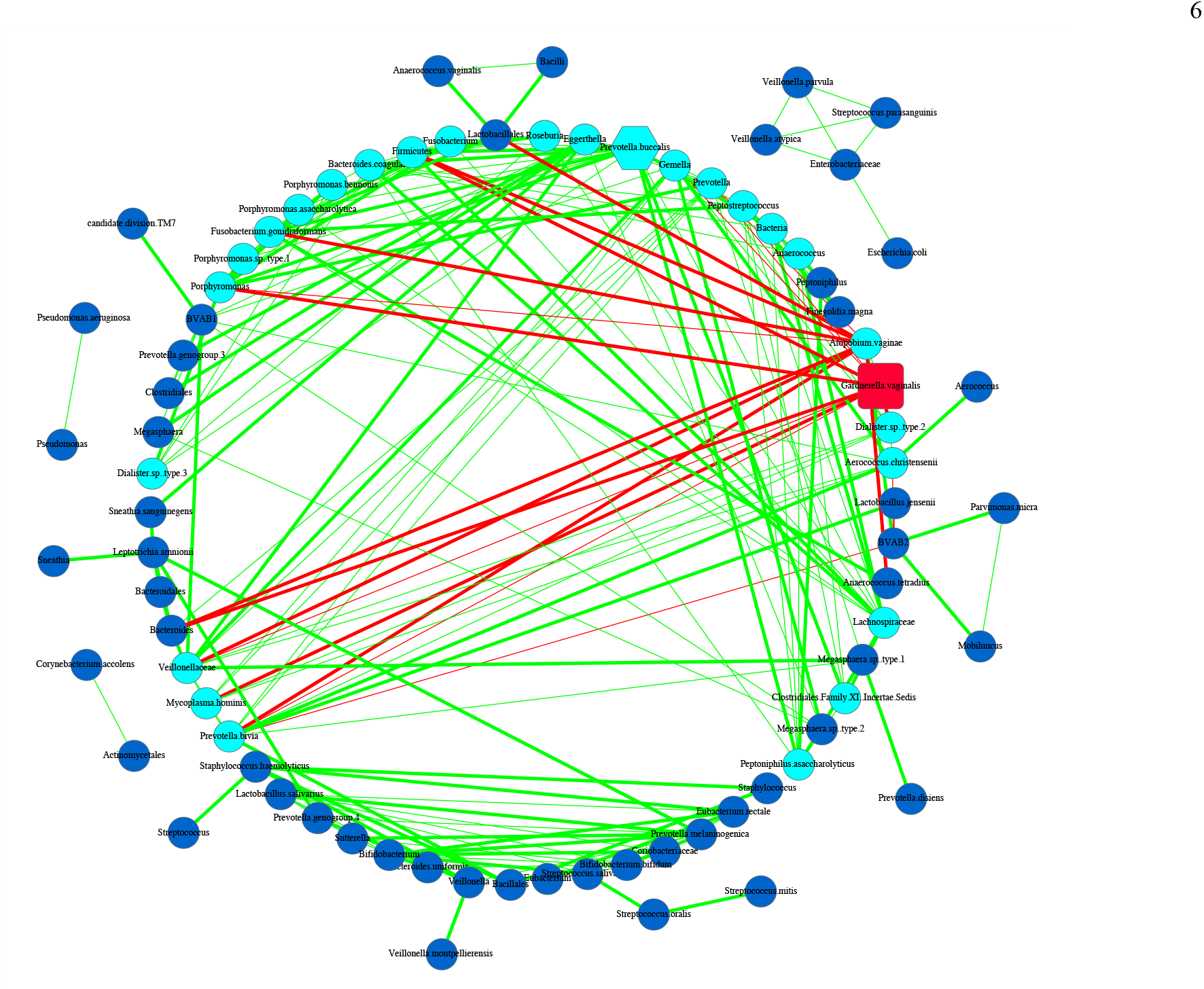
SDN network graph for Subject #s23 of the 25-mixed cohort

**Fig S1-8.**
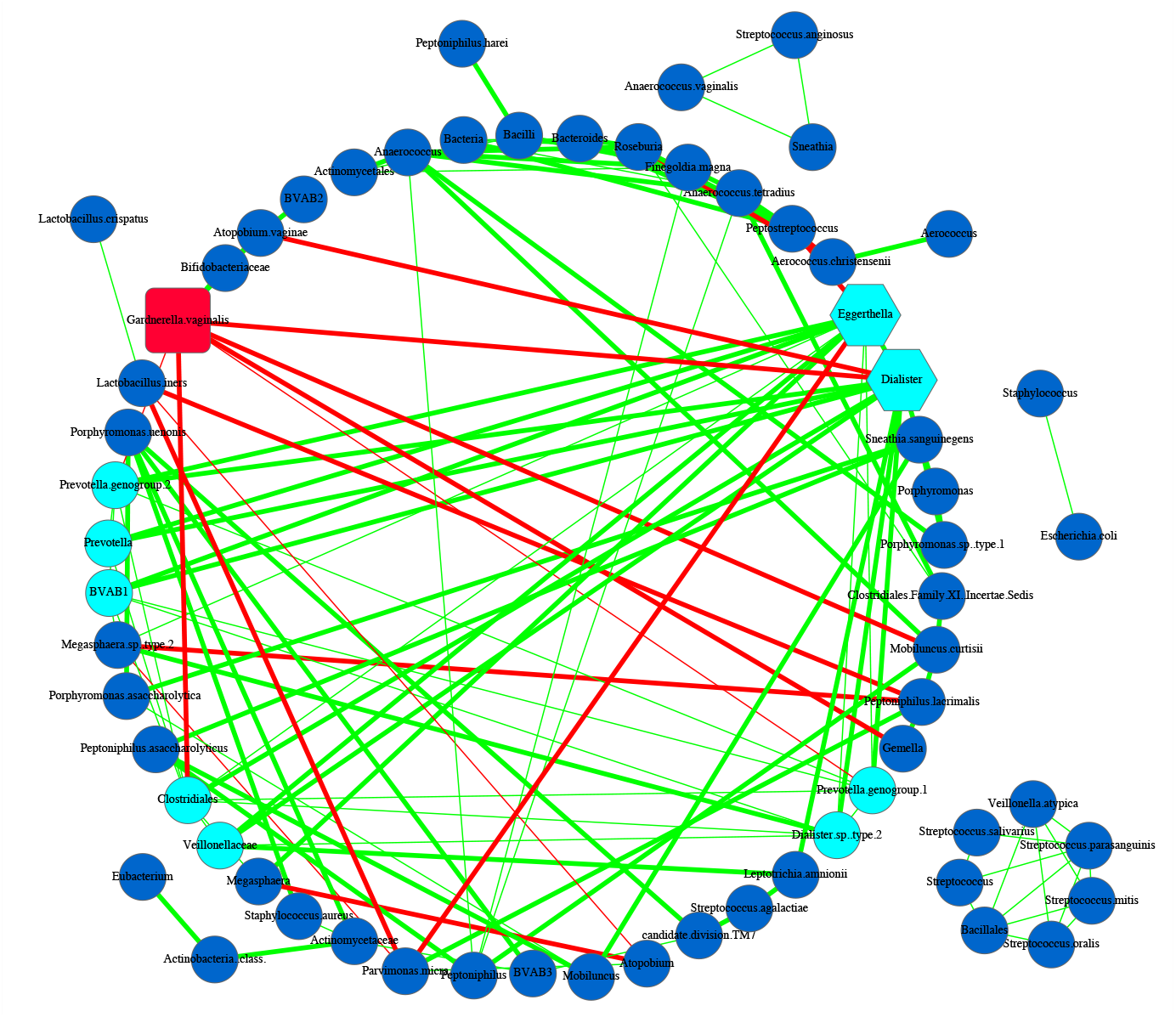
SDN network graph for Subject #s27 of the 25-mixed cohort

**Fig S1-9.**
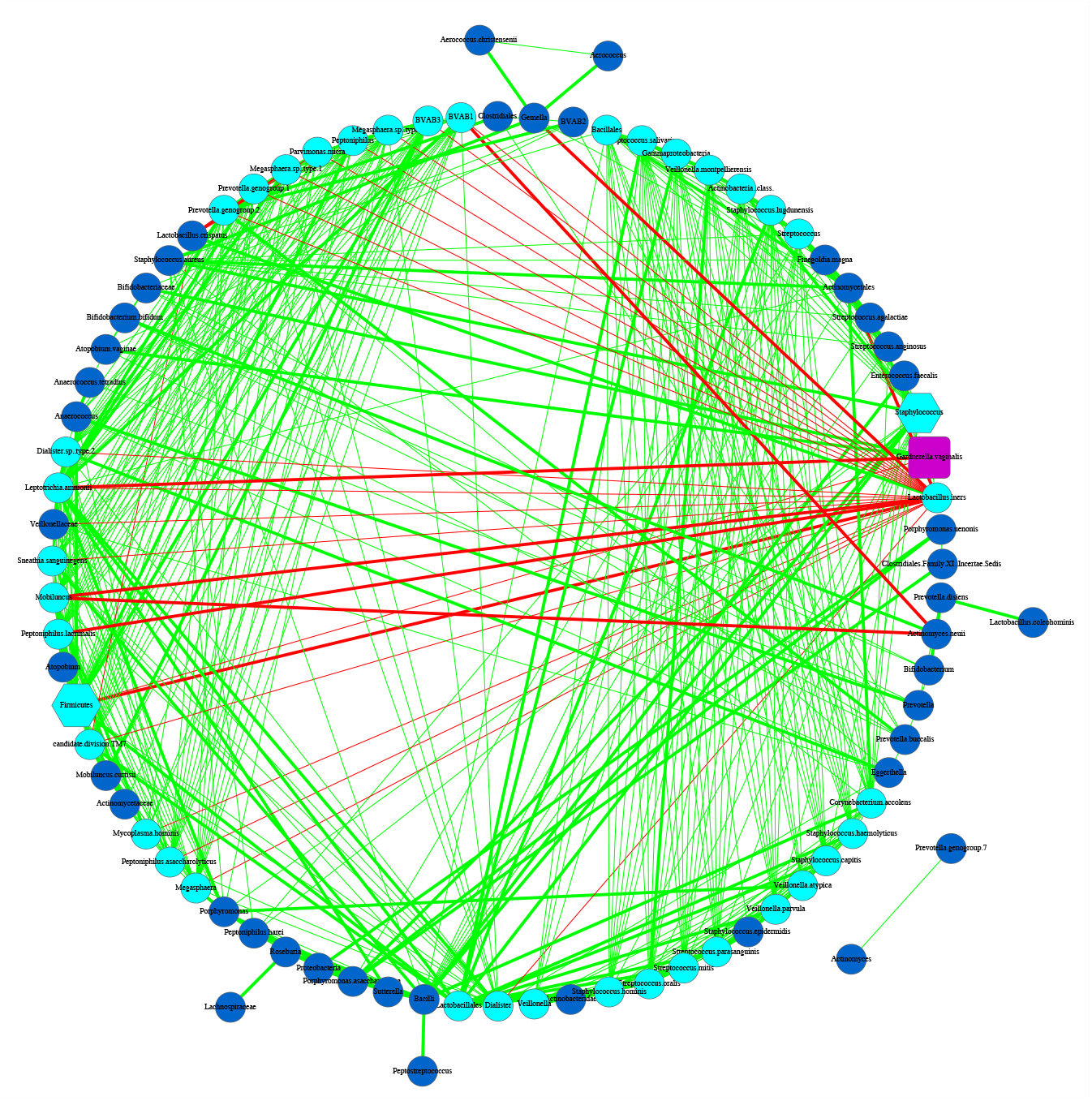
SDN network graph for Subject #s35 of the 25-mixed cohort

**Fig S1-10.**
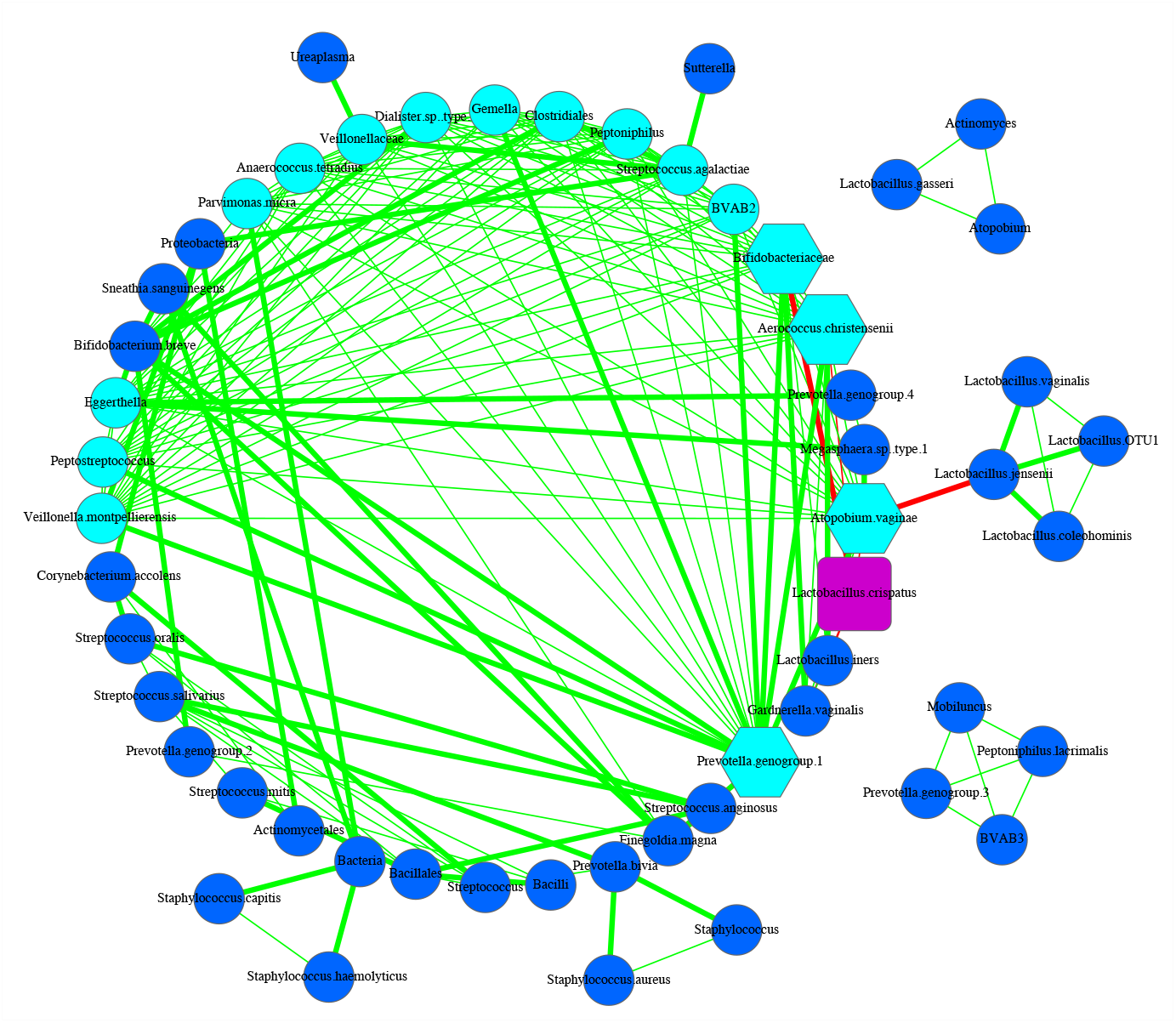
SDN network graph for Subject #s40 of the 25-mixed cohort

**Fig S1-11.**
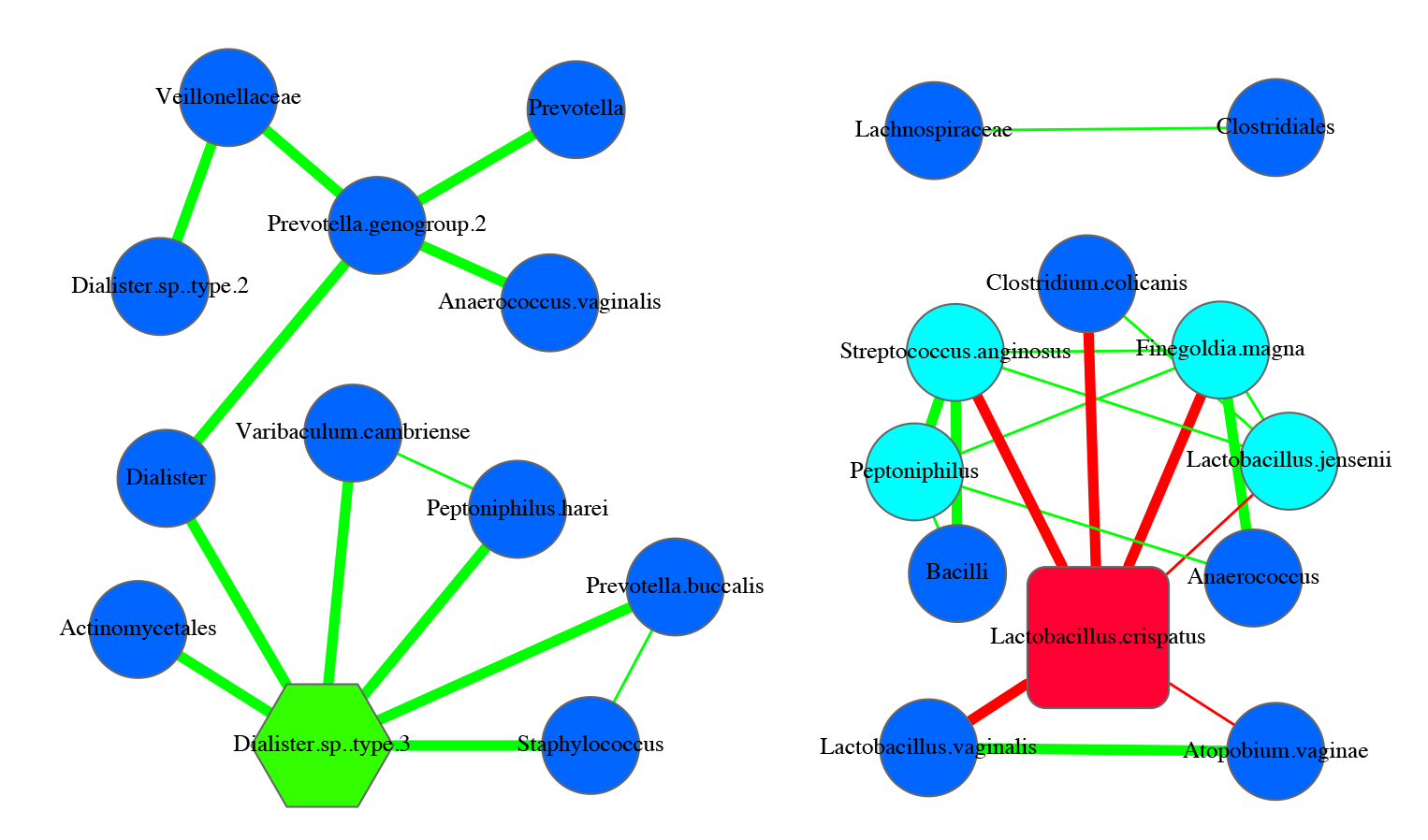
SDN network graph for Subject #s49 of the 25-mixed cohort

**Fig S1-12.**
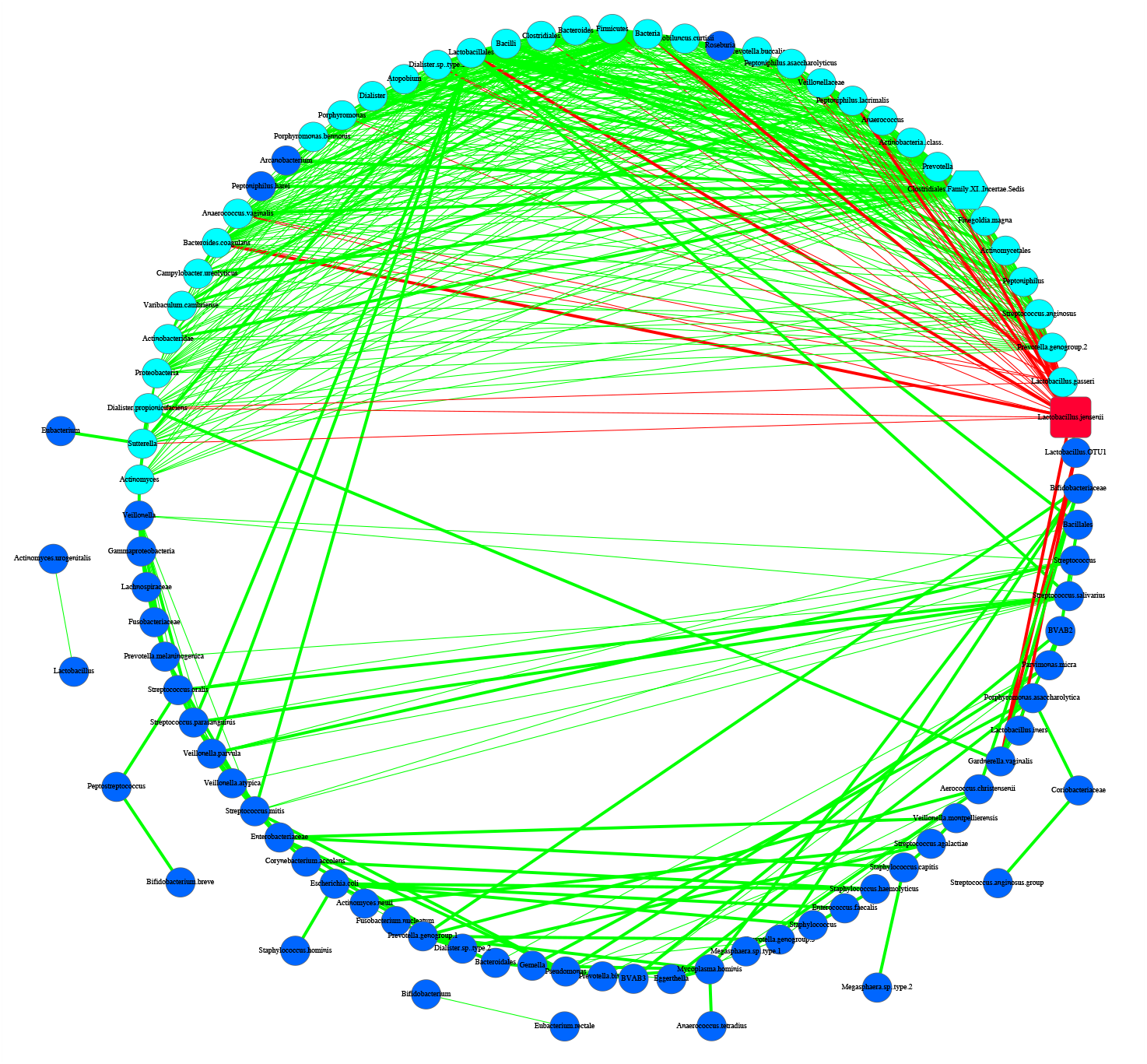
SDN network graph for Subject #s52 of the 25-mixed cohort

**Fig S1-13.**
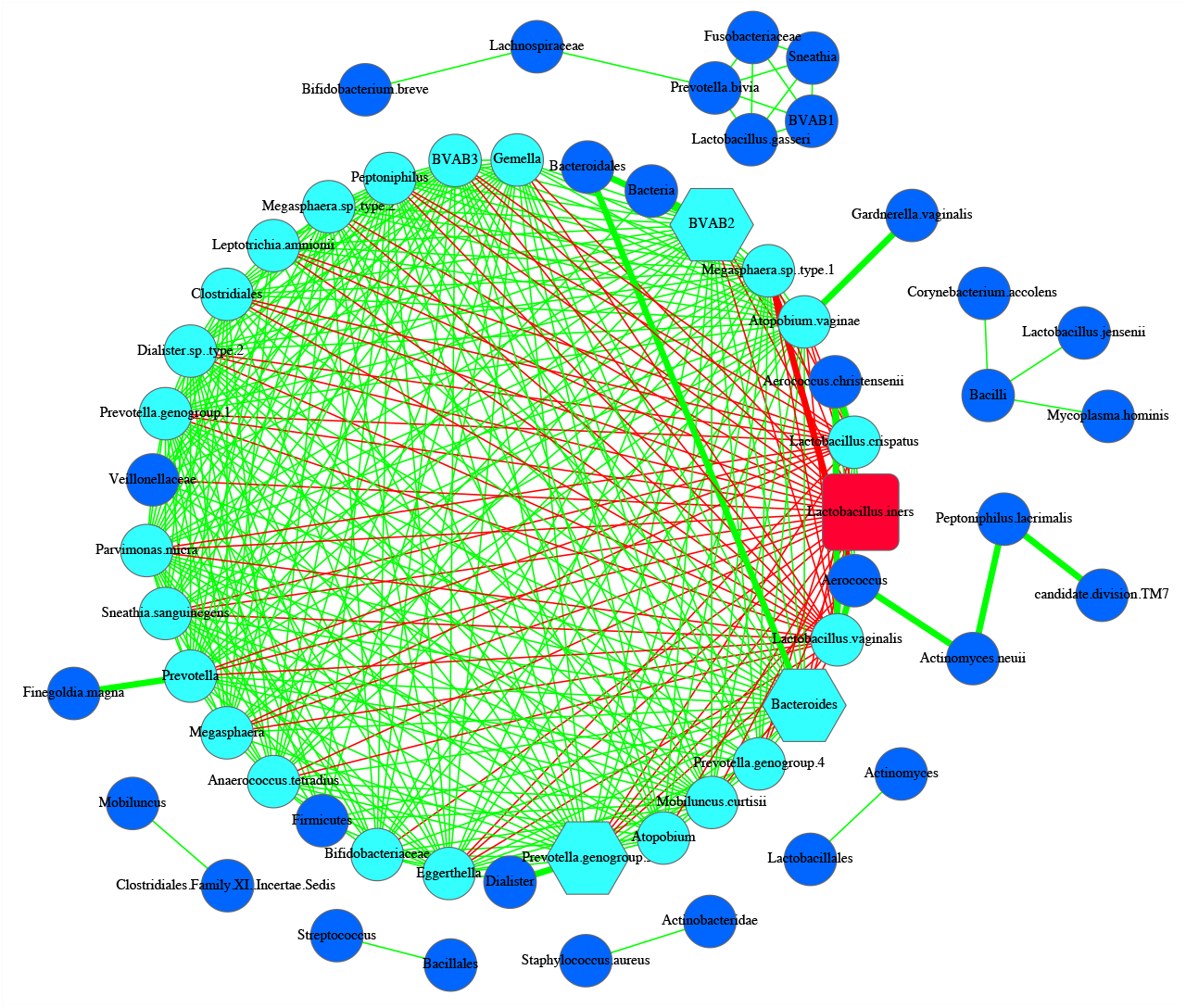
SDN network graph for Subject #s53 of the 25-mixed cohort

**Fig S1-14.**
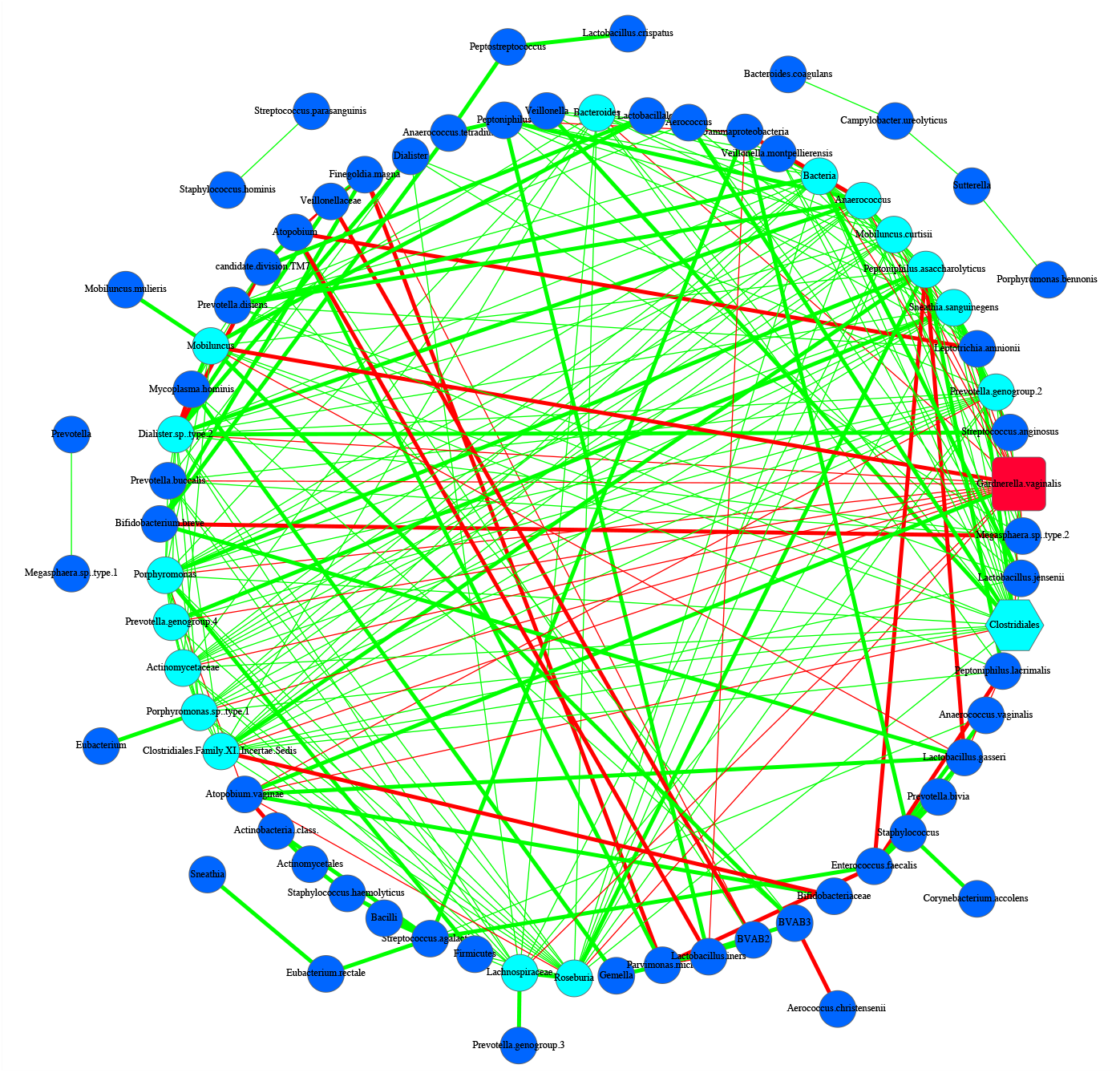
SDN network graph for Subject #s55 of the 25-mixed cohort

**Fig S1-15.**
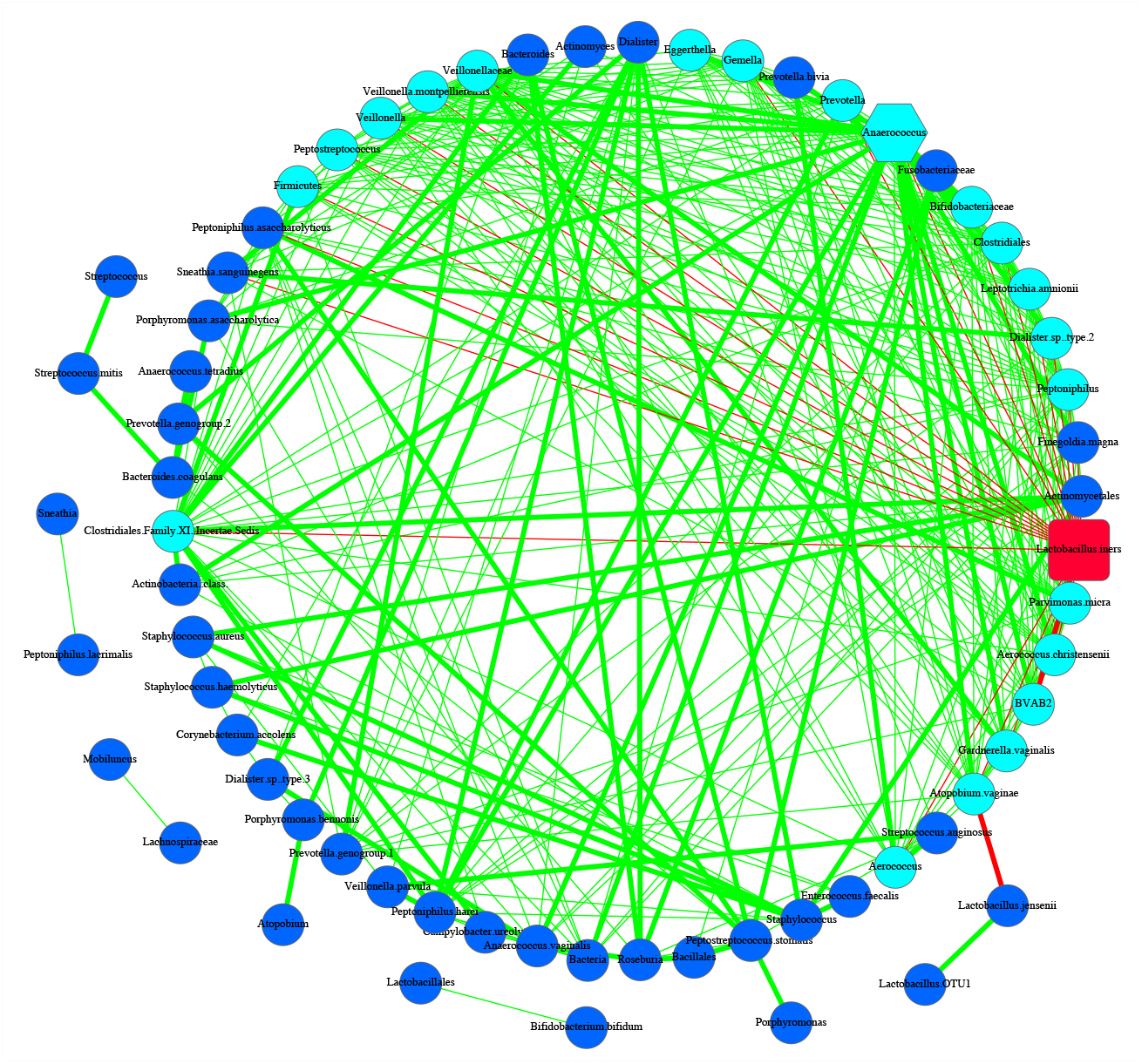
SDN network graph for Subject #s59 of the 25-mixed cohort

**Fig S1-16.**
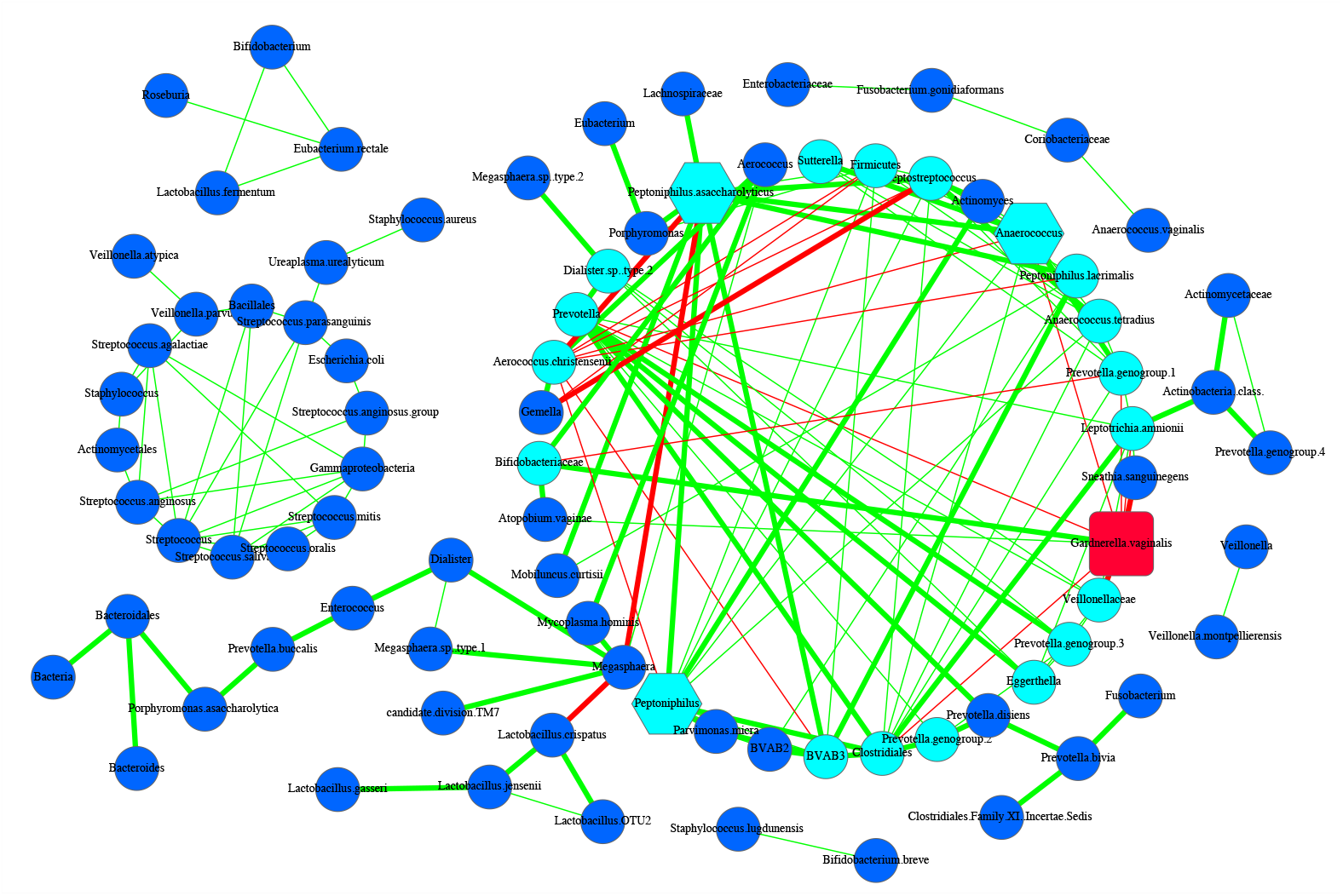
SDN network graph for Subject #s60 of the 25-mixed cohort

**Fig S1-17.**
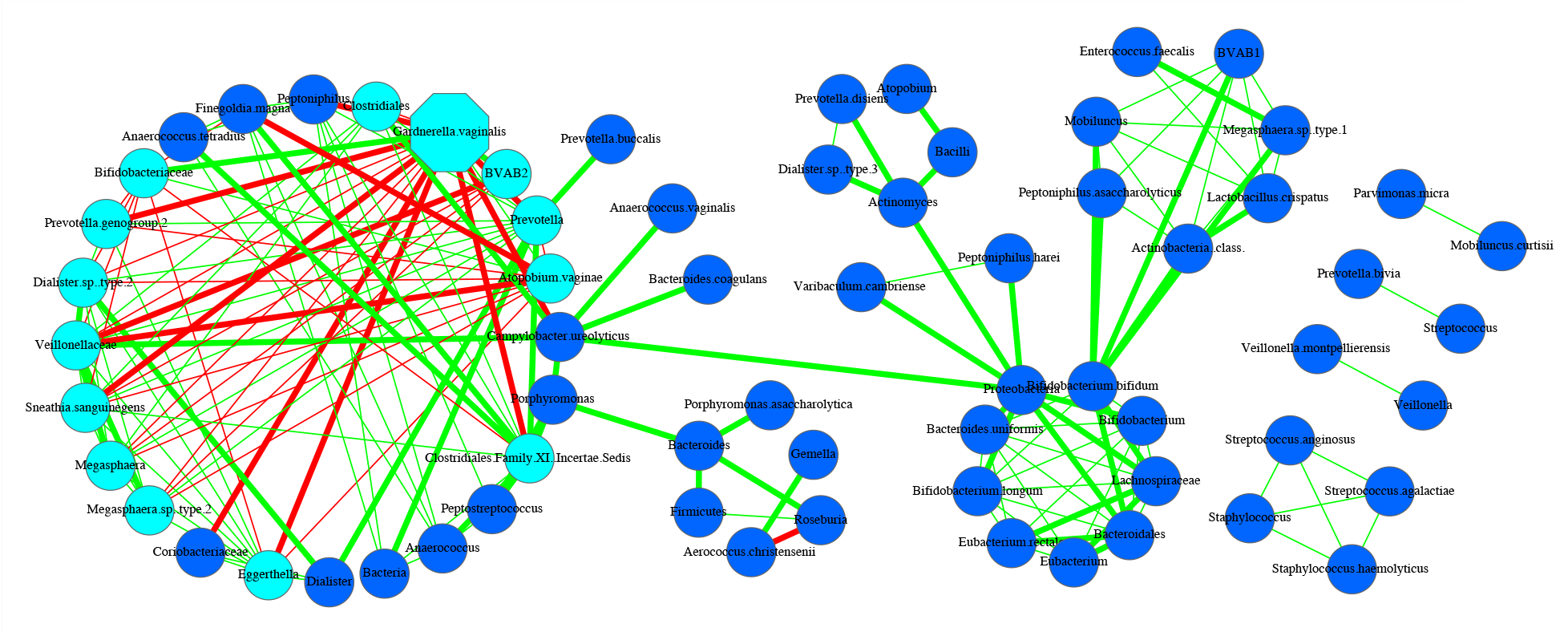
SDN network graph for Subject #s77 of the 25-mixed cohort

**Fig S1-18.**
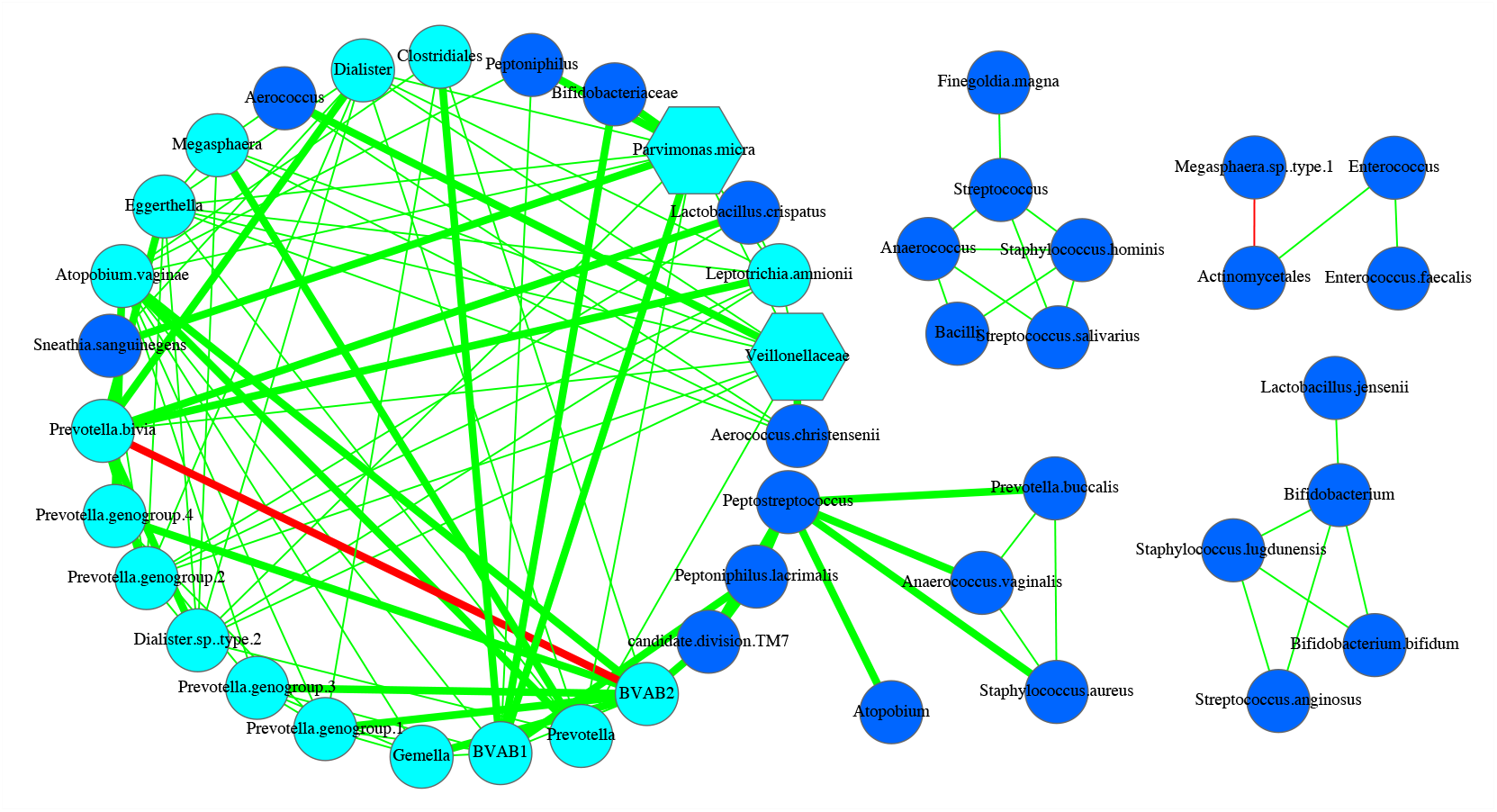
SDN network graph for Subject #s82 of the 25-mixed cohort

**Fig S1-19.**
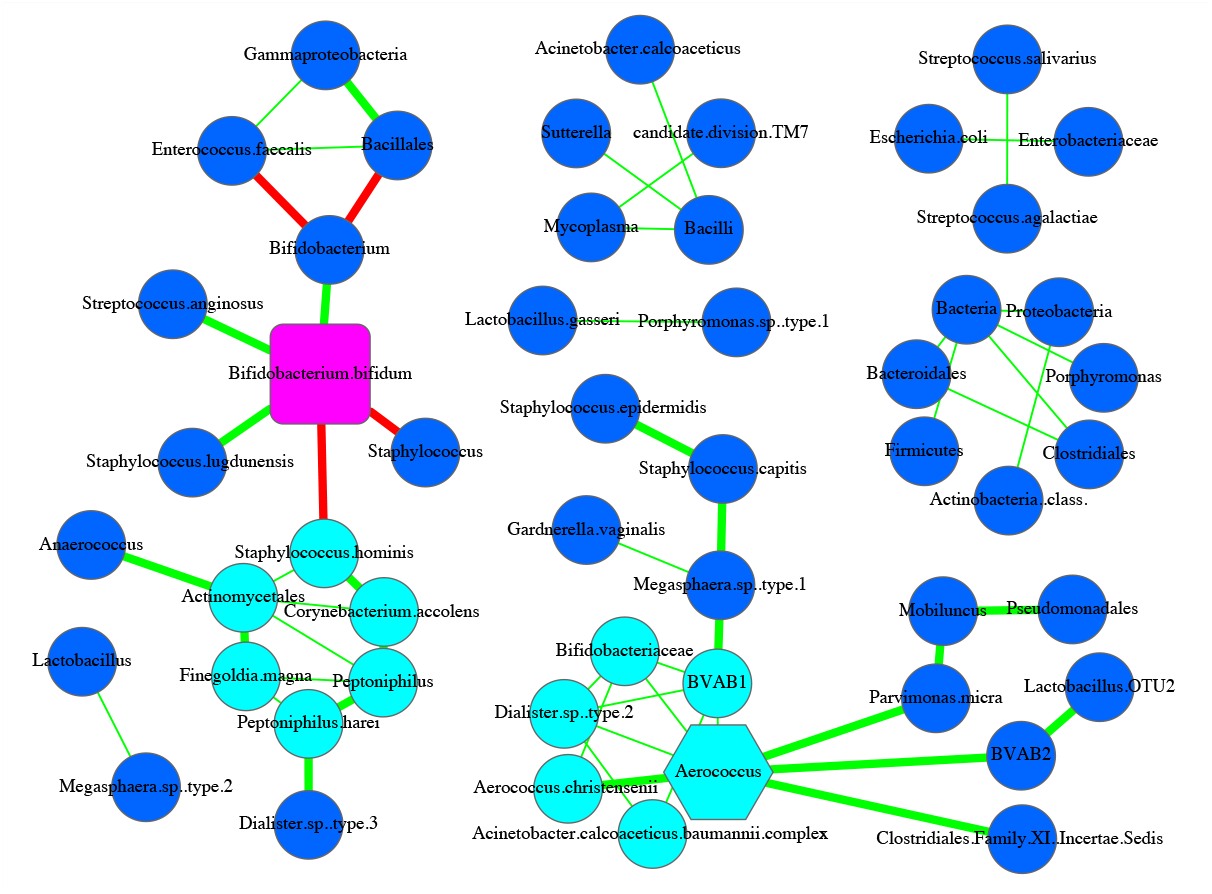
SDN network graph for Subject #s96 of the 25-mixed cohort

**Fig S1-20.**
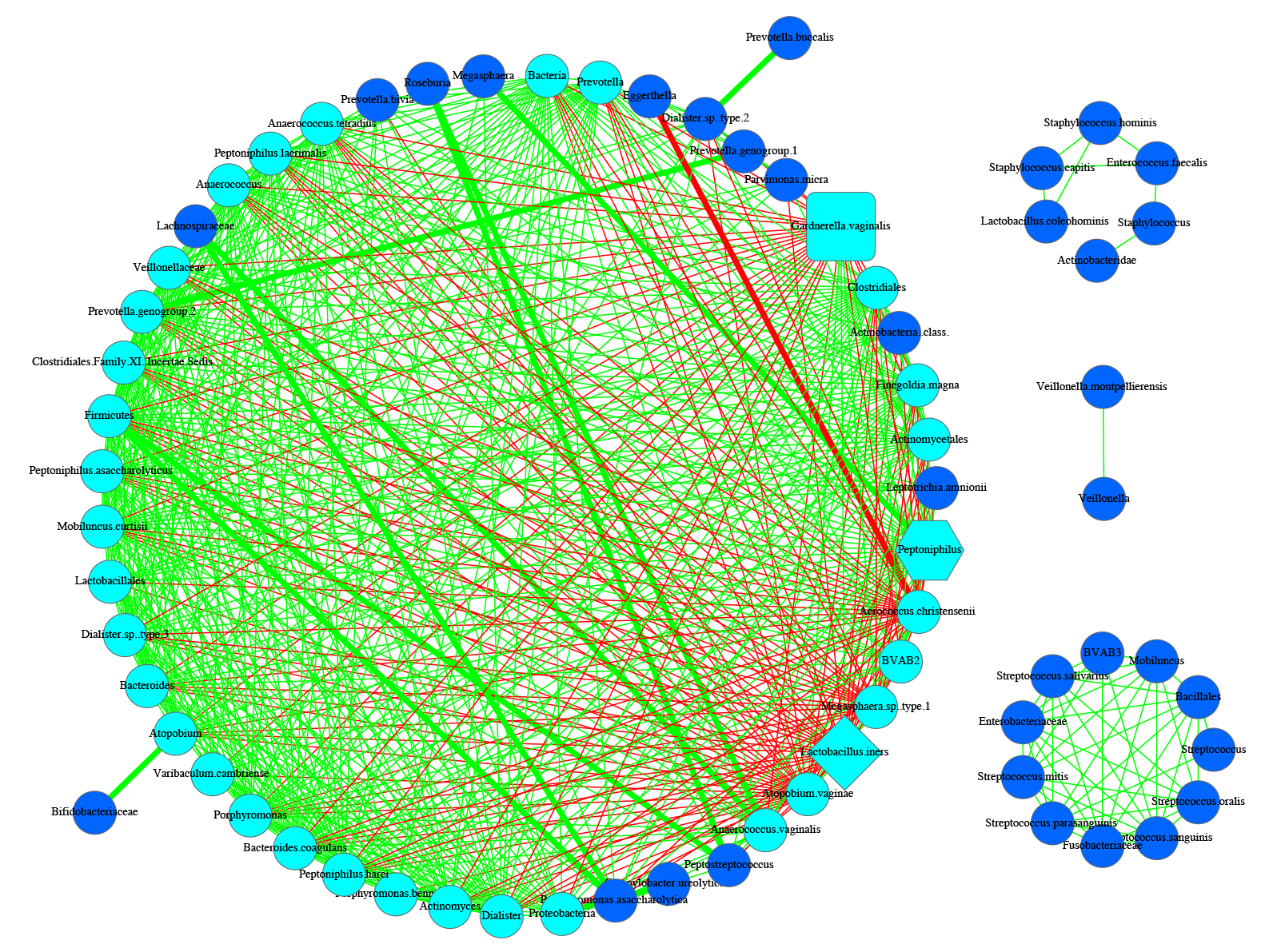
SDN network graph for Subject #s112 of the 25-mixed cohort

**Fig S1-21.**
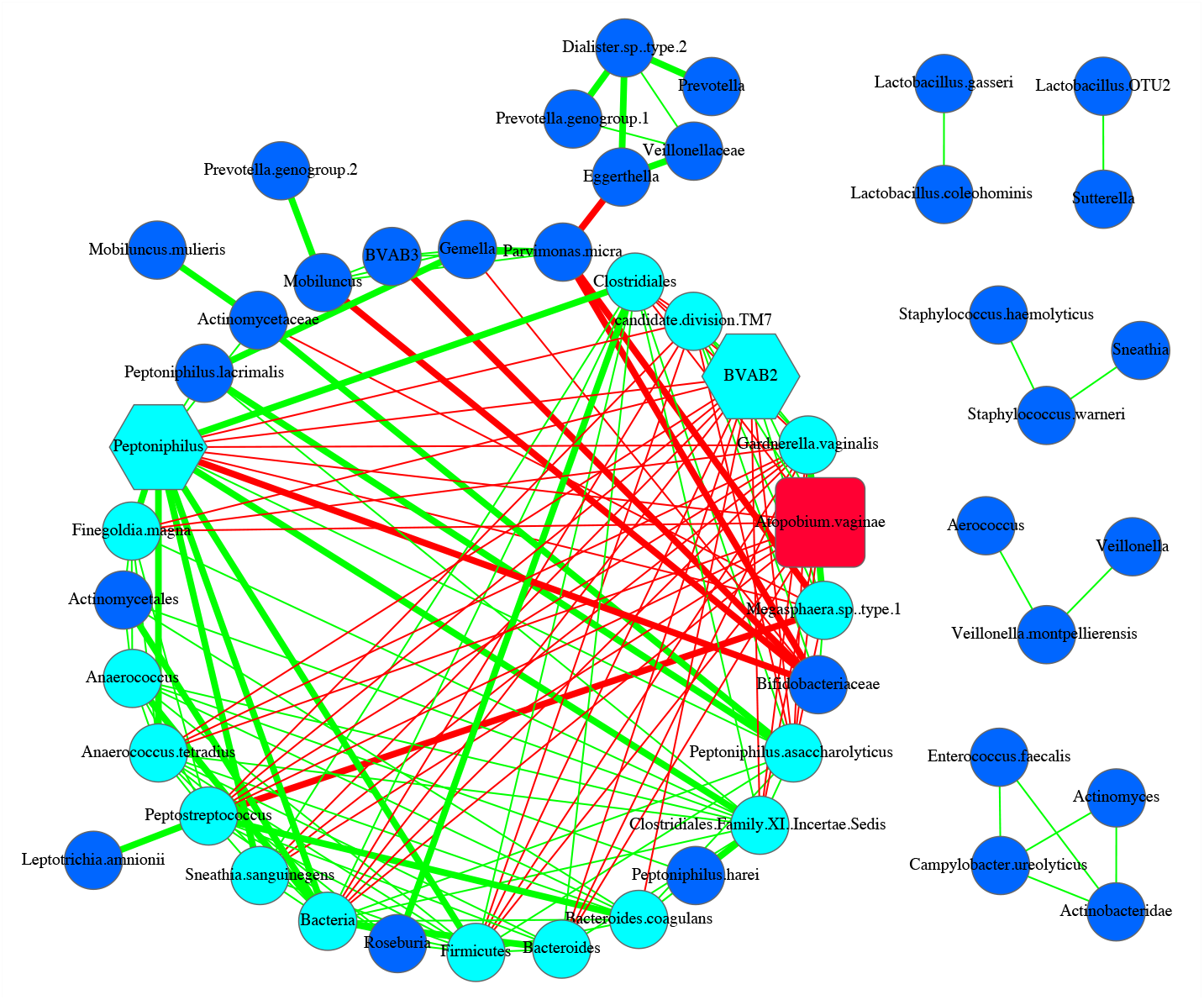
SDN network graph for Subject #s116 of the 25-mixed cohort

**Fig S1-22.**
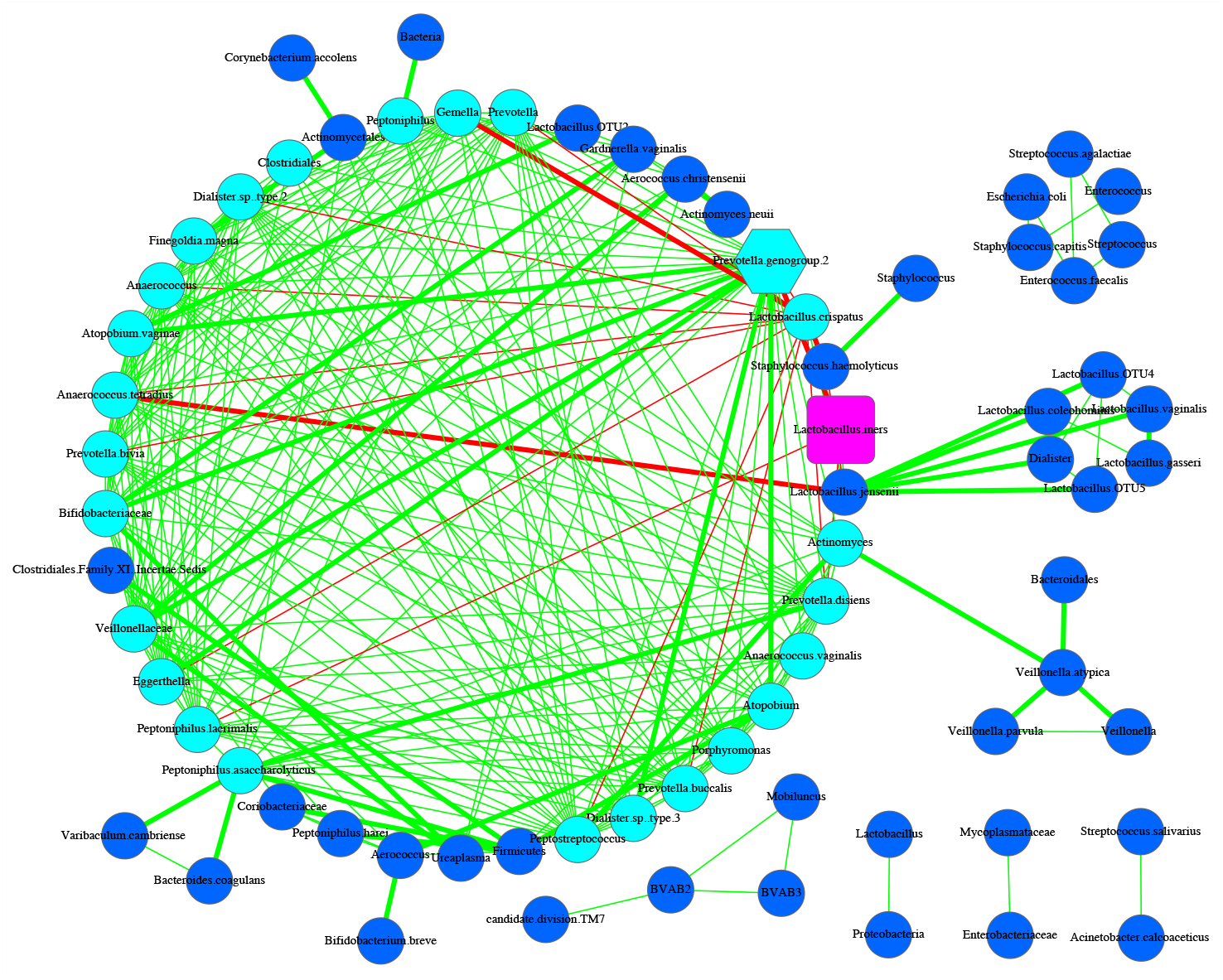
SDN network graph for Subject #s127 of the 25-mixed cohort

**Fig S1-23.**
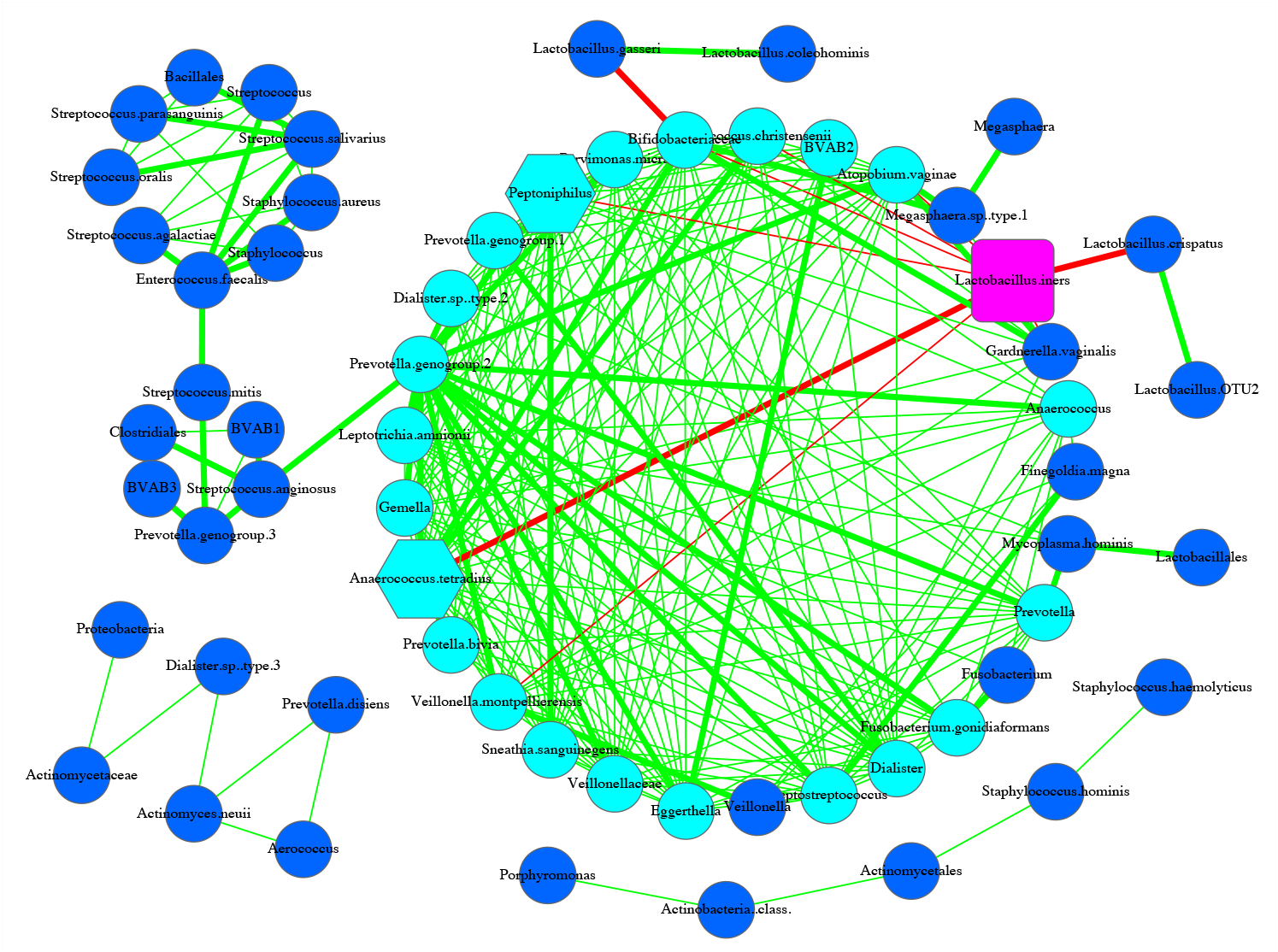
SDN network graph for Subject #s128 of the 25-mixed cohort

**Fig S1-24.**
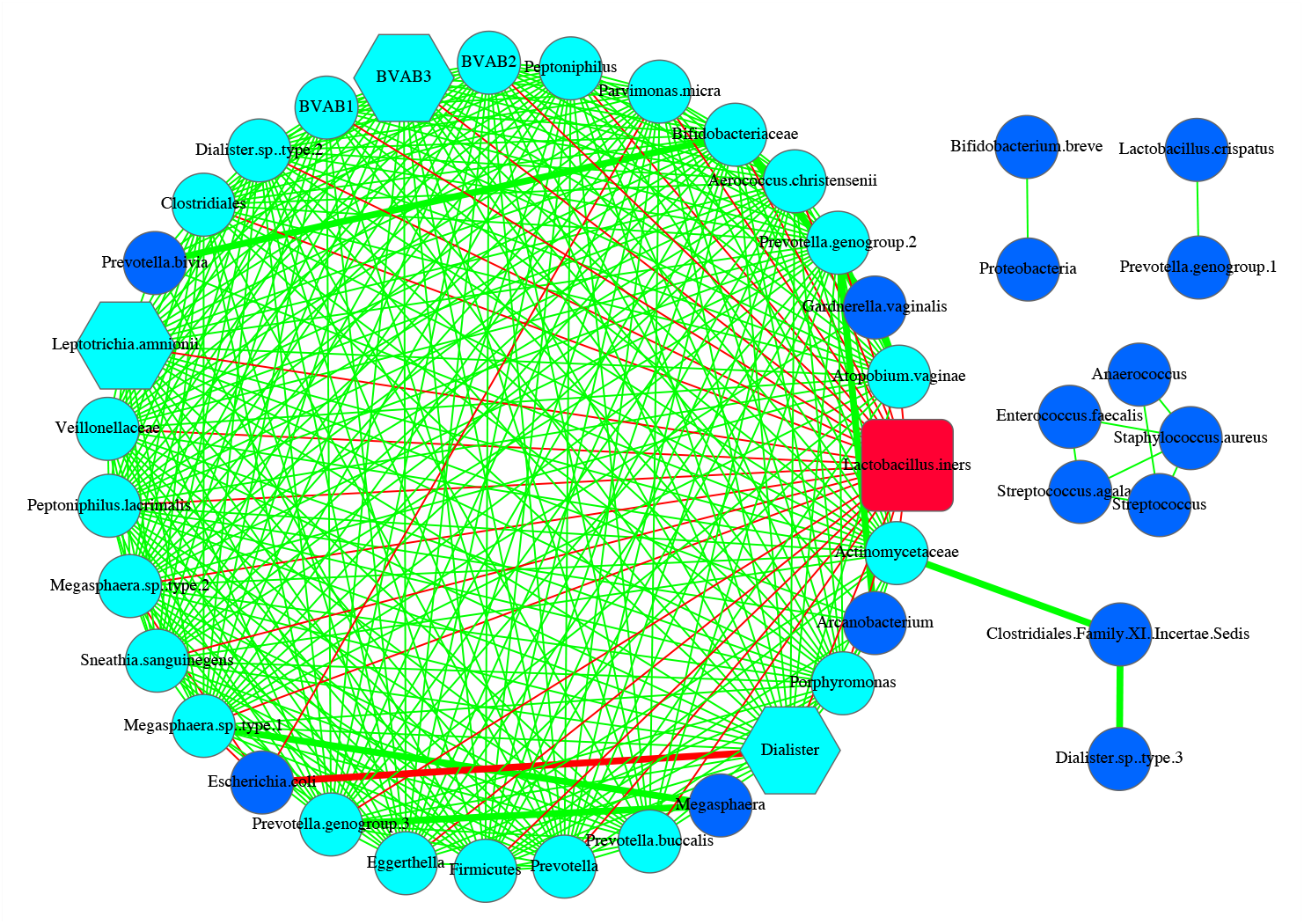
SDN network graph for Subject #s130 of the 25-mixed cohort

**Fig S1-25.**
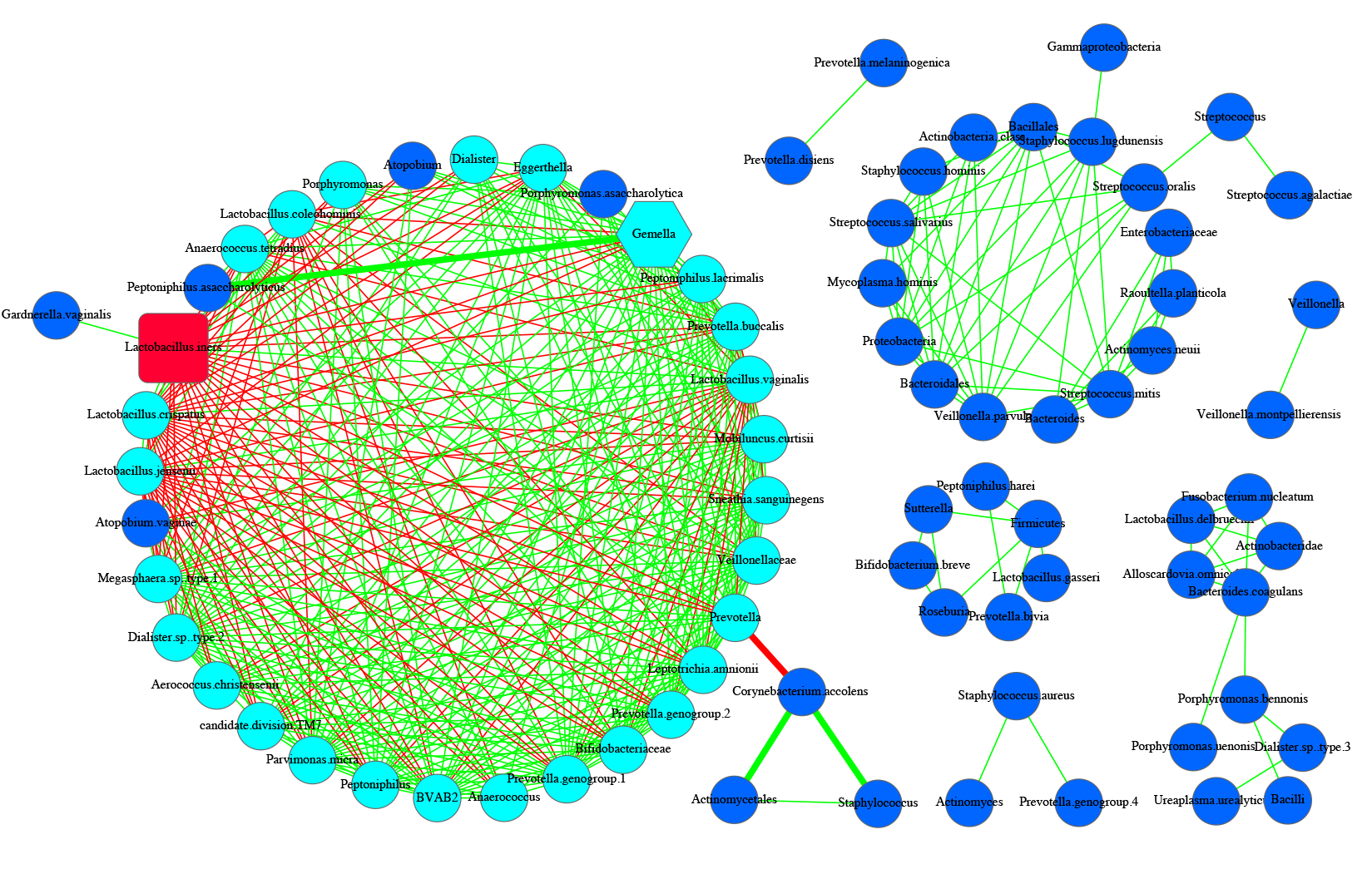
SDN network graph for Subject #s135of the 25-mixed cohort

**Fig S1.** The 25 SDN graphs for the 25 subjects in the 25-mixed cohort. Among the 25 SDNs, there are 23 subjects *(i.e*., s3, s5, s23, s49, s55, s59, s60, s77, s130) whose MDO and MAO are the same overloaded node. There is only two subject (S112 & S17) whose MDO and MAO are different nodes and nothing is overloaded *See* previous Suppl. Table S1-1 for the symbol legends.

**Fig S2.**
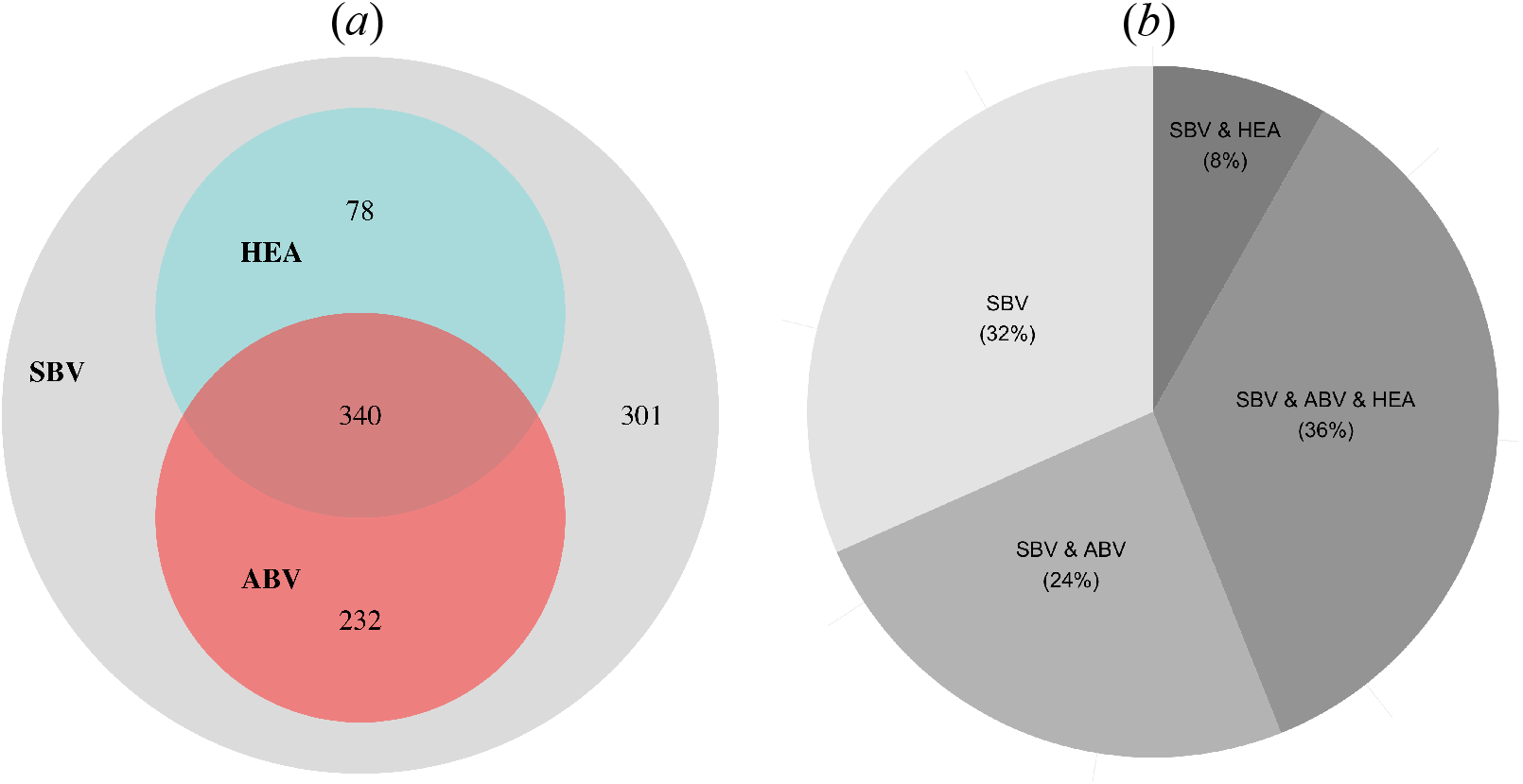
The Venn diagram and pie plot that display the distribution of 951 trios that occurred 4 or more subjects among SBV, ABV and healthy (HEA) groups: • Among 951 trios, there was no trio occurring only in ABV or HEA groups without also occurring in the other one (or two) group(s); • 301 (32%) of 951 trios occurred exclusively in SBV group; • 78 (8%) of 951 trios occurred in both SBV and HEA groups; • 232 (24%) of 951 trios occurred in both SBV and ABV groups; • 340 (36%) of 951 trios occurred in all three groups.

Online Supplementary Figures for: Ma & Ellison (2020) *“Risks and etiology of bacterial vaginosis revealed by species dominance network analysis”*

## References

Aagaard K, Riehle K, Ma J, Segata N, Mistretta TA, Coarfa C, et al. (2012) A metagenomic approach to characterization of the vaginal microbiome signature in pregnancy. PloS One 2012, 7:e36466

Bradshaw CS, Tabrizi SN, Fairley CK, Morton AN, Rudland E, Garland SM (2006) The association of *Atopobium vaginae* and *Gardnerella vaginalis* with bacterial vaginosis and recurrence after oral metronidazole therapy. J Infect Dis. 2006, 194: 828–836.

Bretelle, F, P Rozenberg, A. Pascal, R Favre, C Bohec, A Loundou et al. (2015). High Atopobium vaginae and Gardnerella vaginalis Vaginal Loads Are Associated With Preterm Birth. Clinical Infectious Diseases 60(6):860–7

Brotman RM, Bradford LL, Conrad M, Gajer P, Ault K, Peralta L, et al. (2012) Association between *Trichomonas* vaginalis and vaginal bacterial community composition among reproductive-age women. Sex Transm Dis 2012, 39: 807–812

Csermely, P, A London, LY Wu, et al (2013) Structure and dynamics of core/periphery networks. Journal of Complex Networks, 1: 93–123.

Costello EK, Stagaman K, Dethlefsen L, Bohannan BJ, Relman DA. (2012). The application of ecological theory toward an understanding of the human microbiome. Science 336(6086):1255–1262

Donohue, I., H. Hillebrand, J. M. Montoya, O. L. Petchey, S. L. Pimm, M. S. Fowler, K. Healy, A. L. Jackson, M. Lurgi, D. McClean, N. E. O’Connor, E. J. O’Gorman, and Q. Yang. 2016. Navigating the complexity of ecological stability. Ecology Letters 19: 1172–1185.

Doyle R, Gondwe A, Fan Y, et al. (2018) A Lactobacillus-Deficient Vaginal Microbiota Dominates Postpartum Women in Rural Malawi. Applied and Environmental Microbiology, 84(6): e02150–17

Eren AM, Zozaya M, Taylor CM, Dowd SE, Martin DH, Ferris MJ. (2011). Exploring the diversity of Gardnerella vaginalis in the genitourinary tract microbiota of monogamous couples through subtle nucleotide variation. PLoS One 6: e26732.

Fredricks DN, Fiedler TL, Marrazzo JM. (2005). Molecular identification of bacteria associated with bacterial vaginosis. N. Engl. J. Med. 353: 1899–911

Fredricks DN (2011) Molecular Methods to Describe the Spectrum and Dynamics of the Vaginal Microbiota. Anaerobe, 17(4): 191–195. doi:10.1016/j.anaerobe.2011.01.001.

Gajer, P., R. M. Brotman, G. Bail, J. Sakamoto, U. M. E. Schütte, X. Zhong, S. S. Koeng, L. Fu, Z. Ma, X. Zhou, Z. Abdo, L. J. Forney and J. Ravel. (2012). Temporal Dynamics of the Human Vaginal Microbiota. Science Translational Medicine, 4(132):132ra52.

Gibbs RS (2007) Asymptomatic bacterial vaginosis: is it time to treat? Am J Obstet Gynecol 2007, 196: 495–496.

Grady, D, C Thiemann, D Brockmann (2012) Robust classification of salient links in complex networks. Nature Communications, DOI: 10.1038/ncomms1847

Guedou FA, Van Damme L, Mirembe F, Solomon S, Becker M, Deese J, et al. (2012) Intermediate vaginal flora is associated with HIV prevalence as strongly as bacterial vaginosis in a cross-sectional study of participants screened for a randomized controlled trial. Sex Transm Infect 2012, 88: 545–551.

Hickey RJ, Zhou X, Pierson JD, Ravel J, Forney LJ. (2012). Understanding vaginal microbiome complexity from an ecological perspective. Transl. Res, 2012, 160: 267–282.

Jespers V, Menten J, Smet H, Poradosu S, Abdellati S, Verhelst R, et al. (2012) Quantification of bacterial species of the vaginal microbiome in different groups of women, using nucleic acid amplification tests. BMC Microbiol 2012, 12: 83.

Koumans, EH, Kendrick, JS. 2001. Preventing adverse sequelae of bacterial vaginosis: a public health program and research agenda. Sexually Transmitted Diseases, 28(5):292–297.

Lambert JA, Kalra A, Dodge CT, et al. (2013) Novel PCR-Based Methods Enhance Characterization of Vaginal Microbiota in a Bacterial Vaginosis Patient before and after Treatment. Applied and Environmental Microbiology, 79(13): 4181–4185

Lamont RF, Taylor-Robinson D (2010) The role of bacterial vaginosis, aerobic vaginitis, abnormal vaginal flora and the risk of preterm birth. BJOG 2010, 117: 119–120.

Ling Z, Kong J, Liu F, Zhu H, Chen X, Wang Y, et al. (2010) Molecular analysis of the diversity of vaginal microbiota associated with bacterial vaginosis. BMC Genom 2010, 11: 488.

Ma, B, LJ Forney, J. Ravel. 2012. The Vaginal Microbiome: Rethinking Health and Disease. Annual Review of Microbiology. 66:371–389. doi: 10.1146/annurev-micro-092611-150157

Ma, ZS, Abdo Z, Forney L (2011) Caring about trees in the forest: incorporating frailty in risk analysis for personalized medicine. Personalized Medicine 8: 681–688.

Ma, Z.S. et al (2015) Network analysis suggests a potentially ‘evil’ alliance of opportunistic pathogens inhibited by a cooperative network in human milk bacterial communities. Sci. Rep. 5, 8275; DOI: 10.1038/srep08275 (2015).

Ma ZS & DD Ye (2017) Trios—promising in silico biomarkers for differentiating the effect of disease on the human microbiome network. Scientific Reports, vol. 7. Article No. 13259

Ma ZS & LW Li (2017) Quantifying the human vaginal community state types (CSTs) with the species specificity index. PeerJ, 5:e3366 https://doi.org/10.7717/peerj.3366

Ma ZS and AM Ellison (2018) A unified concept of dominance applicable at both community and species scales. Ecosphere 9(10):e02477. 10.1002/ecs2.2477

Ma ZS and AM Ellison (2019). Dominance network analysis provides a new framework for studying the diversity-stability relationship. Ecological Monographs, vol. 89(2): e01358, https://doi.org/10.1002/ecm.1358

Ma ZS, Li LW, Gotelli NJ (2019) Diversity-disease relationships and shared species analyses for human microbiome-associated diseases, The ISME Journal. https://www.nature.com/articles/s41396-019-0395-y

Mania-Pramanik J, Kerkar SC, Salvi VS (2009) Bacterial vaginosis: a cause of infertility? Int J Std Aids 2009, 20: 778–781

Menard JP, Fenollar F, Henry M, Bretelle F, Raoult D (2008) Molecular quantification of Gardnerella vaginalis and Atopobium vaginae loads to predict bacterial vaginosis. Clin Infect Dis 2008, 47: 33–43.

Pleckaityte M, M. Zilnyte and A. Zvirbliene. (2012) Insights into the CRISPR/Cas system of *Gardnerella vaginalis*. BMC Microbiology 12:301. doi: 10.1186/1471-2180-12-301

Randis TM, Ratner AJ. (2019) Gardnerella and Prevotella: Co-conspirators in the Pathogenesis of Bacterial Vaginosis. The Journal of Infectious Diseases, doi: 10.1093/infdis/jiy705

Ravel J., Gajer, P., Abdo, Z., Schneider, G. M., Koenig, S. S. K., McCulle, S. L., Karlebach, S., et al. (2011). Vaginal microbiome of reproductive-age women. Proceedings of the US National Academy of Sciences, 108 Suppl. 1, 4680–4687.

Ravel, J. Jacques Ravel, Rebecca M Brotman, Pawel Gajer, Bing Ma, Melissa Nandy, Douglas W Fadrosh, Joyce Sakamoto, Sara SK Koenig, Li Fu, Xia Zhou, Roxana J Hickey, Jane R Schwebke, Larry J Forney (2013) Daily temporal dynamics of vaginal microbiota before, during and after episodes of bacterial vaginosis. Microbiome 2013, 1: 29

Romero, R, SS Hassan, P. Gajer, AL Tarca, DW Fadrosh, L. Nikita, M. Galuppi, RF Lamont, P. Chaemsaithong, J. Miranda, T. Chaiworapongsa and J. Ravel (2014) The composition and stability of the vaginal microbiota of normal pregnant women is different from that of non-pregnant women. Microbiome, 2014, 2:4 doi:10.1186/2049-2618-2-4

Shekhtman, LM, JP Bagrow, & D Brockmann (2014) Robustness of skeletons and salient features in networks. Journal of Complex Networks, doi:10.1093/comnet/cnt019

Sobel JD (1999) Is there a protective role for vaginal flora? Curr. Infect. Dis. Rep. 1: 379–83

Spear GT, St John E, Zariffard MR (2007) Bacterial vaginosis and human immunodeficiency virus infection. AIDS Res Ther 2007, 4: 25

Srinivasan S, Liu C, Mitchell CM, Fiedler TL, Thomas KK, Agnew KJ, et al. (2010) Temporal variability of human vaginal bacteria and relationship with bacterial vaginosis. 2010, PloS One 2010, 5:e10197.

White, BA, D. J. Creedon, K. E. Nelson, B. A. Wilson (2011) The vaginal microbiome in health and disease. Trends in Endocrinology and Metabolism, 2011, 22(1): 389–393

